# Long-term cardiac symptoms following COVID-19: a systematic review and meta-analysis

**DOI:** 10.1101/2023.01.16.23284620

**Authors:** Boya Guo, Chenya Zhao, Mike Z. He, Camilla Senter, Zhenwei Zhou, Jin Peng, Song Li, Annette L Fitzpatrick, Sara Lindström, Rebecca C Stebbins, Grace A Noppert, Chihua Li

## Abstract

**Background:** There is growing body of literature on the long-term cardiac symptoms following COVID-19. We conducted a systematic review and meta-analysis to synthesize and evaluate related evidence to inform clinical management and future studies.

**Methods:** We searched two preprint and seven peer-reviewed article databases from January 1, 2020 to January 8, 2022 for studies investigating cardiac symptoms that persisted for at least 4 weeks among individuals who survived COVID-19. A customized Newcastle–Ottawa scale was used to evaluate the quality of included studies. Random-effects meta-analyses were performed to estimate the proportion of symptoms with 95% confidence intervals (CI), and stratified analyses were conducted to quantify the proportion of symptoms by study characteristics and quality.

**Results:** A total of 101 studies describing 49 unique long-term cardiac symptoms met the inclusion criteria. Based on quality assessment, only 15.8% of the studies (n=16) were of high quality, and most studies scored poorly on sampling representativeness. The two most examined symptoms were chest pain and arrhythmia. Meta-analysis showed that the proportion of chest pain was 10.1% (95% CI: 6.4-15.5) and arrhythmia was 9.8% (95% CI: 5.4-17.2). Stratified analyses showed that studies with low-quality score, small sample size, unsystematic sampling method, and cross-sectional design were most likely to report high proportions of symptoms. For example, the proportion of chest pain was 21.3% (95% CI: 10.5-38.5), 9.3% (95% CI: 6.0-14.0), and 4.0% (95% CI: 1.3-12.0) in studies with low, medium, and high-quality scores, respectively. Similar patterns were observed for other cardiac symptoms including hypertension, cardiac abnormalities, myocardial injury, thromboembolism, stroke, heart failure, coronary disease, and myocarditis.

**Discussion:** There is a wide spectrum of long-term cardiac symptoms following COVID-19. Findings of existing studies are strongly related to study quality, size and design, underscoring the need for high-quality epidemiologic studies to characterize these symptoms and understand their etiology.

**Research in context:** *Evidence before this study:* Accumulating evidence shows long-term cardiac symptoms following COVID-19. However, no previous reviews systematically evaluated and synthesized findings from studies on long-term cardiac symptoms.

*Added value of this study:* This is the first systematic review and meta-analysis that focused on studies of long-term cardiac symptoms of COVID-19. We included 101 studies and identified 49 cardiac symptoms that are indicative of cardiac abnormalities. We scored their quality based on epidemiologic principles and identified domains of study design that need improvements. We quantified proportions of multiple long-term cardiac symptoms including chest pain, arrhythmia, and others. We also observed systematic differences in reported proportions of these symptoms by selected study characteristics, including total quality assessment score, sample size, sampling representativeness, and study design. High-quality studies identified here can provide important guidelines for future studies of long-term symptoms following COVID-19.

*Implications of all the available evidence:* Multiple domains of study design, especially sampling representativeness, need to be improved in future studies on long-term cardiac symptoms following COVID-19. Notably, low-quality and smaller studies tend to report a larger proportion of symptoms, are more likely to be subject to greater sampling variation, and hence are less precise. These studies should be revisited with the emergence of large studies with rigriours study designs. This systematic review and meta-analysis highlight the scope of persistent cardiac symptoms among those who survived the acute phase of COVID-19, and the importance of synthesizing rigorous evidence to inform post-COVID surveillance and management plans.

## Background

Coronavirus disease 2019 (COVID-19) infection affects multiple organs and is associated with a wide spectrum of persistent symptoms after the acute phase.^1^ While definitions of long-term COVID-19 symptoms vary, the National Institutes of Health (NIH) and Centers for Disease Control and Prevention (CDC) describe them as a wide range of new, returning, or ongoing health problems that extend beyond four weeks after initial infection.^1–3^ These long-term symptoms include cardiovascular, respiratory, neurological, otorhinolaryngological, and other complications. Many of these long-term cardiac complications mirror those encountered by survivors of Severe Acute Respiratory Syndrome (SARS), Middle East Respiratory Syndrome (MERS), and H1N1A influenza, underscoring the need to recognize the long-term impact of COVID-19 on cardiovascular health.^4–6^

Accumulating evidence shows that long-term cardiac symptoms of COVID-19 are common and can last for months and even beyond one year.^7–9^ Commonly observed symptoms include chest pain, arrhythmia, inflammatory heart disease, and others.^7,10^ Myocarditis, or inflammation of the heart muscle, which can be an acute consequence of viral infection can also evolve into overt or subclinical myocardial dysfunction and/or electrophysiological abnormalities with long-term implications.^11^ The severity and duration of these long-term cardiac symptoms vary by patients, and the identification of their risk factors remains an active and critical line of investigation.

Research on these topics can inform surveillance and management of these symptoms and therefore lower the burden of subsequent morbidity and mortality of patients who survived COVID-19.

To date, multiple reviews on long-term symptoms of COVID-19 have been conducted to synthesize relevant literature.^10–20^ They summarized observed long-term symptoms, quantified their prevalence, and discussed potential biological mechanisms. However, none of them focused on cardiac symptoms and nor did they include comprehensive documentation of related studies. Further none of the reviews have performed a systematic examination of reported findings and assessed their quality from the perspective of epidemiologic principles. This is important because findings on long-term cardiac symptoms of COVID-19 from published literature are heterogeneous and the methodological quality of related studies varies substantially.

In this systematic review and meta-analysis, we focus on studies that examined long-term cardiac symptoms of COVID-19. We summarize their study characteristics and main findings and perform a quality assessment using epidemiologic principles. We also conduct a meta-analysis to identify patterns of reported findings. We aim to synthesize available evidence on long-term cardiac symptoms of COVID-19, identify knowledge gaps, highlight existing methodological concerns, and formulate recommendations for future studies. It is imperative for clinicians and healthcare providers to have this information, understand the scope of long-term cardiac symptoms, and develop scientific surveillance and management plans for patients with post-COVID conditions.

## Methods

Our systematic review followed the Preferred Reporting Items for Systematic Reviewers and Meta-analysis (PRISMA) guidelines (**Table S1**).^21^ Long-term COVID-19 was defined as symptoms that last for 4 weeks and beyond from the index of date of follow-up, including symptom onset/time at first diagnosis, hospitalization admission, and hospital discharge. The study is registered on Research Registry (Research Registry unique identifying number: reviewregistry1538) with a detailed protocol.

### Search strategy and selection criteria

The initial search was conducted from January 1, 2020, to May 10, 2021 in seven electronic databases (PubMed, Embase, Web of Science, CINAHL Cochrane Library, Scopus, and PsycINFO), and two preprint servers (medRxiv and bioRxiv). We searched for both preprints and peer-reviewed articles that were published. The following broad search terms and keywords were used: (“COVID-19” OR “SARS-CoV-2” OR “COVID” OR “COVID19” OR “2019-nCoV” OR “Coronavirus” OR “SARS-CoV-2” OR “severe acute respiratory syndrome coronavirus-2”) AND (”post-acute COVID-19 syndrome” OR “post-acute” OR “sequelae” OR “long-term symptoms” OR “long-COVID” OR “post-COVID syndrome” OR “post-COVID symptoms” OR “hauler” OR “long-haul” OR “lingering” OR “chronic covid” OR “chronic symptoms” OR “persistent” OR “recurrent” OR “recurring” OR “complication” OR “subacute”). To include the up-to-date articles and have a manageable number of articles to screen, we conducted a second-round article search in PubMed, and searched for peer-reviewed articles published on PubMed from May 11, 2021, to January 8, 2022, using the same search terms as the previous data pull and including cardiac related terms: “heart” OR “chest” OR “cardiovascular” OR “myocardial” OR “cardiac” OR “palpitation” OR “tachycardia” OR “arrhythmia” OR “myocarditis”.

There were no language restrictions for articles included. The exclusion criteria were as follows: 1) reviews, commentaries, abstracts only, posters, or conference proceedings; or 2) studies having a follow-up duration of fewer than four weeks (28 days) or reporting no information on follow-up duration; or 3) studies with a sample size of less than 30 participants; or 4) cases with long-term cardiac symptoms among individuals who had not tested positive for COVID-19; or 5) non-human studies; or 6) study participants who were not alive at the time of the study (ex. post-mortem examination); or 7) studies that only presented data from modeling outputs or did not use primary data (i.e., secondary data, or summarize findings from previously published papers); or 8) no relevant symptoms or only reported biomarkers rather than symptoms. When several studies were based on the same or overlapping study participants, the study with the largest sample size was used as the representative study.

### Screening process and data extraction

The article screening process was conducted using Covidence, a web-based collaboration software platform that streamlines the production of systematic and other literature reviews.^22^ At least two reviewers independently screened the title and abstract of each study for full-text examination. Two reviewers further conducted the full-text examination to evaluate if the study met the eligibility criteria. Any disagreements among reviewers were resolved through group discussion. For articles meeting the inclusion criteria, the following information was extracted by a single reviewer to a spreadsheet and checked by another reviewer: 1) author and publication information; 2) study characteristics (study country, study period, patient diagnosis/recruitment period, study design, COVID-19 diagnosis tools, study setting, study population, sample size, start of follow-up time, outcome assessment time points, outcome assessment method); 3) participants characteristics (age, sex, comorbidities or symptoms at baseline, COVID-19 treatment, vaccination status, hospitalization status, severity); and 4) results (prevalence of the outcome, outcome duration) (**Table S2 and S3**).

### Quality assessment

A modified Newcastle–Ottawa scale (NOS)^23^ was developed to evaluate quality of seven items for each included study: sampling representativeness, sample size, exposure assessment, outcome assessment, covariate assessment, follow-up, and statistics analysis (**Table S4**). The quality of each item was scored as ‘good’ (2), ‘fair’ (1), or ‘poor’ (0), and a total score was calculated (range: 0-14). A study with a total score of 11 to 14 was classified as of ‘high’ quality, a score of 7 to 10 was classified as of ‘medium’ quality, and a score of 0 to 6 as ‘low’ quality.

Two reviewers appraised each study independently, and disagreements were resolved through consensus.

### Data synthesis and statistical analysis

All analyses were performed using R (version 4.2.2). We described the characteristics of included studies and presented proportions of commonly reported cardiac symptoms, calculated as the number of COVID-19 survivors who reported a specific cardiac symptom divided by the total number of COVID-19 survivors. Meta-analysis was performed using the *meta* package in R using both fixed-effects and random-effects models to estimate the pooled proportion of long-term cardiac symptoms following COVID-19 and 95% confidence intervals (CI). Heterogeneity between studies was assessed using the τ^2^ (between-study variance) and I^2^ (total proportion of variance owing to heterogeneity) statistics. The Cochrane Q-test was used to determine statistical significance. Studies that did not report the number of COVID-19 survivors with long-term symptoms and/or size of the total study population were excluded from meta-analysis, and were only included in the systematic review. Case-control studies were not included in the meta-analysis because the proportion of each symptom was determined by study design. Additional stratified analyses were performed based on quality assessment score (low, medium, high), sample size (30-99, 100-999, ≥ 1000), sampling representativeness (online survey and single hospital, multiple hospitals, national studies), and study design (cross-sectional, retrospective cohort, prospective cohort) to examine how each long-term cardiac symptom was distributed by these study characteristics. Funnel plots were plotted using the log transformation of the proportion of the symptom against the standard error of proportion to explore the presence of publication bias, followed by Egger’s test to assess funnel plot asymmetry.

### Findings

#### Study selection

The initial search of studies published between January 2020 and May 2021 yielded 40,215 records from electronic databases (**Figure 1**). After the removal of 17,176 duplicates and the screening of titles and abstracts of 23,039 unique records, 321 studies underwent full-text examinations, and 65 studies met the inclusion criteria. The second-round search of studies published between May 2021 and January 2022 yielded 3,830 new records, among which a total of 36 studies met the inclusion criteria. In total, 101 studies on long-term cardiac symptoms following COVID-19 were included in this systematic review, and 92 studies provided data that could be used in the meta-analysis.

**Figure 1.**
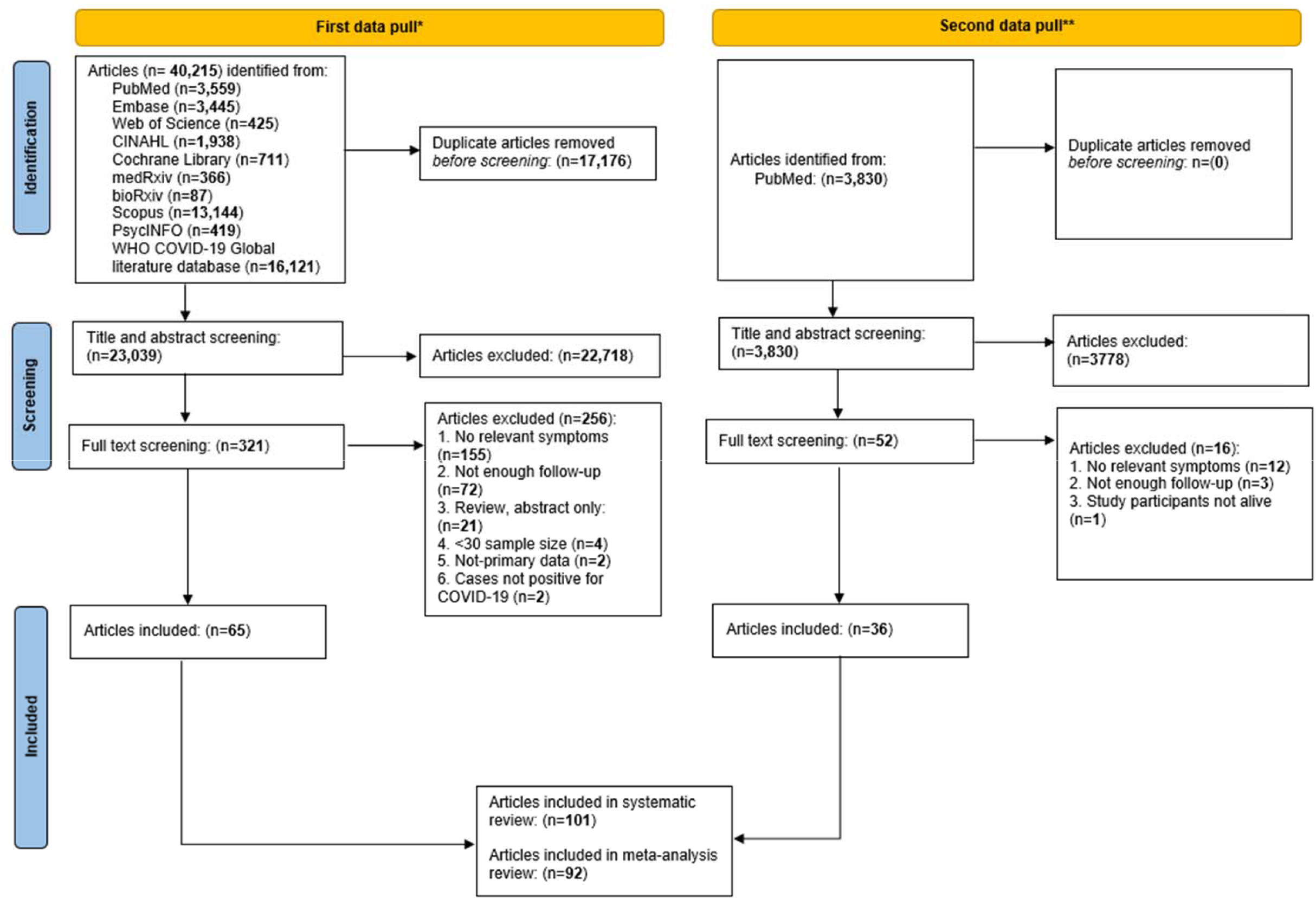
PRISMA flow diagram for systematic reviews.

#### Study characteristics

**Table 1** summarizes the characteristics of included studies. In total, 49 different long-term cardiac symptoms following COVID-19 were examined across these studies, among which chest pain, and arrhythmia were the two most widely examined symptoms (**Table S5**). Over two-thirds of studies (n=68) were prospective cohorts, 14 were retrospective cohorts, and 15 were cross-sectional studies. Included studies were mostly online surveys and single-hospital studies (n=65) or multiple-hospital and regional studies (n=30). Around 60% (n=59) had a sample size of 100-999 participants, and 28 studies had a sample size of at least 1,000 individuals. Most studies had COVID-19 cases confirmed clinically (n=76). Long-term cardiac symptoms were either clinically evaluated (n=42) or self-reported (n=56). Studies varied in index dates at which time follow-up of participants began. Forty-five studies recruited inpatients, 15 recruited outpatients, and 35 studies recruited both. Most studies (n=63) included COVID-19 cases regardless of severity, and tools used to evaluate severity varied substantially between studies.

**Table 1.** Summary of study characteristics.

| **Study characteristics** | **N (%)** |
| --- | --- |
| **Publication year** |  |
| 2020 | 11 (10.9) |
| 2021 and early 2022 | 90 (89.1) |
| **Preprint** |  |
| Yes | 10 (9.9) |
| No | 91 (90.1) |
| **Study design** |  |
| Cross-sectional | 15 (14.9) |
| Prospective cohort | 68 (67.3) |
| Retrospective cohort | 14 (13.7) |
| Other designs * | 4 (4.1) |
| **Sampling representativeness** |  |
| Online surveys and single-hospital studies | 65 (64.4) |
| Multiple-hospital and regional studies † | 30 (29.7) |
| National studies | 6 (5.9) |
| **Continent** |  |
| America | 27 (26.7) |
| Asia | 15 (14.9) |
| Europe | 54 (53.5) |
| Other continents § | 5 (4.9) |
| **Sample size** |  |
| 30-99 | 14 (13.9) |
| 100-999 | 59 (58.4) |
| ≥1000 | 28 (27.7) |
| **Age group #** |  |
| Children (0-14) | 6 (5.9) |
| Youth (15-24) | 1 (1.0) |
| Adults (25-64) | 76 (75.3) |
| Senior (65+) | 11 (10.9) |
| Not reported | 7 (6.9) |
| **COVID-19 testing method** |  |
| Clinically confirmed ¶ | 76 (75.2) |
| Self-reported | 6 (5.9) |
| Not reported | 19 (18.9) |
| **Symptom assessment method** |  |
| Clinically evaluated | 42 (41.6) |
| Self-reported | 56 (55.4) |
| Others and not reported | 3 (3.0) |
| **Index date of follow-up** |  |
| Symptom onset/time at first diagnosis | 31 (30.7) |
| Hospitalization admission | 54 (53.5) |
| Hospitalization discharge | 12 (11.9) |
| Others and not reported | 4 (3.9) |
| **Hospitalization status** |  |
| Inpatient | 45 (44.5) |
| Outpatient | 15 (14.9) |
| Mixed | 35 (34.7) |
| Not reported | 6 (5.9) |
| **COVID-19 severity of cases** |  |
| Mild | 3 (3.0) |
| Moderate to severe | 63 (62.4) |
| Severe | 4 (4.0) |
| Not reported | 31 (30.6) |

\* Other designs include study designs of case-control, case-series, and ambidirectional cohorts
† Two single-hospital studies were classified as regional studies as COVID-19 cases captured by them well-represented populations infected in their regions.
§ Other continents include studies conducted in Africa and Australia, and studies conducted in more than one continent

#### Quality assessment

**Figure 2A** shows the number of studies by the total score of quality assessment. Only 16 studies out of 101 studies received a total score of 11 to 14, considered to be of ‘high’ quality; 66 studies received a total score of 7 to 10, which were of ‘medium’ quality; and the remaining 19 studies received a total score of 6 and below, considered of ‘low’ quality. **Figure 2B** summarizes the proportion of studies receiving a score of ‘good’ (2), ‘fair’ (1), and ‘poor’ (0) for each of the seven quality assessment items. Across seven quality assessment items, the proportion of studies scoring ‘good’ was highest for exposure assessment and lowest for sampling representativeness and follow-up. A detailed quality assessment for each study by domain is presented in **Figure S1**.

**Figure 2.**
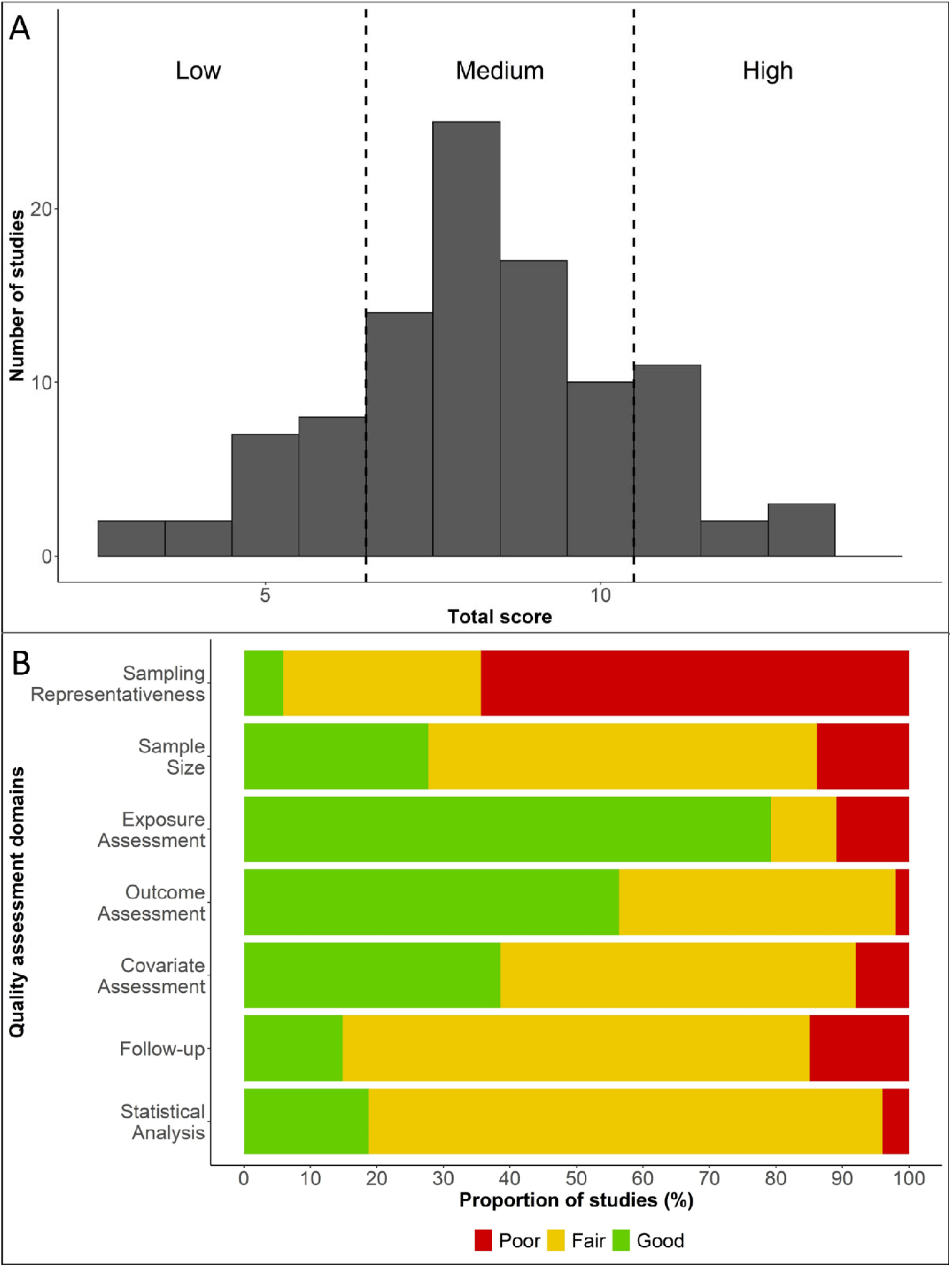
Quality assessments of the included studies.

#### Meta-analysis

In total, ten cardiac symptoms were reported by five and more studies, including chest pain (n=70), arrhythmia (n=36), hypertension (n=10), cardiac abnormalities (n=10), myocardial injury (n=7), thromboembolism (n=6), stroke (n=6), heart failure (n=6), coronary disease (n=5), and myocarditis (n=5) (**Table S5**). Fixed-effects and random-effects meta-analyses were conducted for these symptoms (**Figure S2**). Substantial heterogeneity across studies was observed for all symptoms. The pooled proportion for each of the symptoms using a random-effects model was as following: chest pain (10.06%, 95% CI: 6.40-15.47), arrhythmia (9.80%, 95% CI: 5.39-17.18), hypertension (6.07%, 95% CI: 3.42-10.53), cardiac abnormalities (0.05%, 95% CI: 0.03-0.08), myocardial injury (2.63%, 95% CI: 0.13-36.30), thromboembolism (2.68%, 95% CI: 0.83-8.37), stroke (0.71%, 95% CI: 0.18-2.84), heart failure (1.23%, 95% CI: 0.41-3.59), coronary disease (0.41%, 95% CI: 0.23-0.70), and myocarditis (0.62%, 95% CI: 0.08-4.44).

Stratified analyses were conducted using the following characteristics: total quality assessment score, sample size, sampling representativeness, and study design. Heterogeneity remained high for most symptoms within the stratum after stratification. **Figure 3** summarizes results of stratified analyses for chest pain and arrhythmia. For both symptoms, studies with the lowest quality score, smallest sample size, poorest sampling schemes, and cross-sectional designs reported the highest proportion of the symptom. For example, the proportion of chest pain among individuals survived COVID-19 was 21.32% (95% CI: 10.50-38.50) for low quality score studies, 9.26% (95% CI: 6.04, 13.95) for medium quality score studies, and 4.04% (95% CI: 1.28, 11.98) for high quality score studies. We observed similar patterns for other symptoms, although a small number of studies within some strata precluded formal analyses **(Figure S3)**.

**Figure 3.**
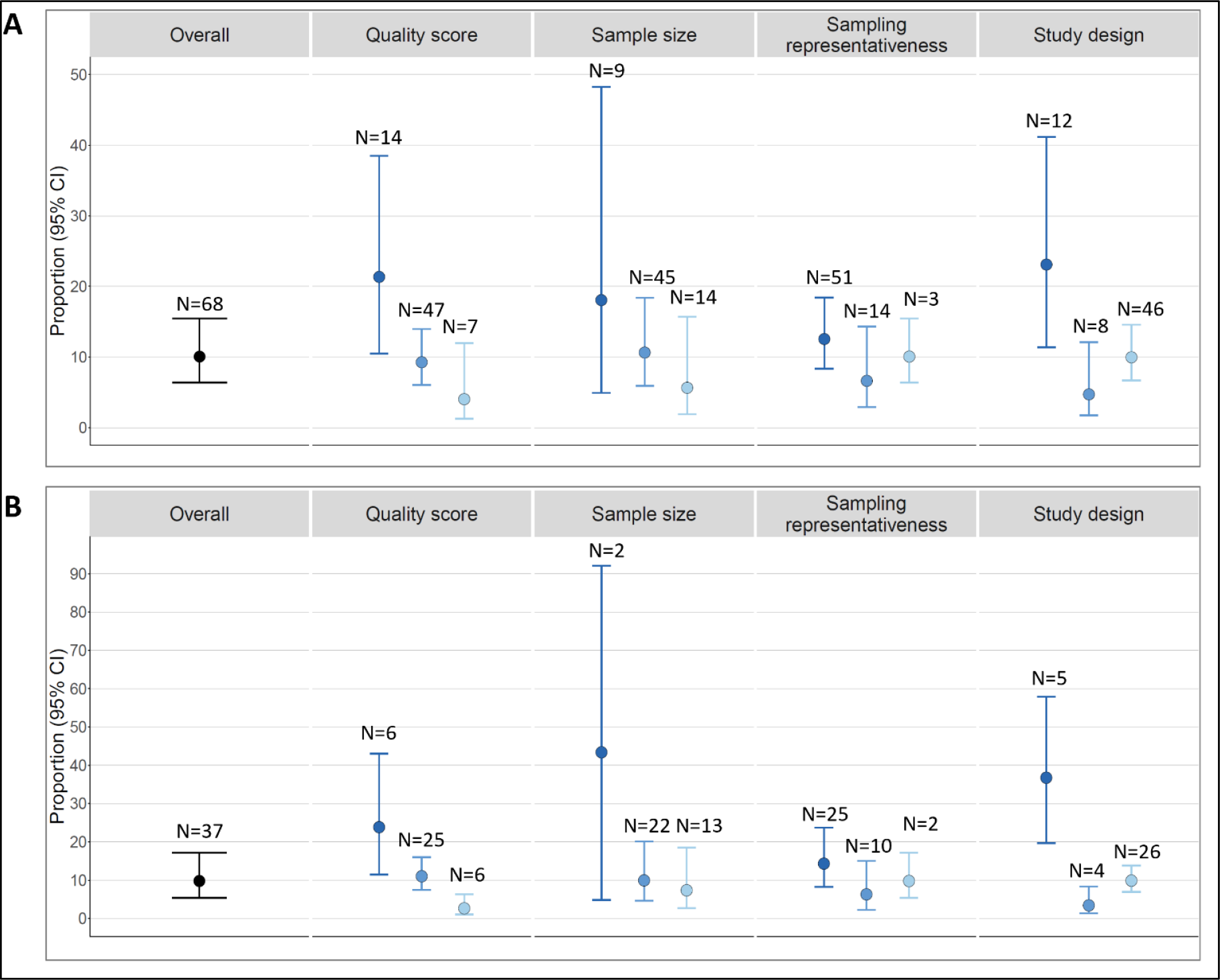
Meta-analysis results of top two most reported symptoms: chest pain and arrhythmia, using a random effects model*. * Figure 3A: Chest pain; 3B: Arrythmia. For stratified analysis of each characteristic from left to right: quality score (low, medium, high), sample size (30-99, 100-999, ≥ 1000), sampling representativeness (online survey and single hospital, multiple hospitals, national studies), and study design (cross-sectional, retrospective cohort, prospective cohort).

#### Publication bias

For the ten cardiac symptoms that underwent meta-analysis, we examined small-study effects or publication bias by visual inspection of the funnel plots and p-value of Egger’s regression tests for meta-analysis. Egger’s test suggested funnel plot asymmetry for arrhythmia (p<0.01) and hypertension (p<0.05) (**Figure S4**), indicating the presence of publication bias.

### Interpretation

Our systematic review and meta-analysis provide a comprehensive overview of evidence for the long-term cardiac symptoms following COVID-19. Up to January 2022, there were 101 studies that examined 49 different long-term cardiac symptoms. These studies varied substantially in design, and a small proportion of them (16%) were of high quality. Chest pain and arrhythmia were the two most studied long-term cardiac symptoms, and results from meta-analysis show that around 10% of participants had them. Meta-analysis identified high heterogeneity across studies for all cardiac symptoms and subgroup analyses showed systematic differences in reported proportions by selected study characteristics, including total quality assessment score, sample size, sampling representativeness, and study design.

It is evident that many COVID-19 survivors may experience chronic cardiovascular symptoms, even those without previous cardiovascular disease, comorbidities, and who have a baseline low risk of cardiovascular disease. To date, multiple reviews have examined the long-term cardiac symptoms following COVID-19 (**Table S6**).^10–12,14–16,18–20^ For example, several reviews quantified chest pain and arrhythmia as two of the most common long-term symptoms, with proportion estimates for chest pain ranging from 5.0% to 16.0% and proportion estimates for arrhythmia ranging from 9.7% to 11%.^10,13,16,18^ Our proportion estimates of these two symptoms broadly agree with findings from these previous reviews. A probable source of differences in estimates across these systemic reviews is that they used different inclusion criteria to select studies.

Although pathophysiological mechanisms underlying COVID-19 cardiac symptoms remain unclear, studies suggest that the chronic inflammatory response may be hyperactivated by persistent viral reservoirs in the initial acute phase, which may lead to post-acute COVID-19 cardiovascular sequelae.^19,20^ Studies have shown that over 20% of patients with acute COVID-19 had evidence of cardiac injury, even if they did not have underlying cardiovascular diseases or pre-existing comorbidities.^24–27^ It is hypothesized that viral invasion through binding angiotensin-converting enzyme-2 (ACE-2) causes a cytokine storm and triggers systemic hyper-inflammation, which can affect multiple organ systems and induce cardiac injury as one of the severe complications.^13,14^ Persistent chest pain and arrythmia may be indicative of underlying cardiac abnormalities and damage resulting from the systematic hyper-inflammation and/or viral yocarditis affecting the cardiac conduction system. It is critical for clinicians to thoroughly examine patients with long-term cardiac symptoms following COVID-19, especially for survivors with pre-existing cardiac conditions and other high-risk comorbidities.

We found that studies with a low-quality assessment score, small sample size, non-systematic sampling methods, and cross-sectional designs were more likely to report high proportions of long-term cardiac symptoms. For example, the pooled proportion of chest pain among low-quality score studies (21%) was over four times higher than that among high quality score studies (4%). Such patterns were also observed for arrhythmia and other long-term cardiac symptoms. This strong relationship between the proportion of long-term cardiac symptoms following COVID-19 and study quality and design characteristics should be considered when interpreting existing studies.

In our quality assessment, we determined that around 16% of included studies were of high quality^6,7,28–41^ and the remaining 84% were of medium or low quality. The small number of high-quality studies demonstrates the urgent need to improve the quality of studies investigating the long-term cardiac symptoms following COVID-19. A key feature among most included studies of medium or low quality was that they were predominantly based on clinical or hospital samples. These studies usually had small sample sizes and had no or only one point of follow-up. While relatively small clinical or hospital-based studies, which are often easier to conduct in shorter time periods, can be useful to establish preliminary evidence of an association especially in an emergent situation such as a pandemic, they often lack population representativeness and statistical power to make broader conclusions about the hypothesized relationships. Furthermore, cross-sectional studies cannot establish the temporality required to infer any causal relationship and also prohibit examinations of changes in symptoms over time.

Based on the above findings, we formulated recommendations for the design and analysis of future studies on long-term cardiac symptoms of COVID-19 (**Table 2**). Many studies adopted convenience sampling schemes, which hinders the interpretability and generalizability of their findings. Therefore, it is important to conduct systematic sampling, which can facilitate a continuing and meaningful exploration of data collected and underpin clinical research. The majority of included studies only assessed long-term symptoms at a single time point. It is therefore challenging to examine how the long-term symptoms may change over time. Many studies did not distinguish between long-term symptoms following COVID and pre-COVID symptoms at baseline level.

**Table 2.** Recommendation list for future long COVID-19 studies.

| **Stage** | **Recommendation** | **Rationale** |
| --- | --- | --- |
| **Sample and survey** | Conduct systematic sampling and oversample participants with major potential risk factors for long-term symptoms | Relate study population to a well-defined source population and increase statistical power to conduct hypothesis test |
|  | Collect information on pre-COVID symptoms and conditions | Allow to distinguish long-term symptoms and pre-COVID symptoms or the population's baseline level |
|  | Conduct multiple follow-up activities to examine changes in long-term symptoms over time | Establish temporality and compare the rate of symptom development between comparison groups |
| **Design and analysis** | Apply appropriate analytical methods, including confounding adjustment for major demographic and socioeconomic factors, pre-existing conditions, and comorbidities | Control for major factors related to long-term symptoms |
|  | Use causal knowledge and graphs to guide covariate adjustments and provide a rationale for a priori selection of potential confounders | Reduce confounding and decrease the risk of including variables that could increase bias |
|  | Present information on number of cases and population at risk by COVID severity status and other important risk factors, and results of both crude and adjusted models | Allow for a close examination on main study results and the uncertainty that may result from small numbers |

Our study has several limitations. First, studies included in our systematic review and meta-analysis are highly heterogenous. We therefore performed subgroup analyses by multiple characteristics, and we believe that existing heterogeneity across studies makes it difficult to generalize our results to the general population. Second, many reported outcomes had a small sample size, which increases the possibility for publication bias. Third, we were unable to stratify our meta-analysis by length of follow-up because of widely varying follow-up times across studies. Finally, we were not able to stratify our analyses by prior comorbidities, previous cardiovascular disease, history of treatment or medication use for cardiac symptoms, or COVID-19 vaccination status, mainly because this information was underreported, especially in the articles published in the early phase of the pandemic. As the pandemic continues to progress, the virus mutates, treatment strategies for acute and long COVID-19 evolve, and as the uptake of vaccine increases, it is possible that the epidemiology of long-term cardiovascular manifestations in COVID-19 might change over time.

Our systematic review and meta-analysis also have multiple strengths. First, to our knowledge, this is the first systematic review focusing on long-term cardiac symptoms following COVID-19 that included preprints and articles published in all languages and in the global network. Second, in an effort to ensure our results were current, we updated our search to capture articles published from the early phases of the pandemic to the most recently published studies. Third, we assessed the quality of included articles from the perspective of study design and epidemiologic principles and provided detailed recommendations on future long-COVID epidemiologic research. Fourth, we performed meta-analysis and subgroup analyses to examine patterns of reported findings.

In conclusion, this systematic review and meta-analysis synthesized the evidence on the long-term cardiac symptoms following COVID-19. We found there were diverse manifestations of cardiac symptoms, and many can last for months and even over a year. There is substantial heterogeneity in terms of study design and systematic differences in reported proportion of symptoms by study characteristics. Specifically, we found that studies with low-quality, small sample size, unsystematic sampling method, or cross-sectional design were most likely to report high estimates of symptoms among individuals survived COVID-19. We believe that a deeper understanding of long COVID is currently prevented by the limitations of the published literature. Our study underscores the need to conduct high-quality studies on long COVID and the importance of long-term cardiac surveillance of COVID-19 survivors.

## Data Availability

All data relevant to the study are included in the article or included as supplementary materials.

## Data availability statement

All data relevant to the study are included in the article or included as supplementary materials.

## Author contributions

All authors contributed to study conceptualization, data analysis, writing, interpretation of results and reviewing the final draft.

