## Supplementary Figures for "Long-term cardiac symptoms following COVID-19: a systematic review and meta-analysis"

**Figure S1. Quality assessment by studies on long-term cardiac symptoms of COVID-19 infection**

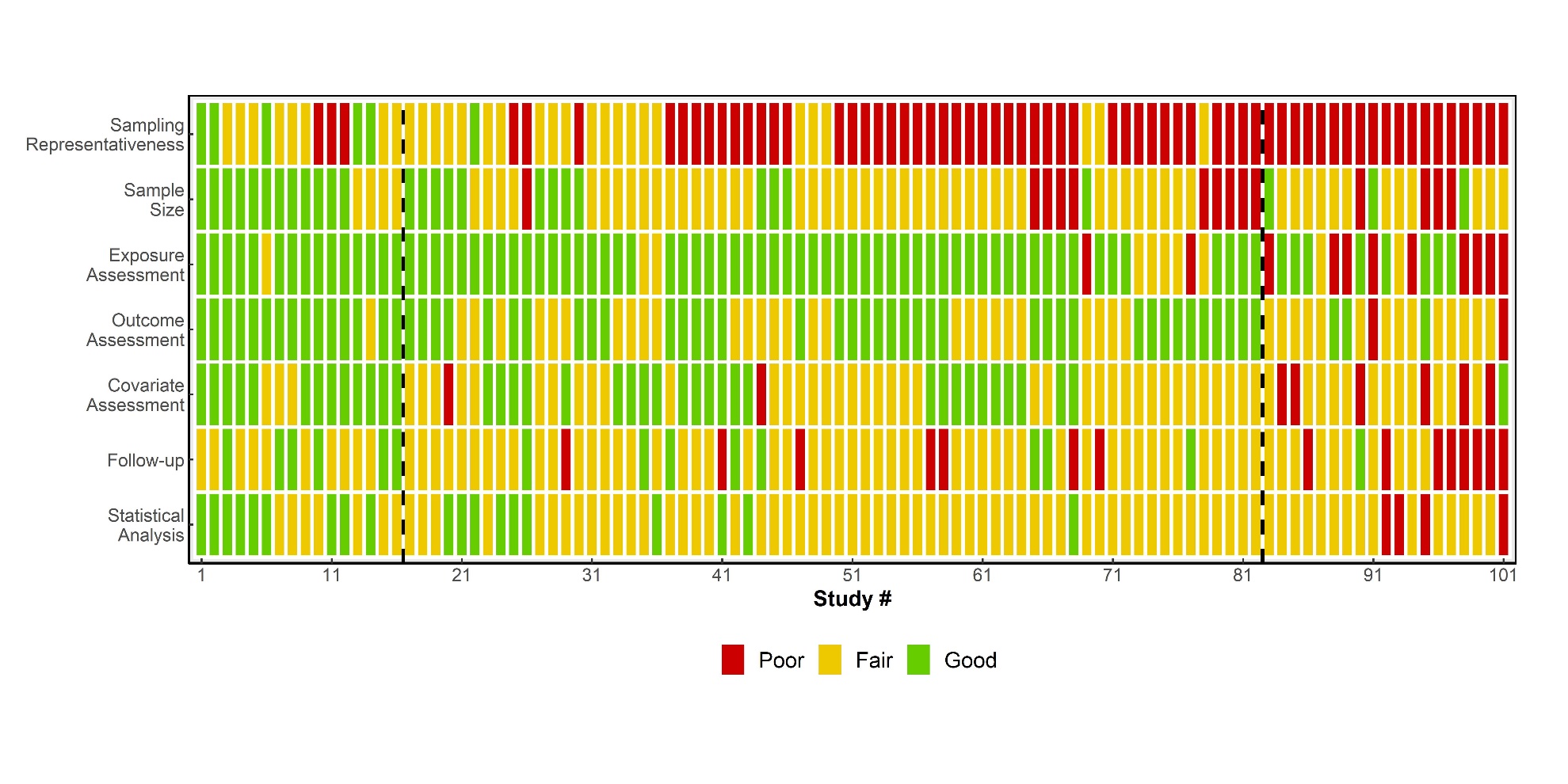

**Figure S2.** Forest plots of the proportions of the top 10 most reported long-term cardiac symptoms among COVID-19 survivors.

**Chest pain**

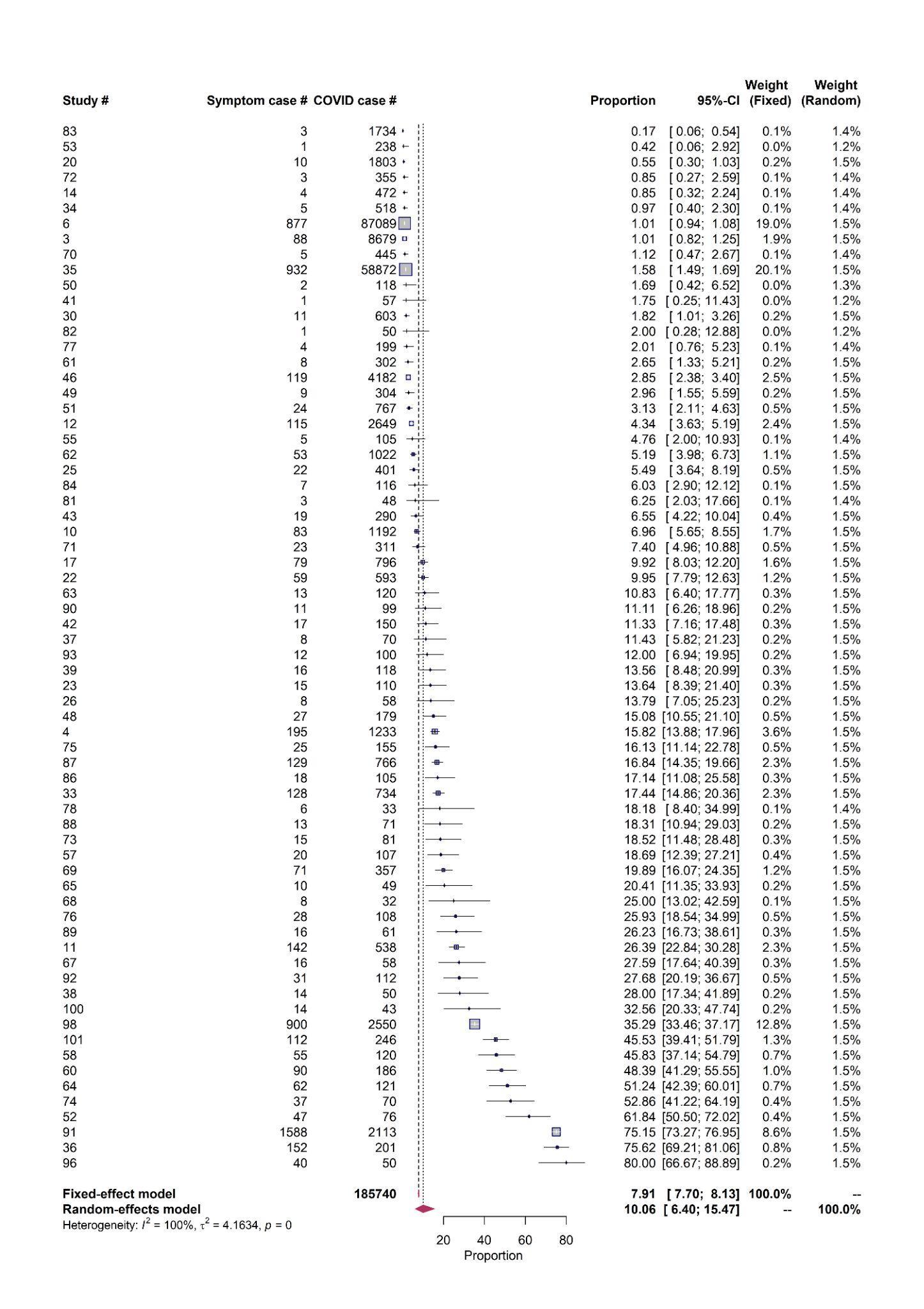

**Arrhythmia**

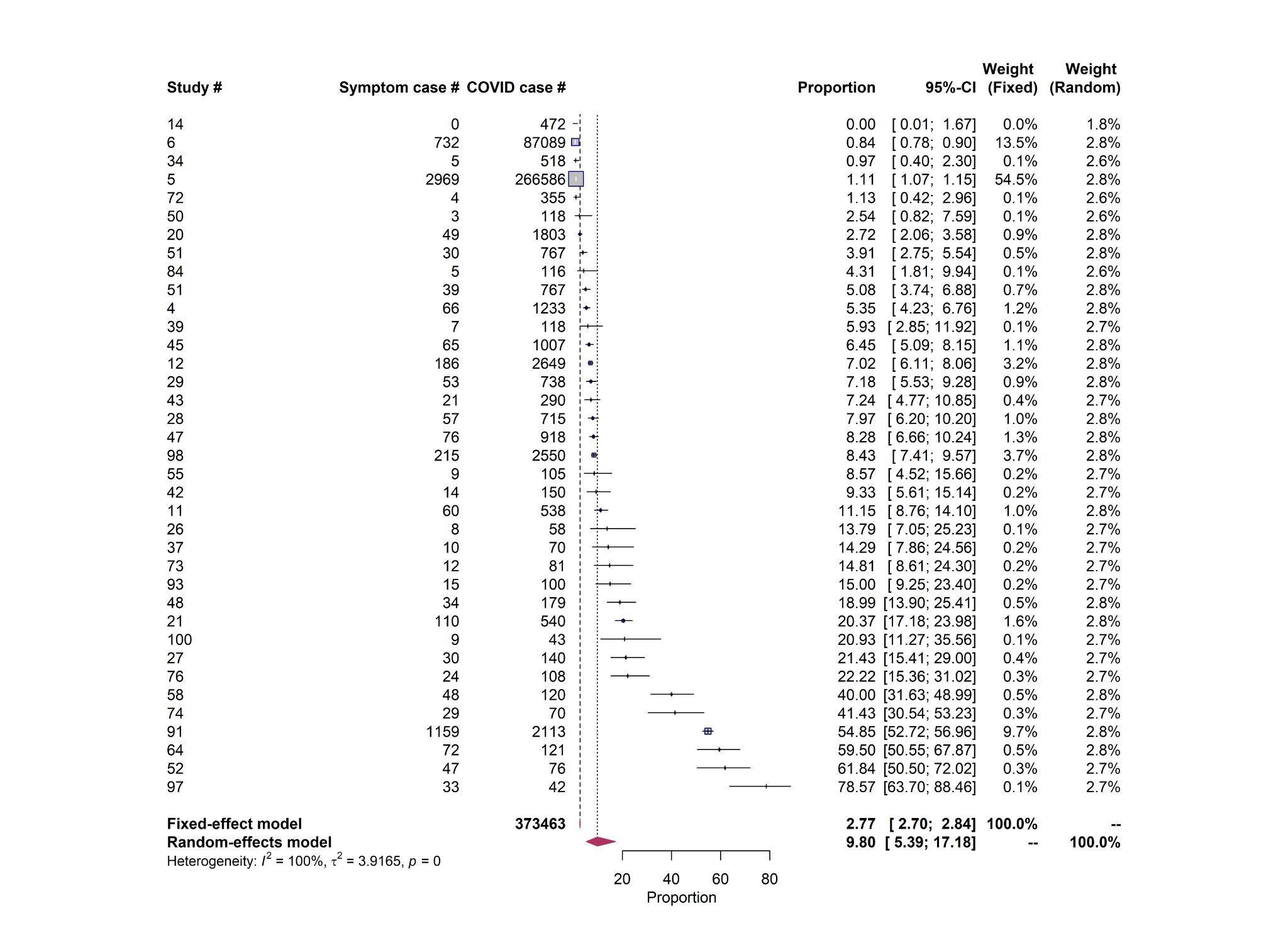

**Hypertension**

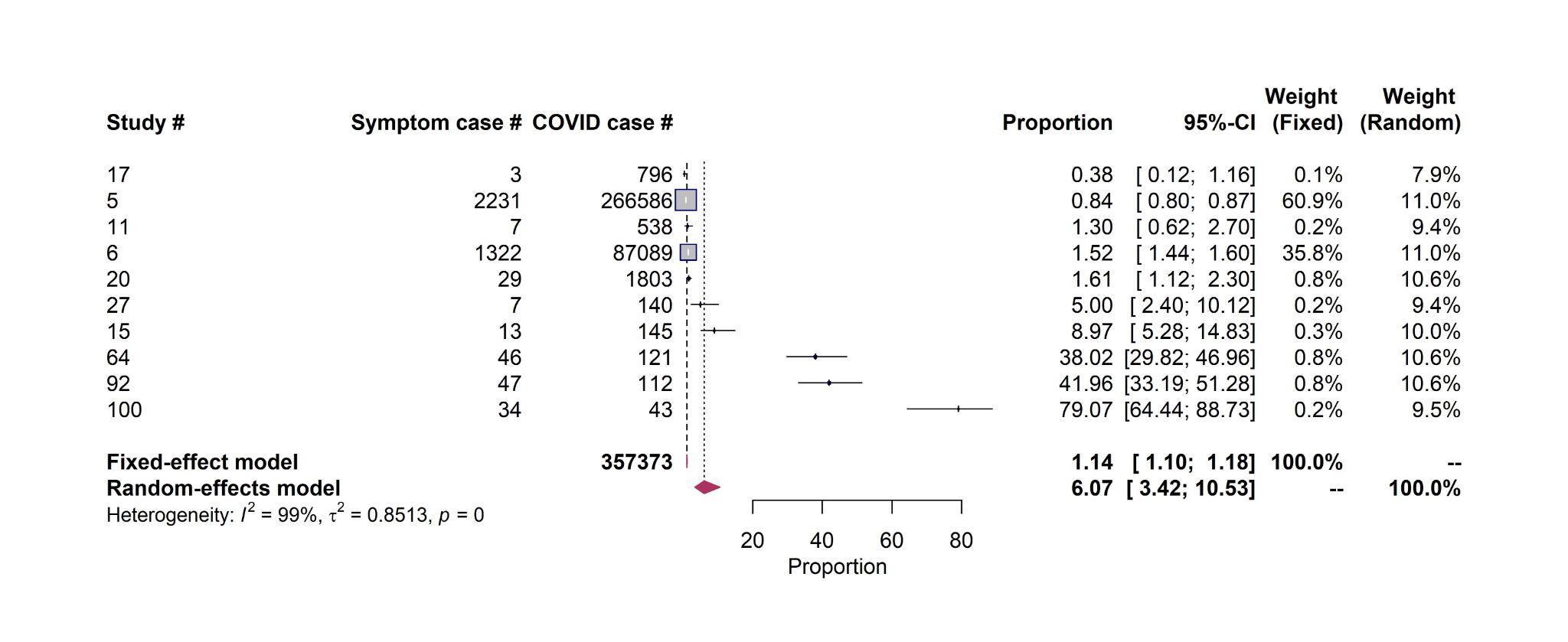

**Cardiac abnormalities**

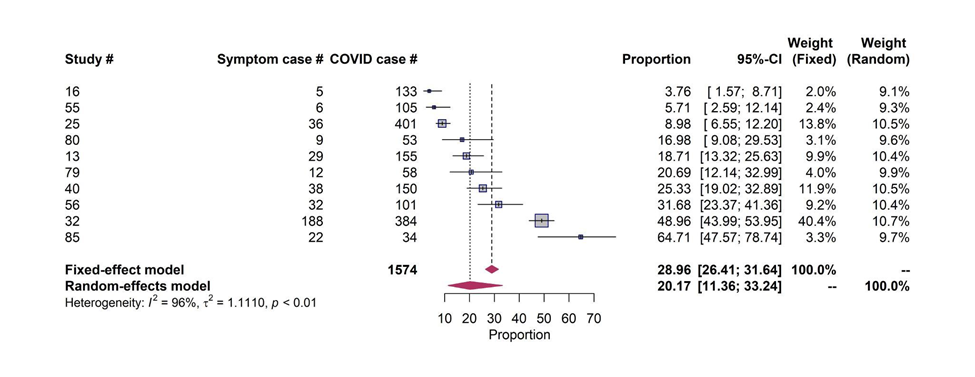

**Myocardial injury**
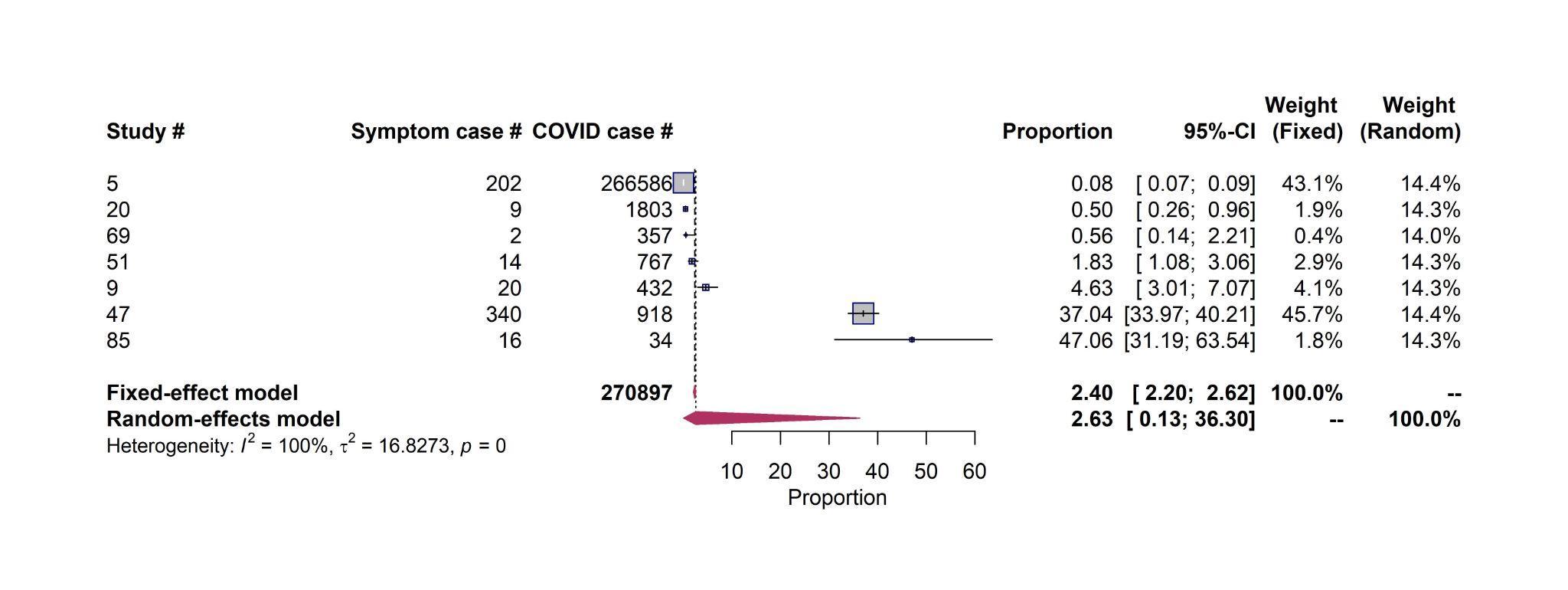

**Thromboembolism**

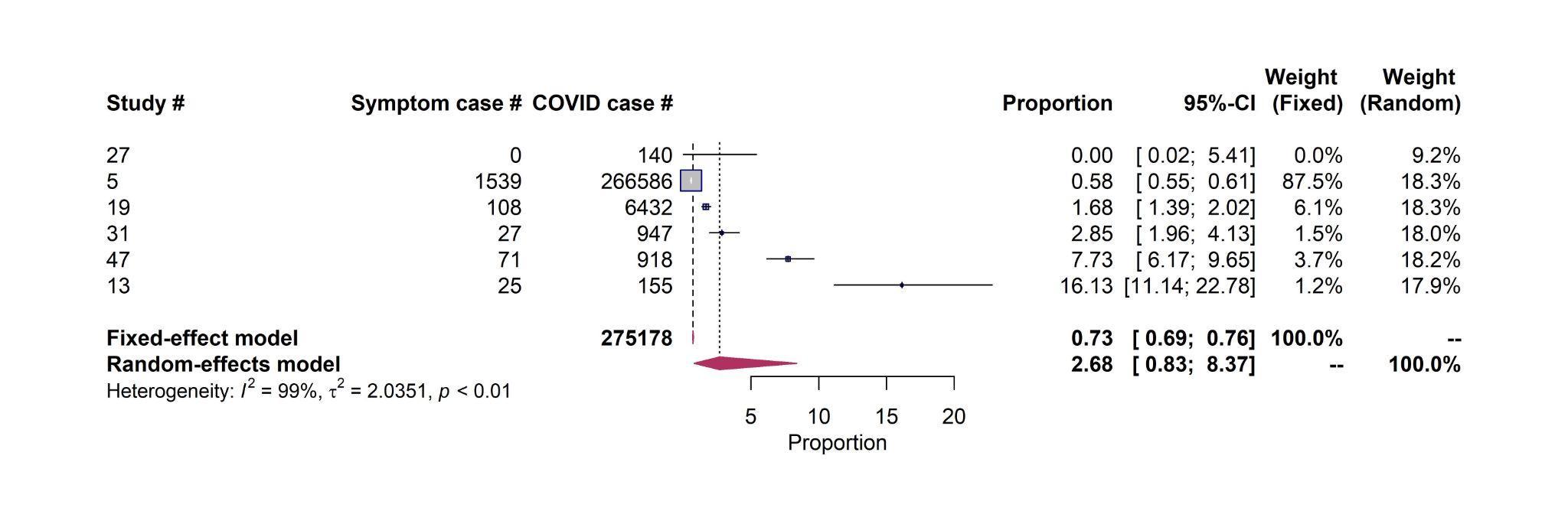

**Stroke**

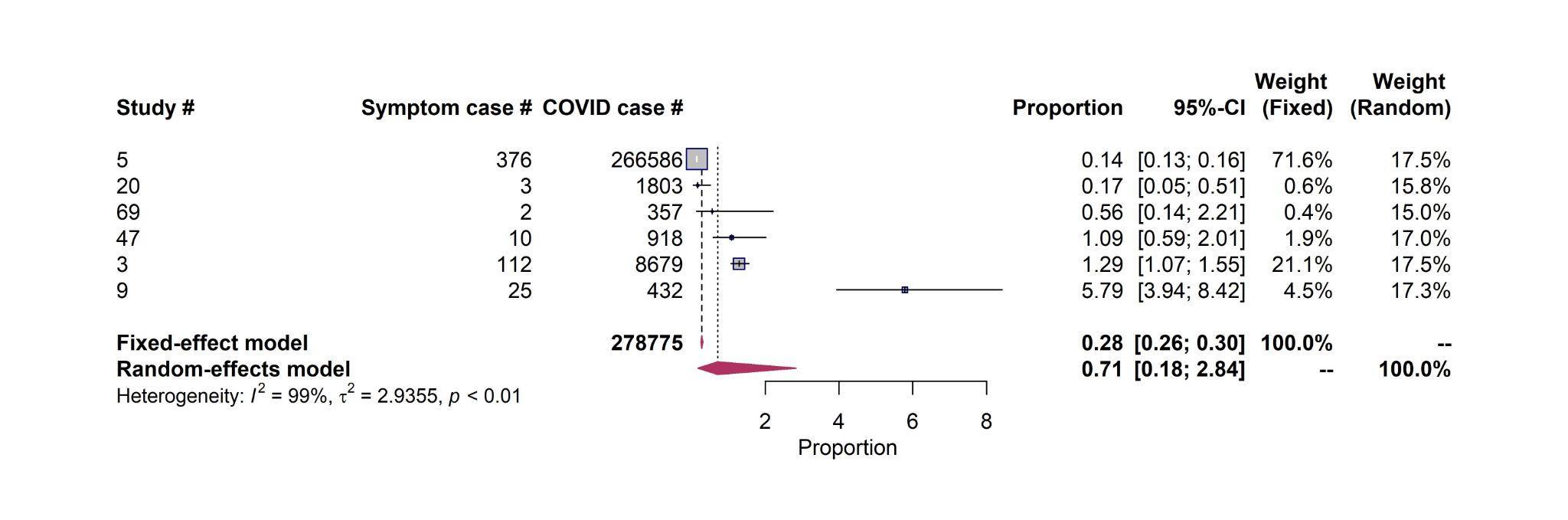

**Heart failure**

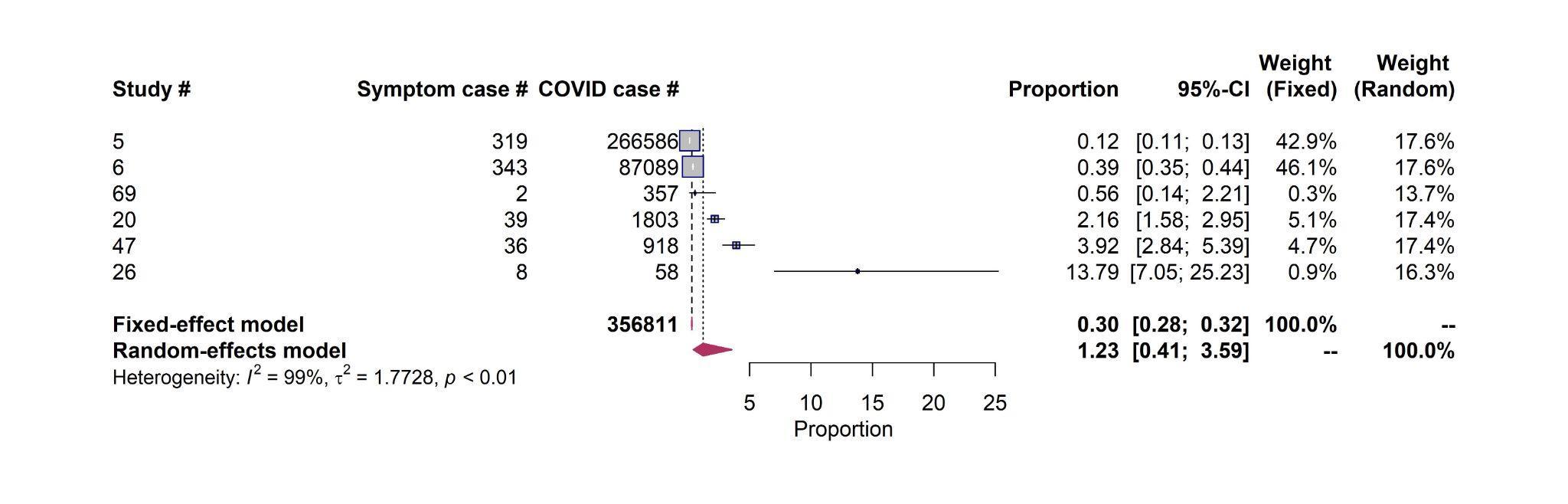

**Coronary disease**
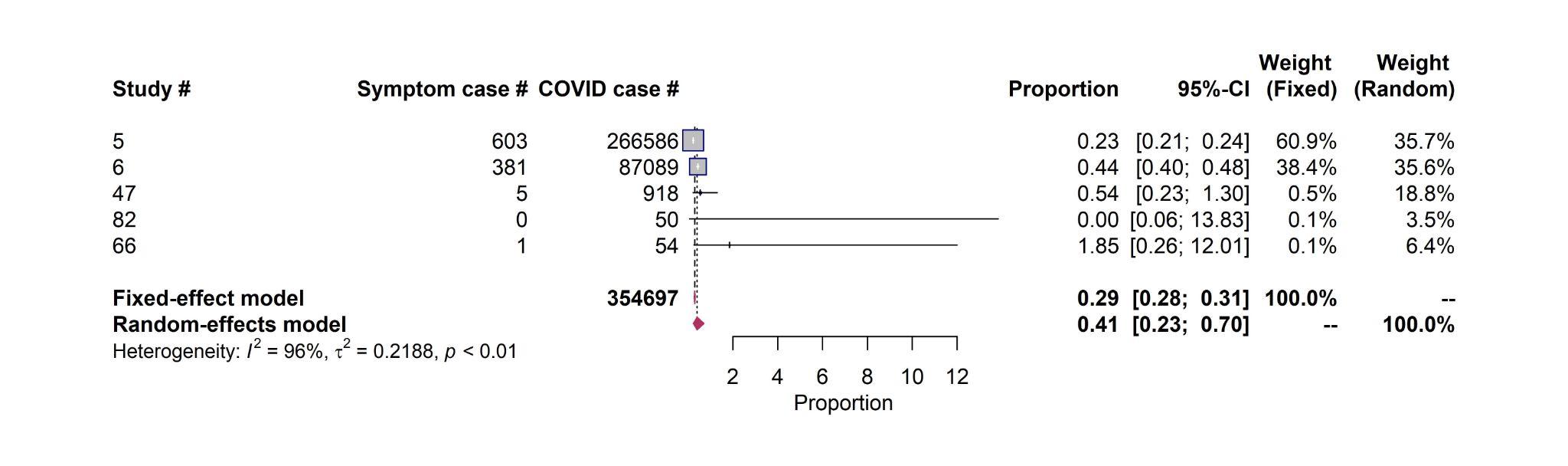

**Myocarditis**

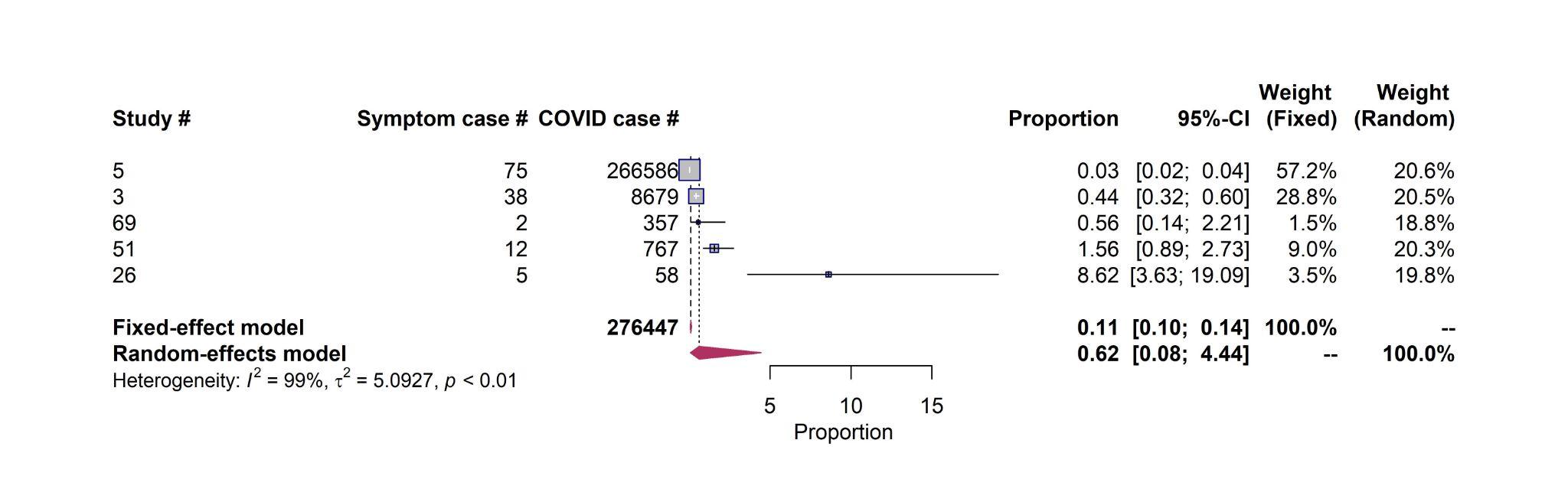

**Figure S3.** Forest plots of the proportions of the top 10 most reported long-term cardiac symptoms among COVID-19 survivors, stratified by study characteristics.

**Chest pain – Quality score**

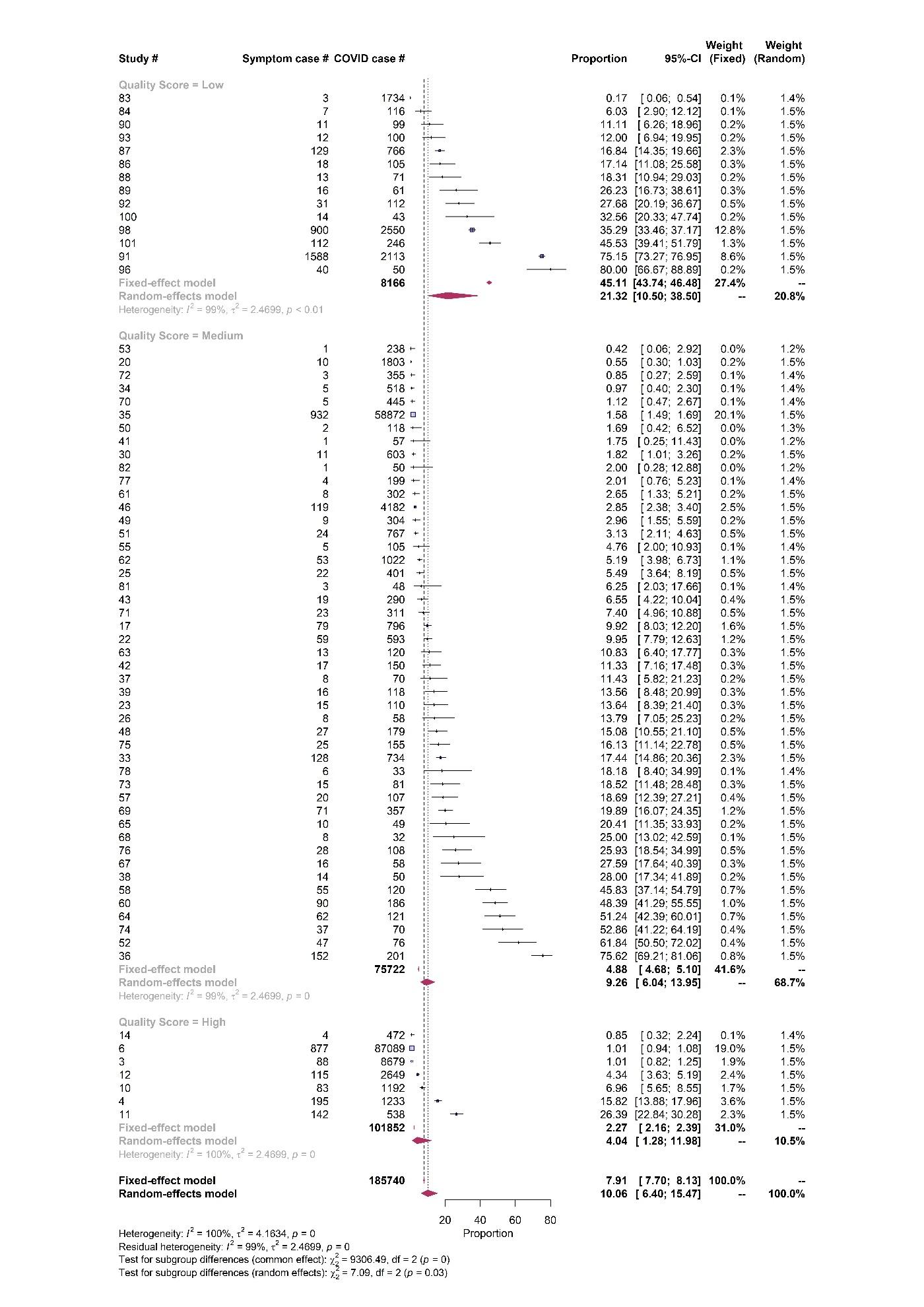

**Chest pain – Sample size**

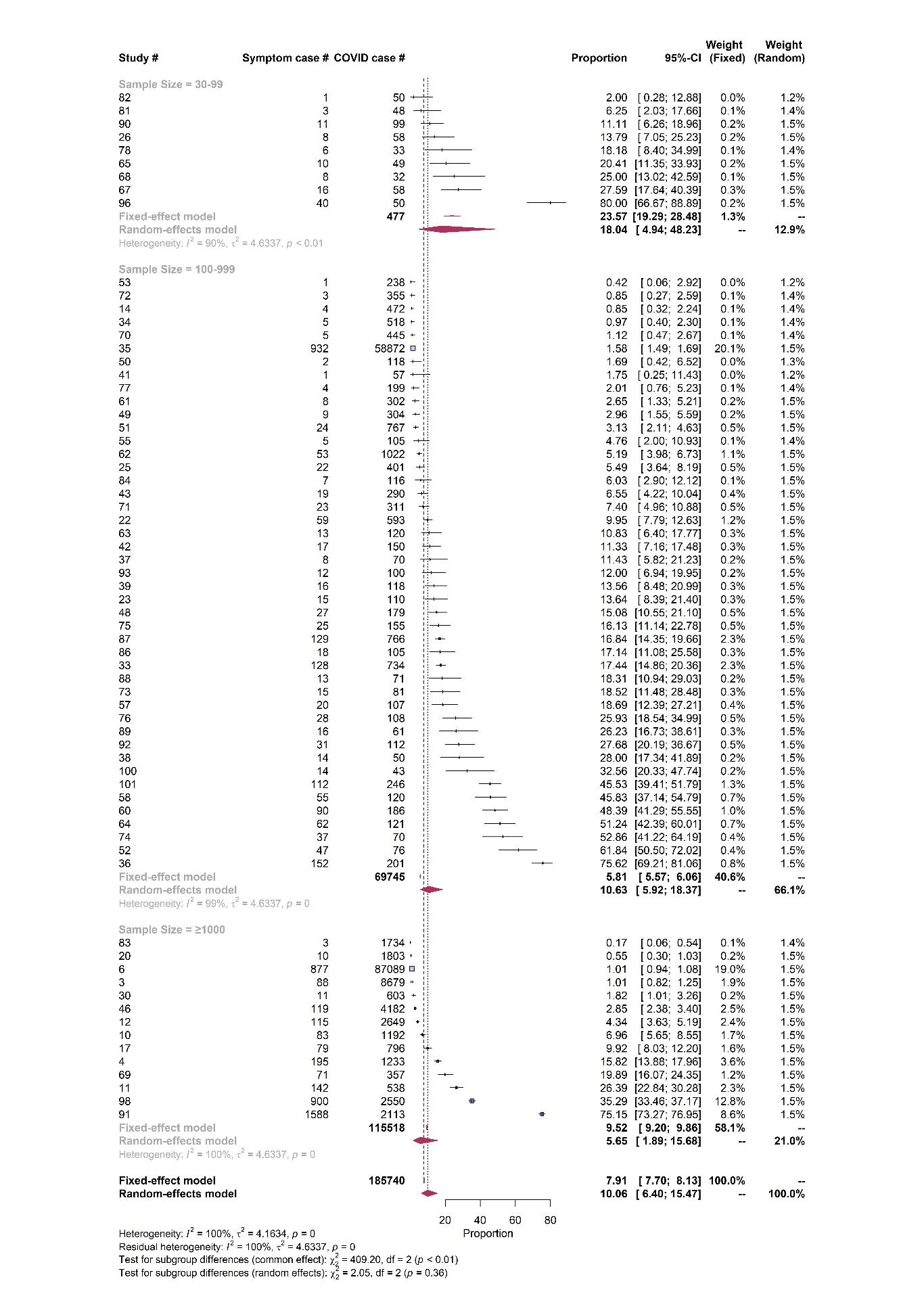

**Chest pain – Sampling representativeness**

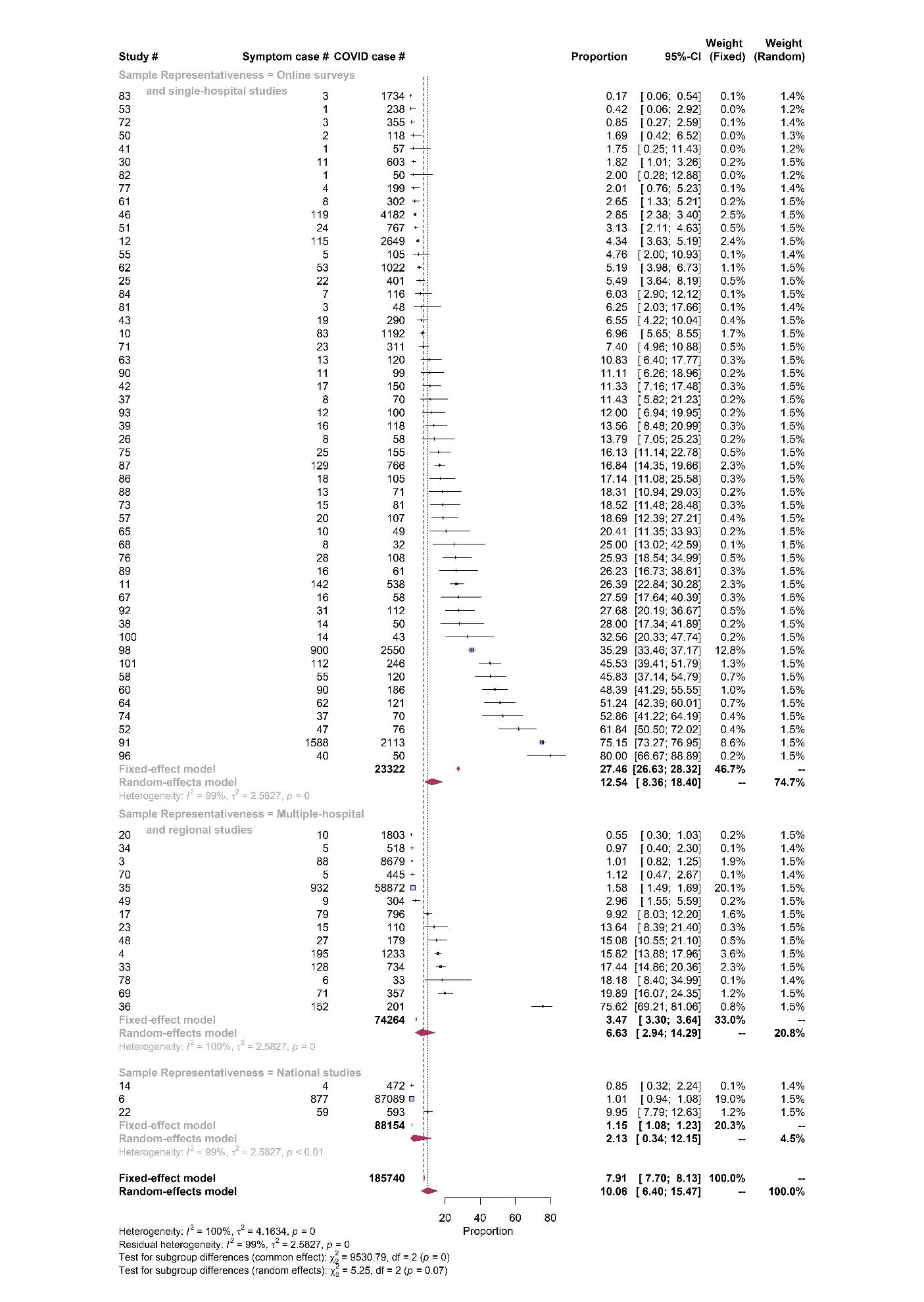

**Chest pain – Study design**

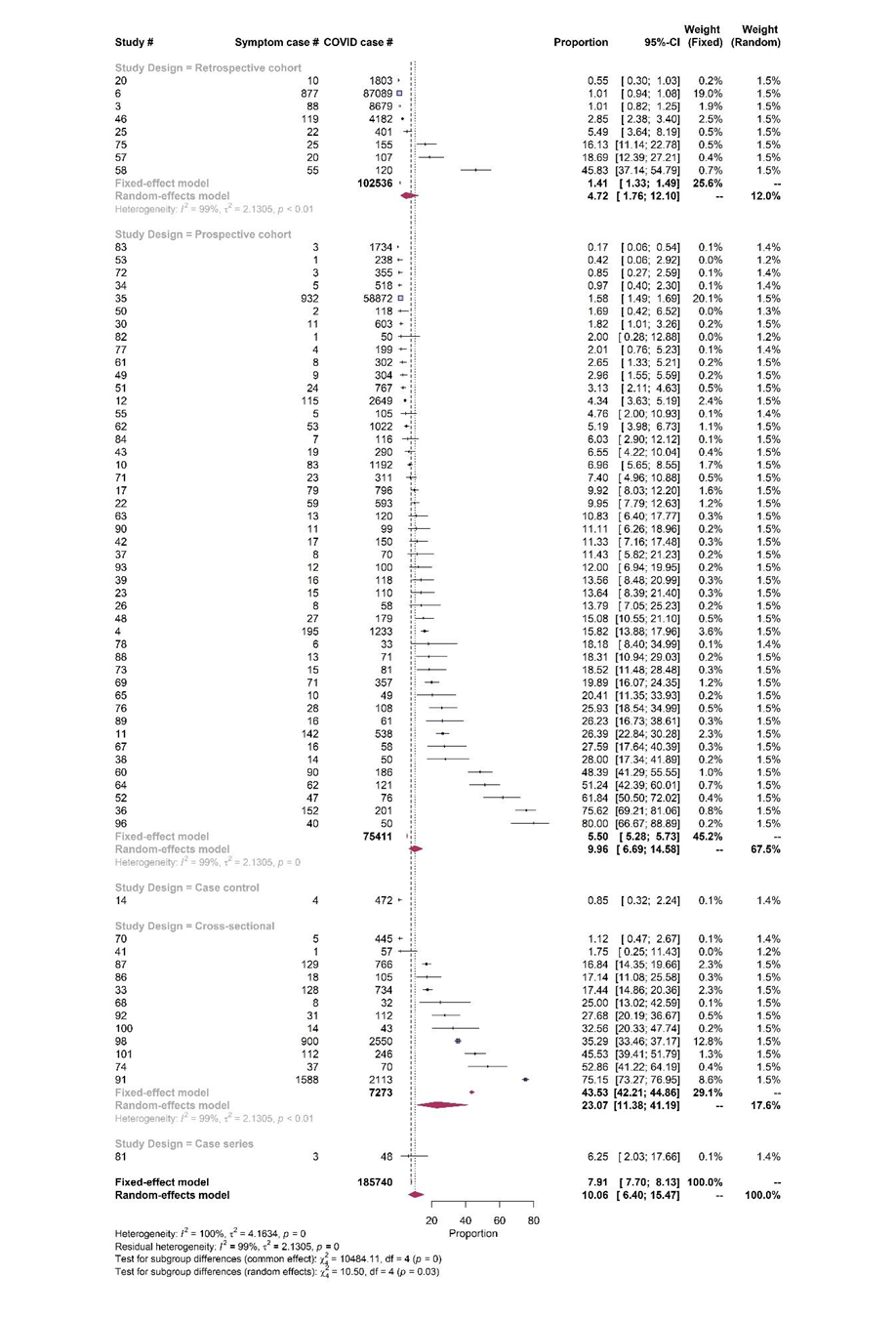

**Arrhythmia – Quality score**

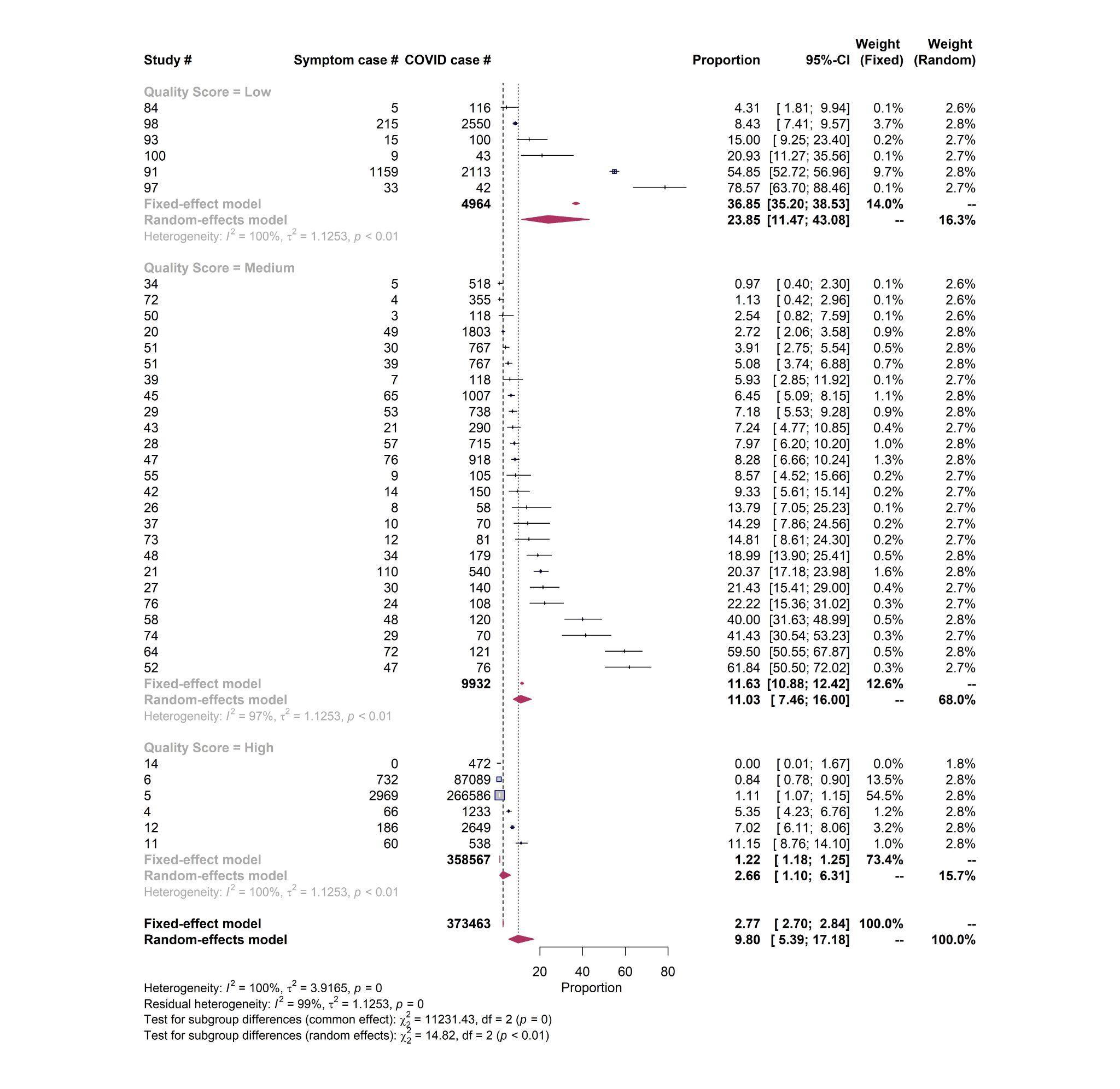

**Arrhythmia – Sample size**

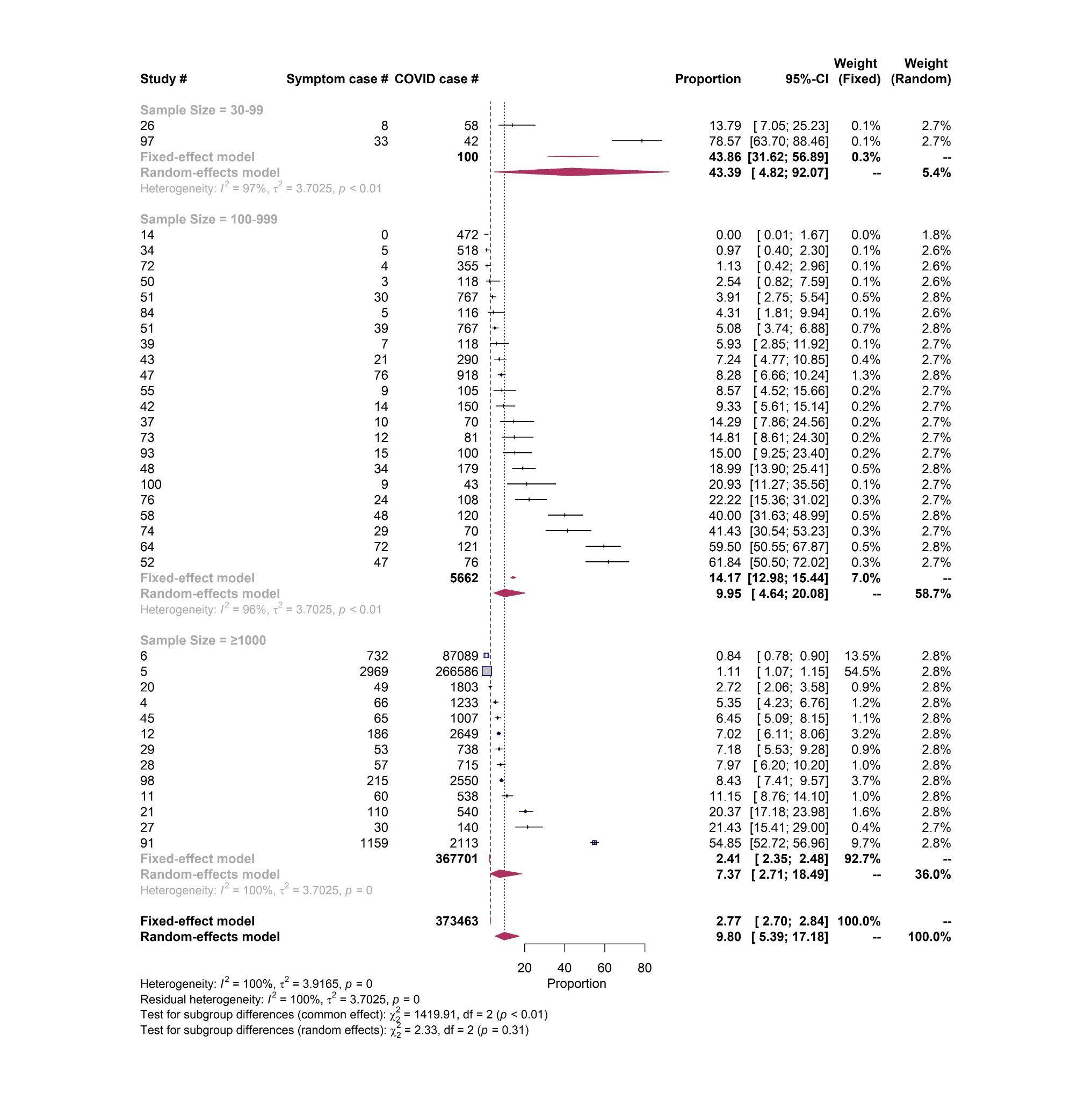

**Arrhythmia – Sampling representativeness**

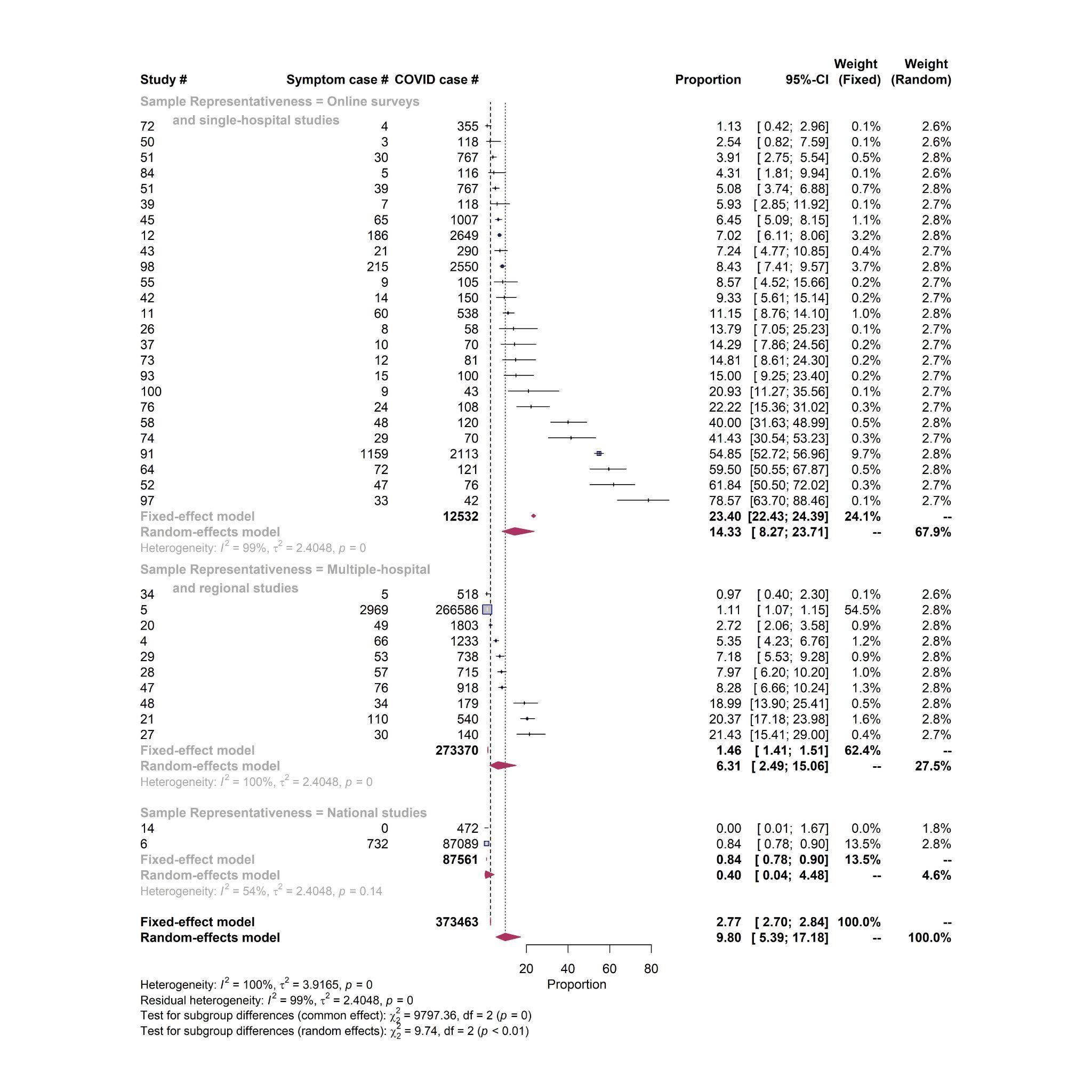

**Arrhythmia – Study design**

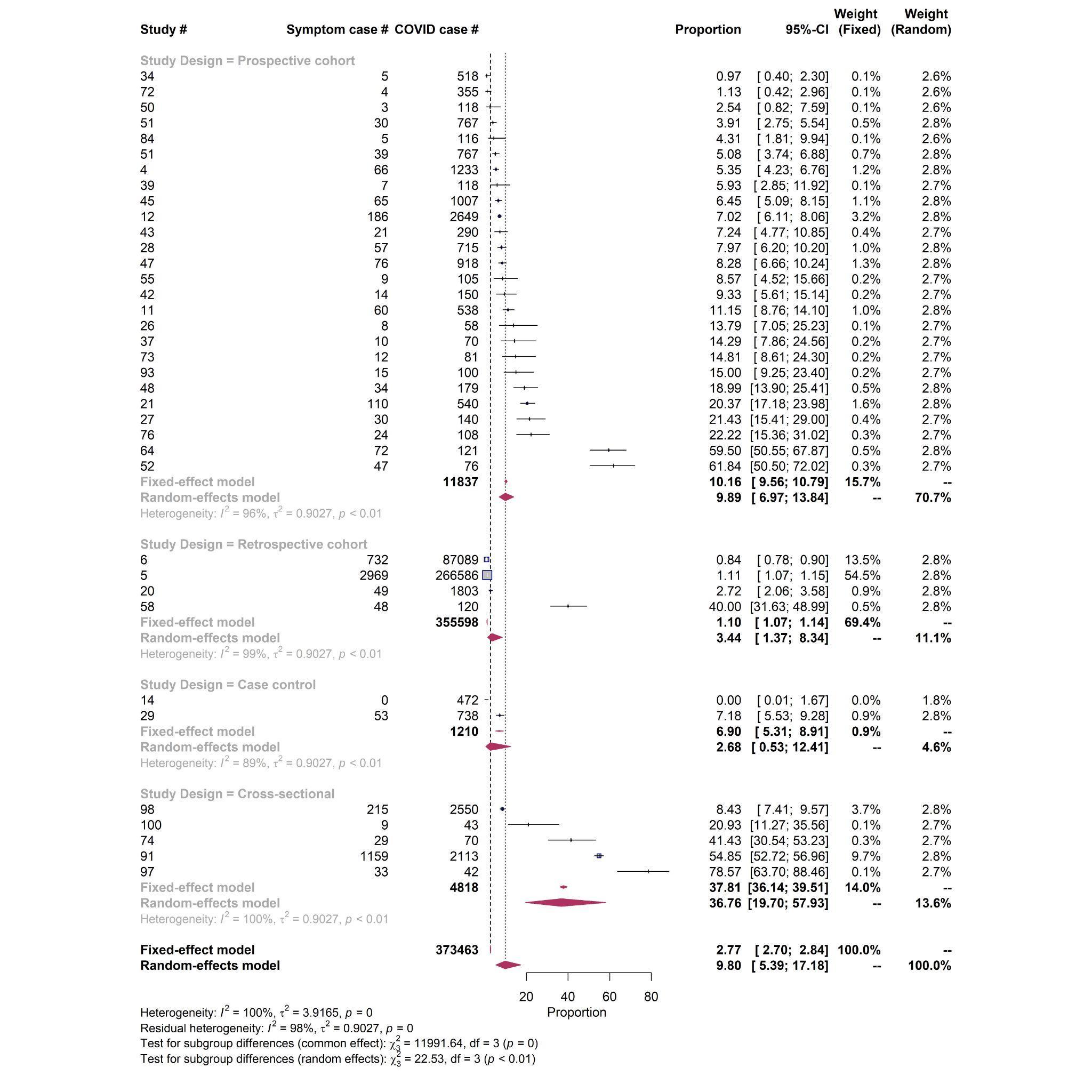

**Hypertension – Quality score**
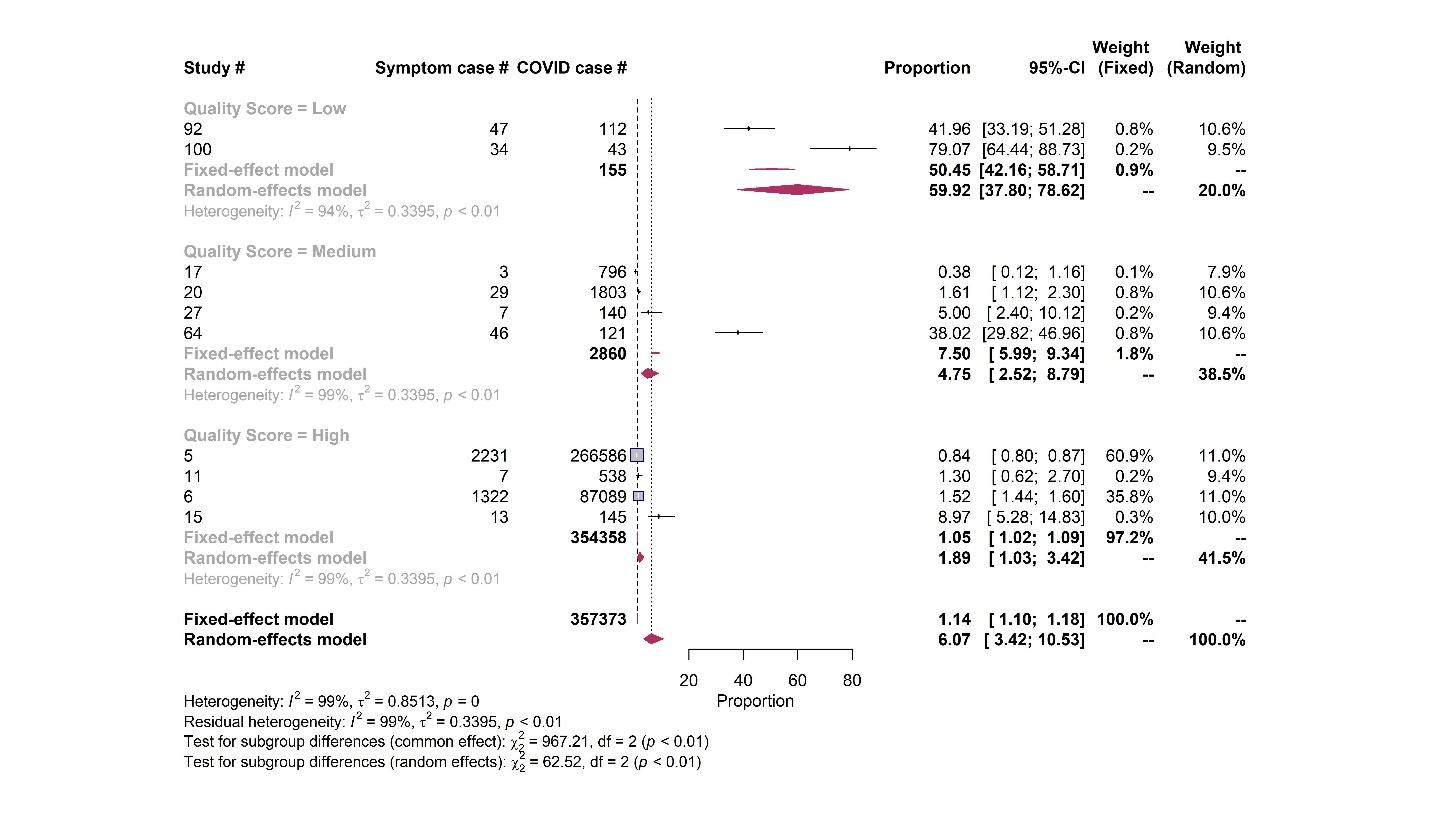

**Hypertension – Sample size**

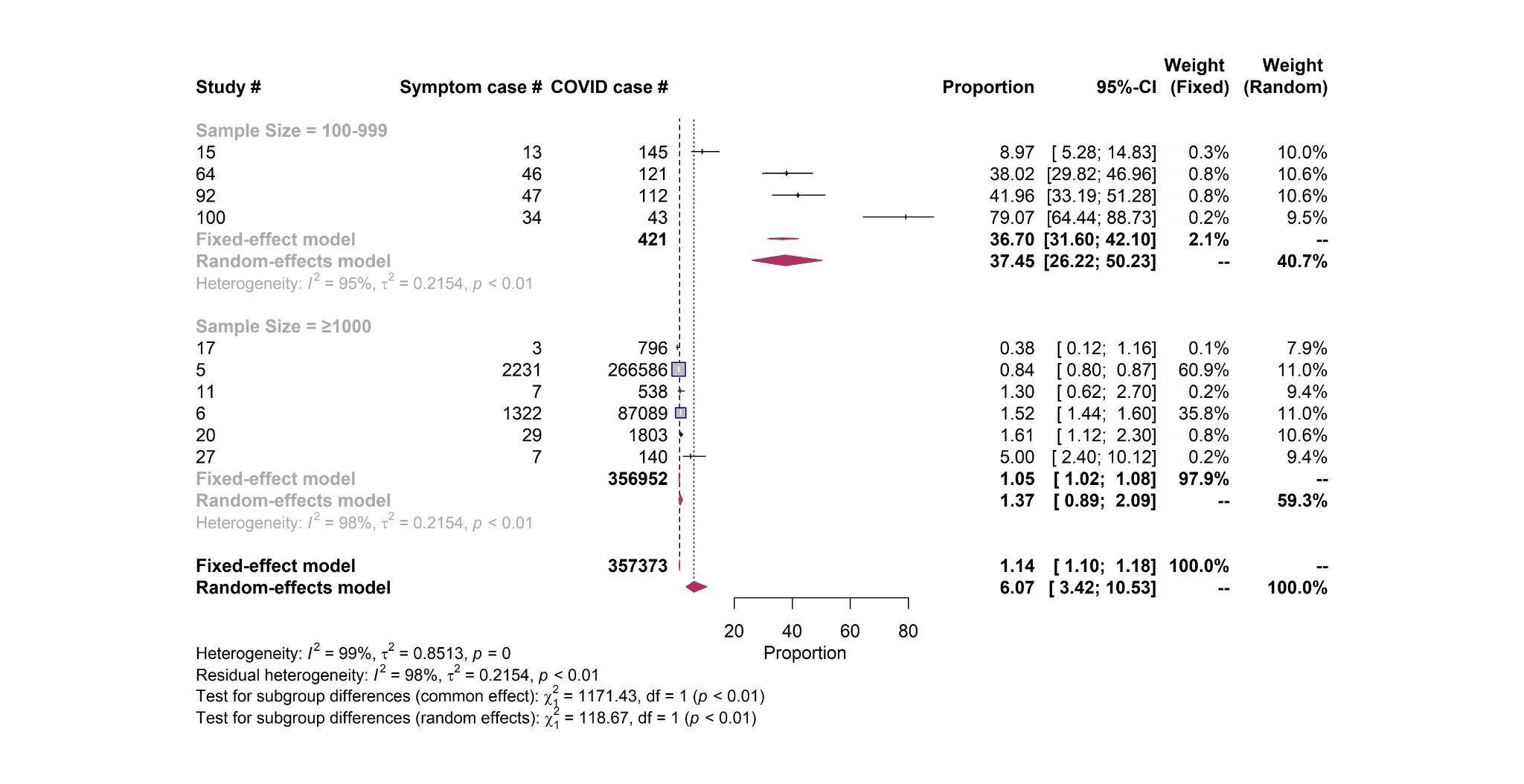

**Hypertension – Sampling representativeness**

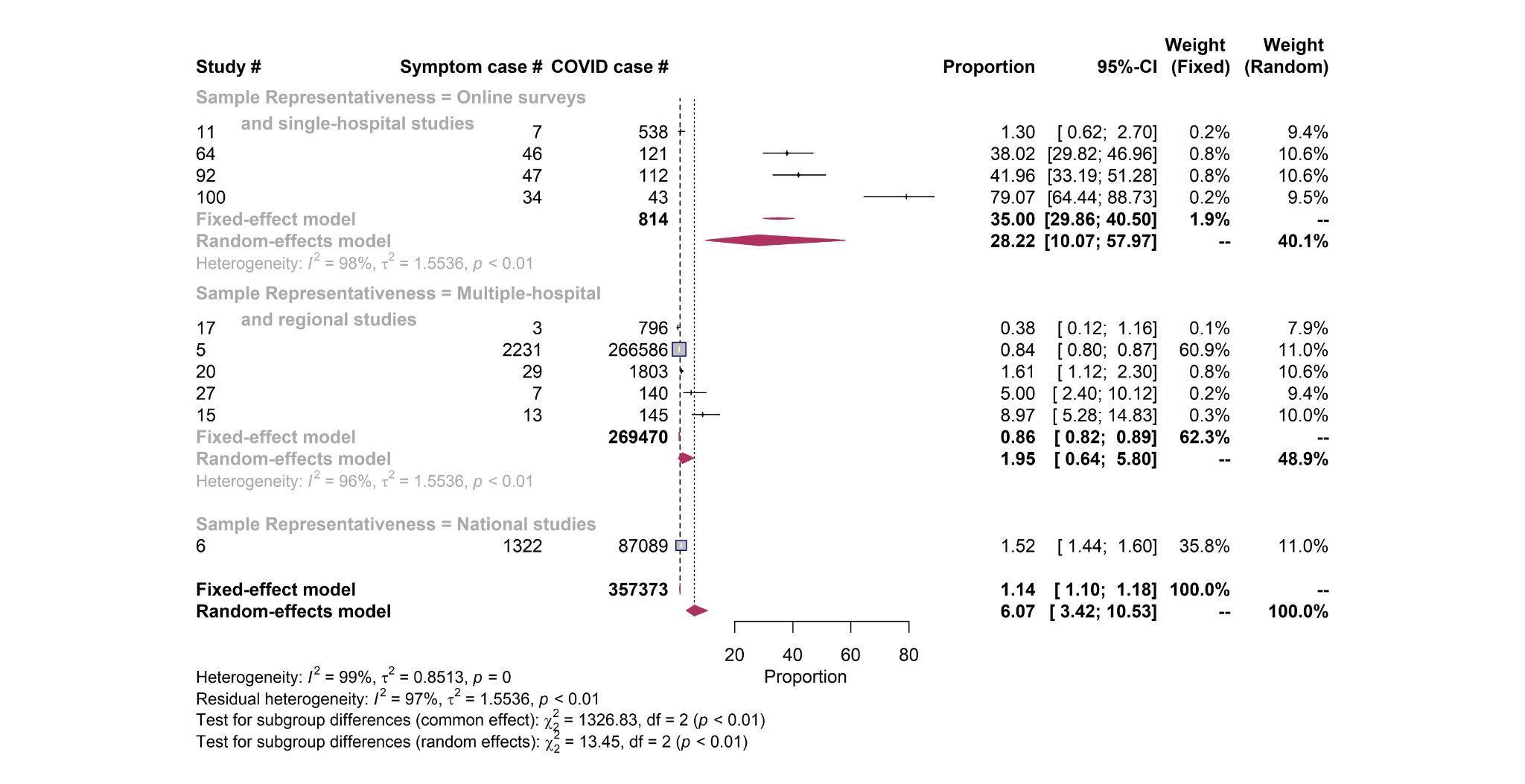

**Hypertension – Study design**

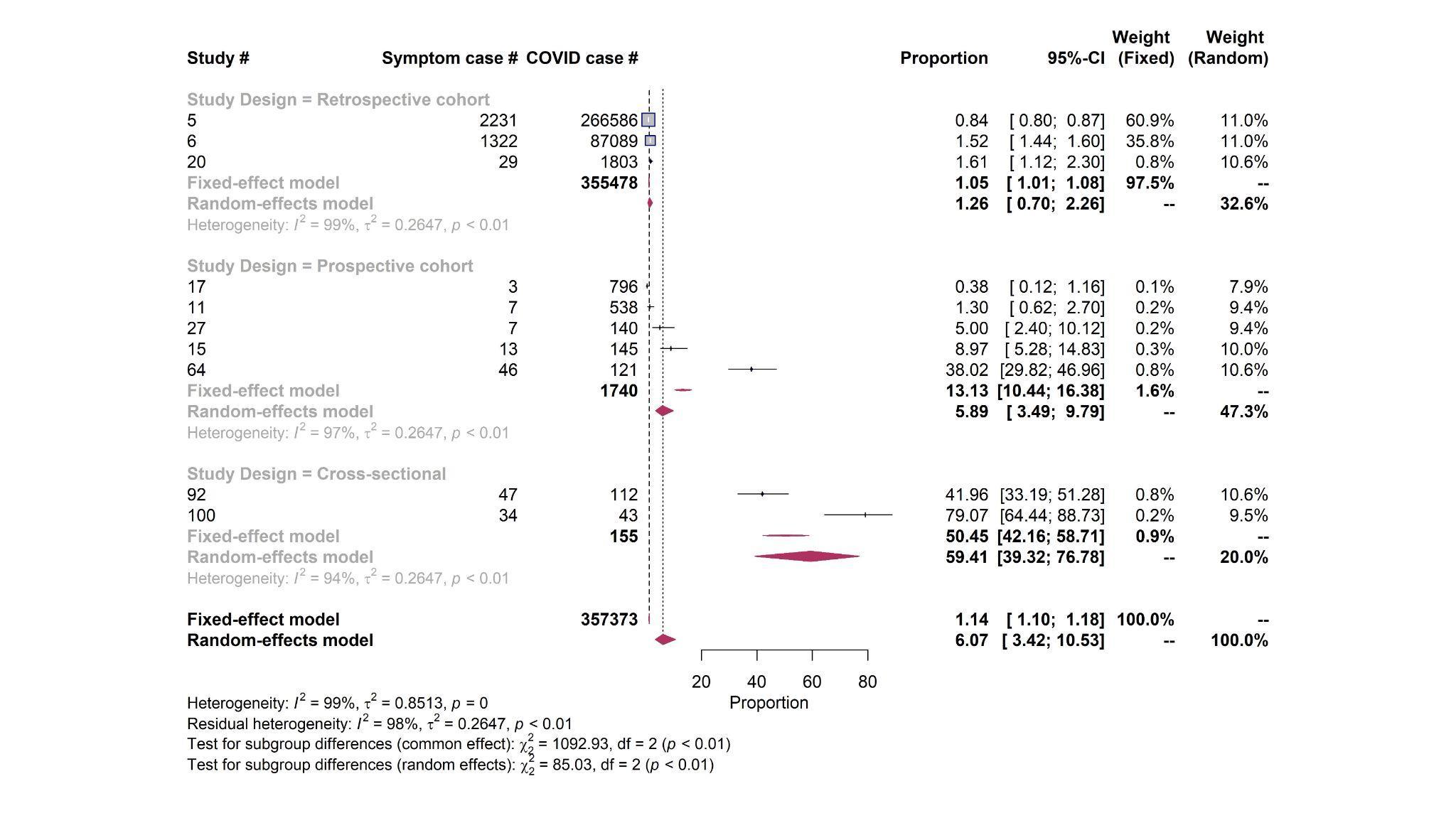

**Cardiac abnormalities – Quality score**

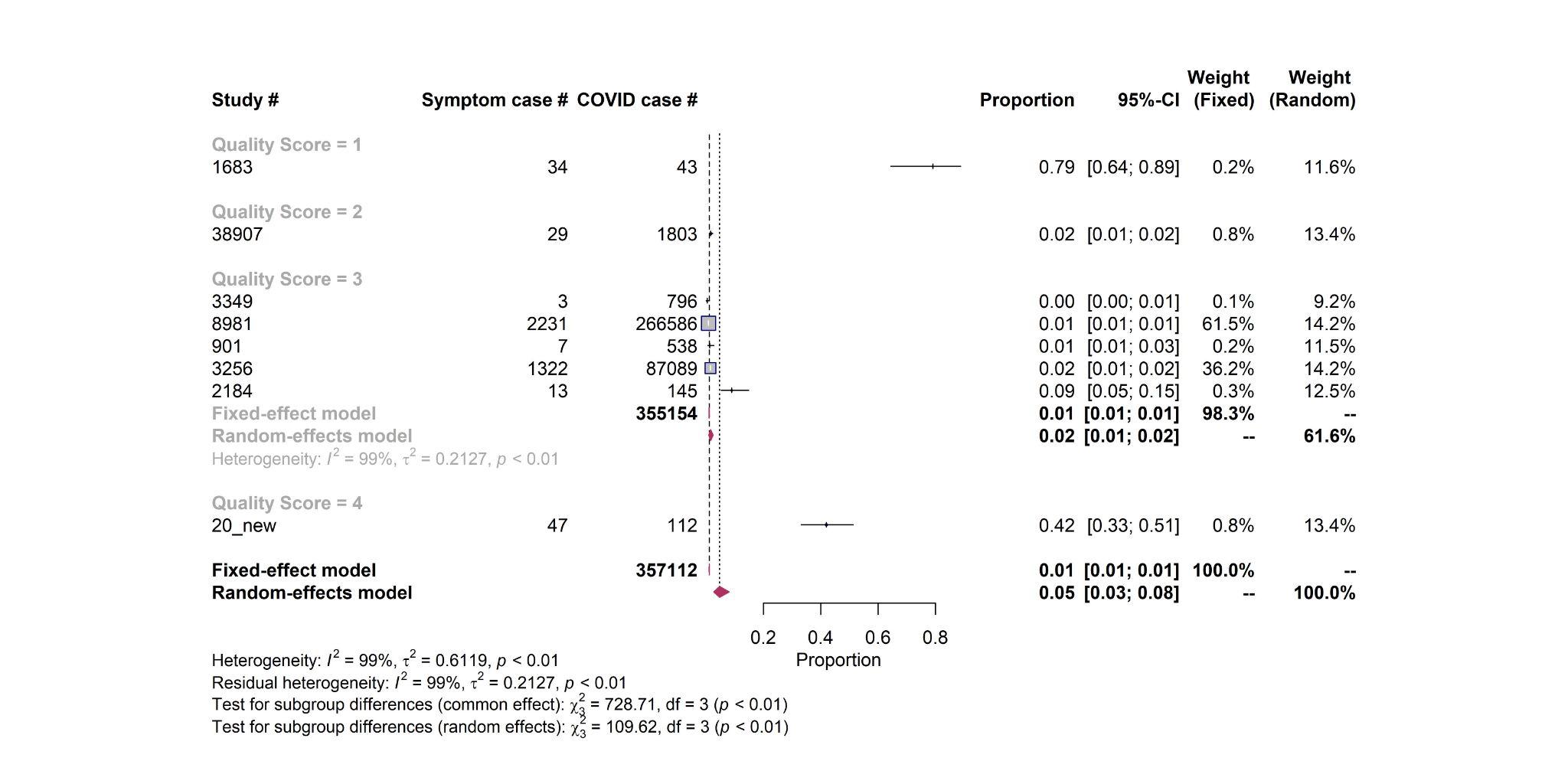

**Cardiac abnormalities – Sample size**

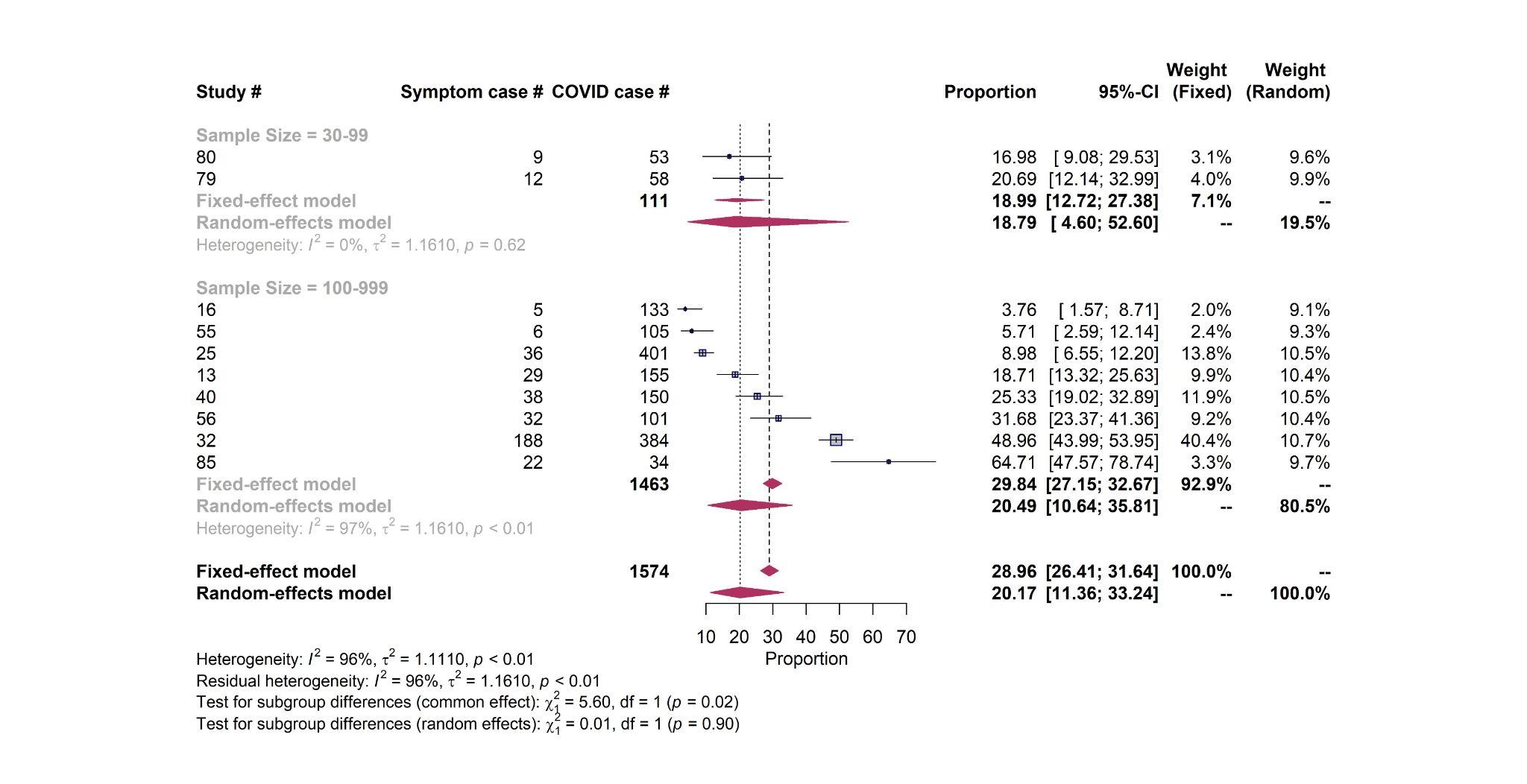

**Cardiac abnormalities – Sampling representativeness**

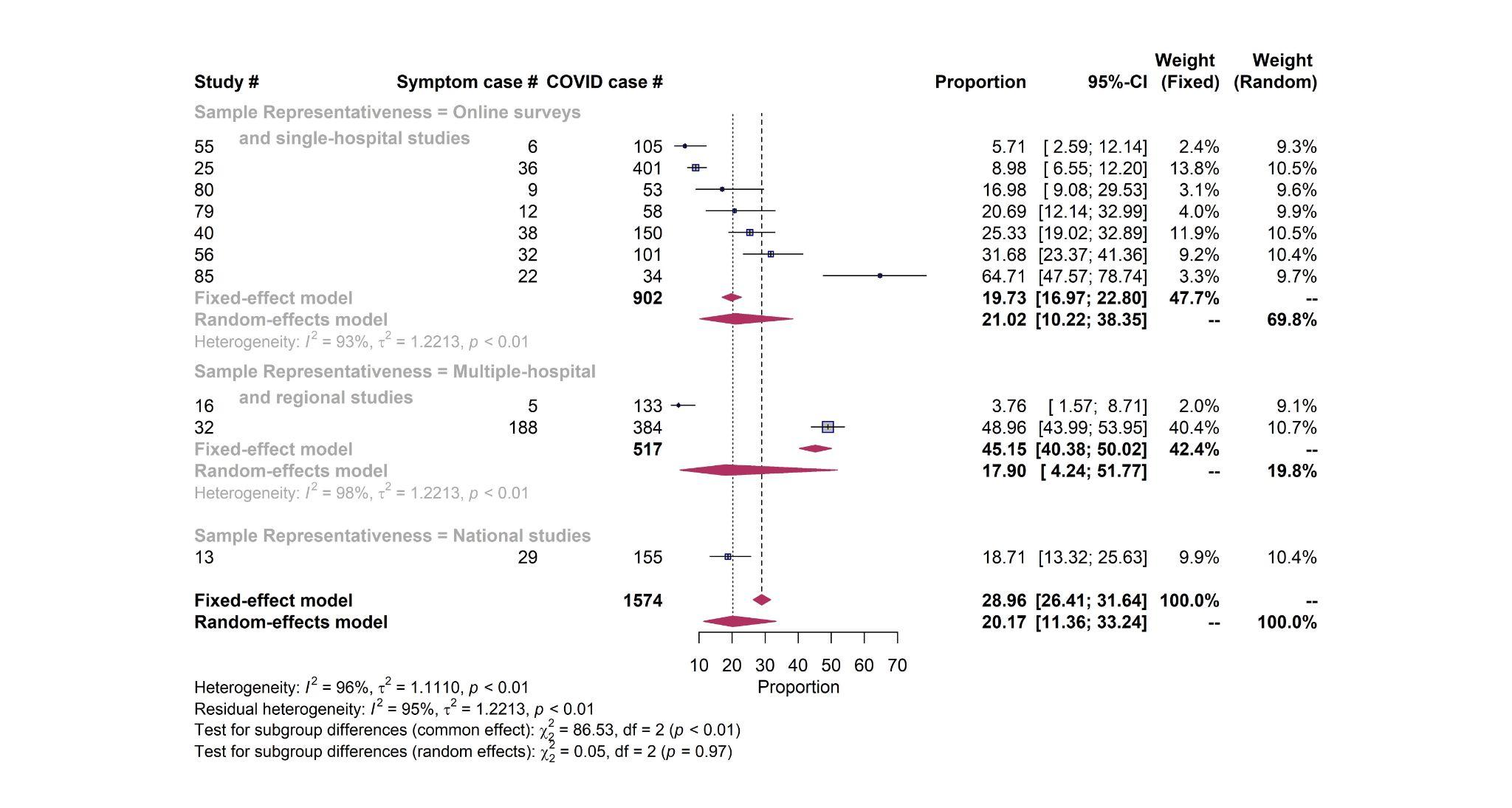

**Cardiac abnormalities – Study design**

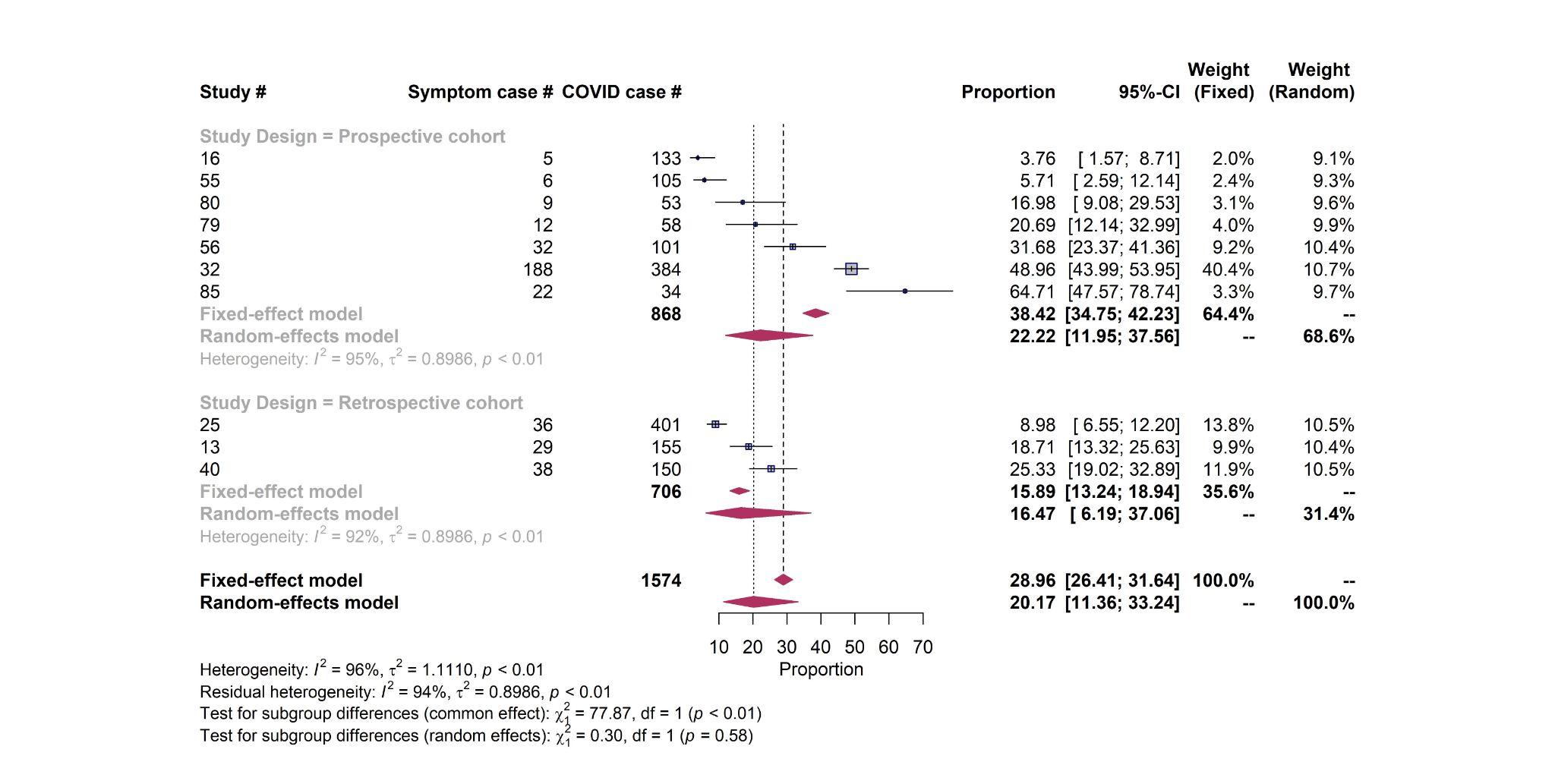

**Myocardial injury – Quality score**

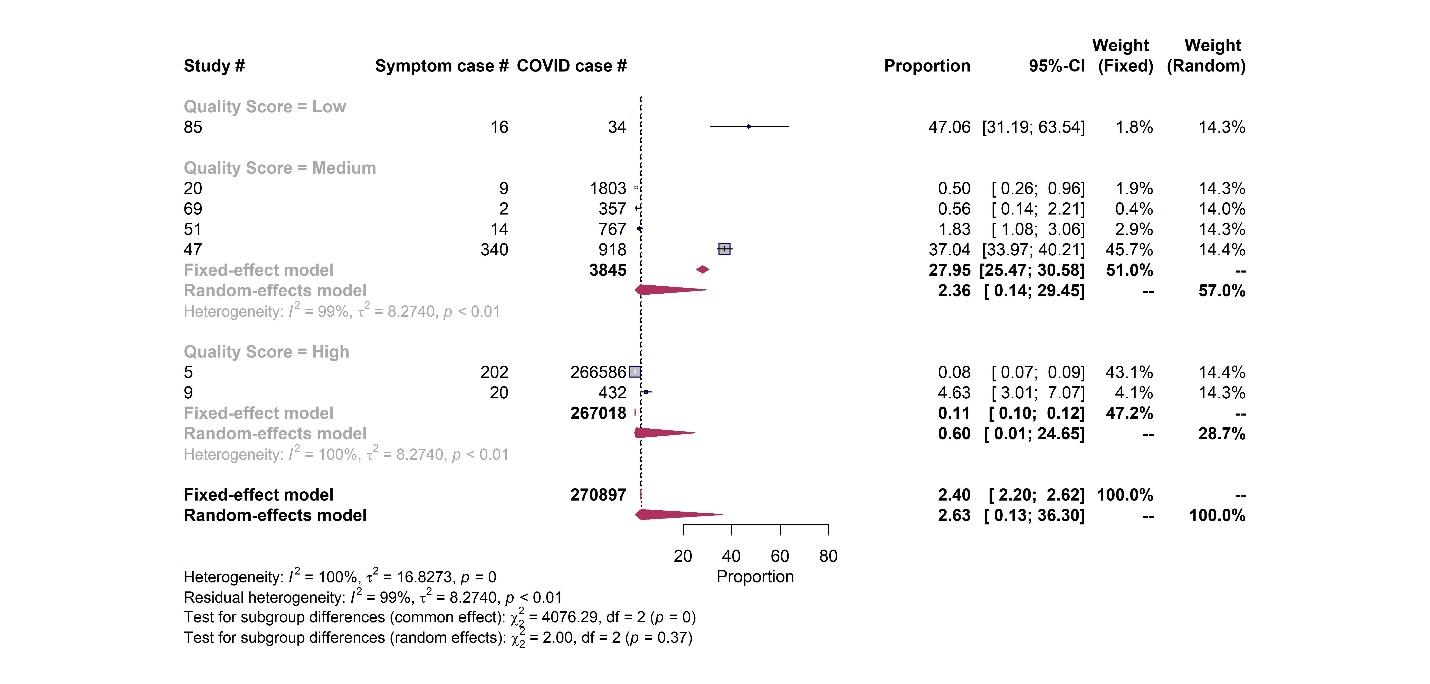

**Myocardial injury – Sample size**

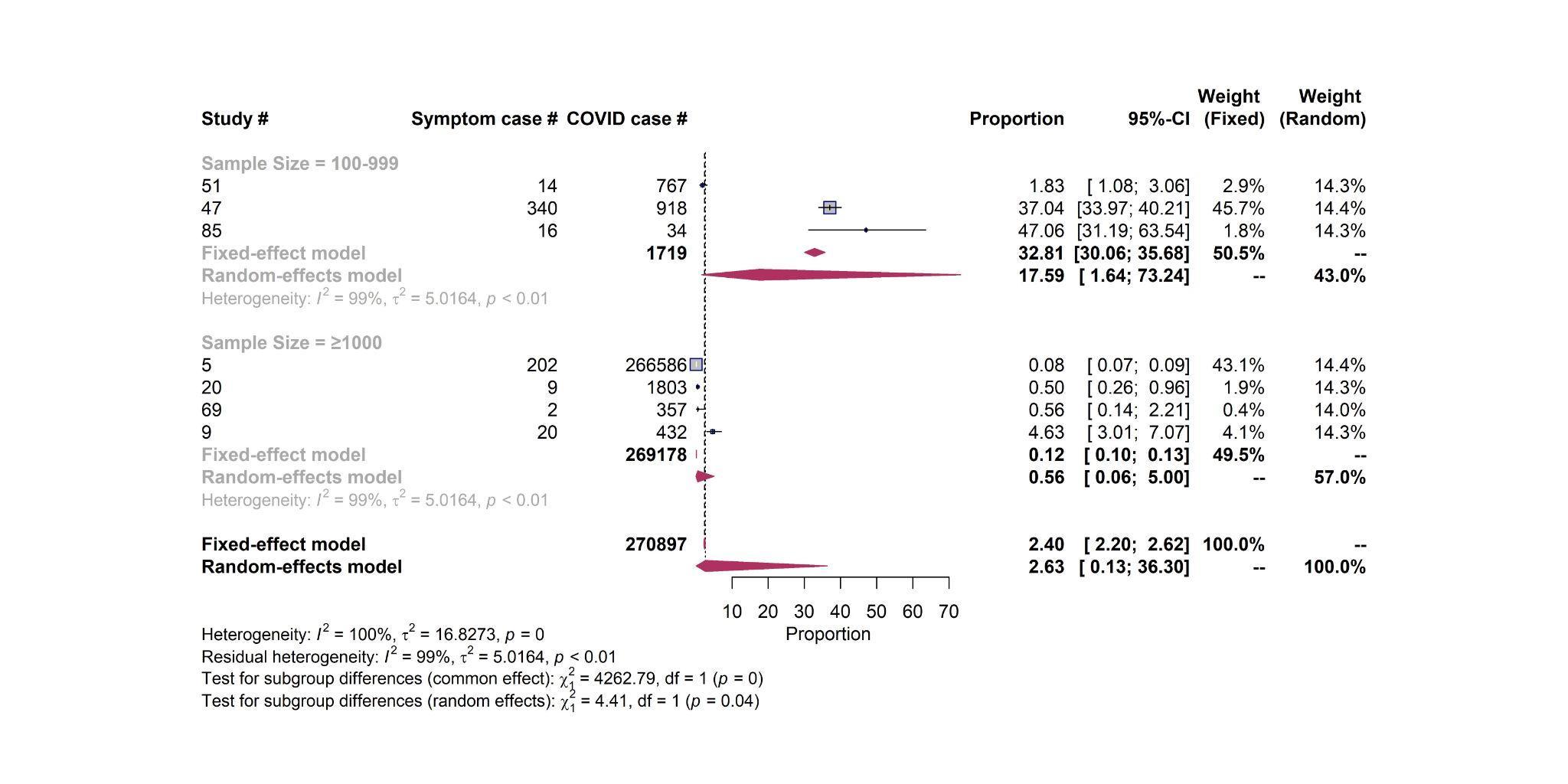

**Myocardial injury – Sampling representativeness**

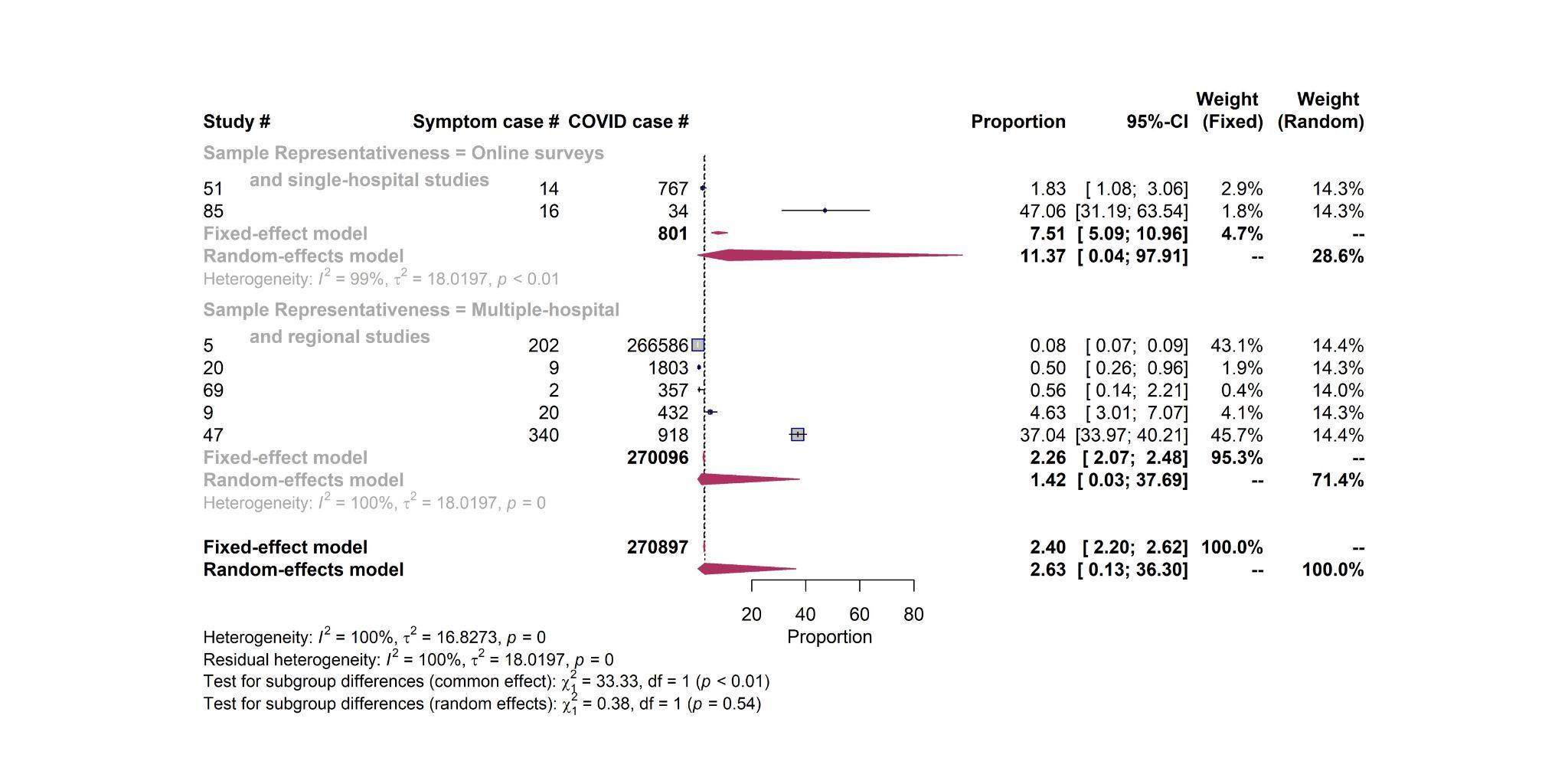

**Myocardial injury – Study design**

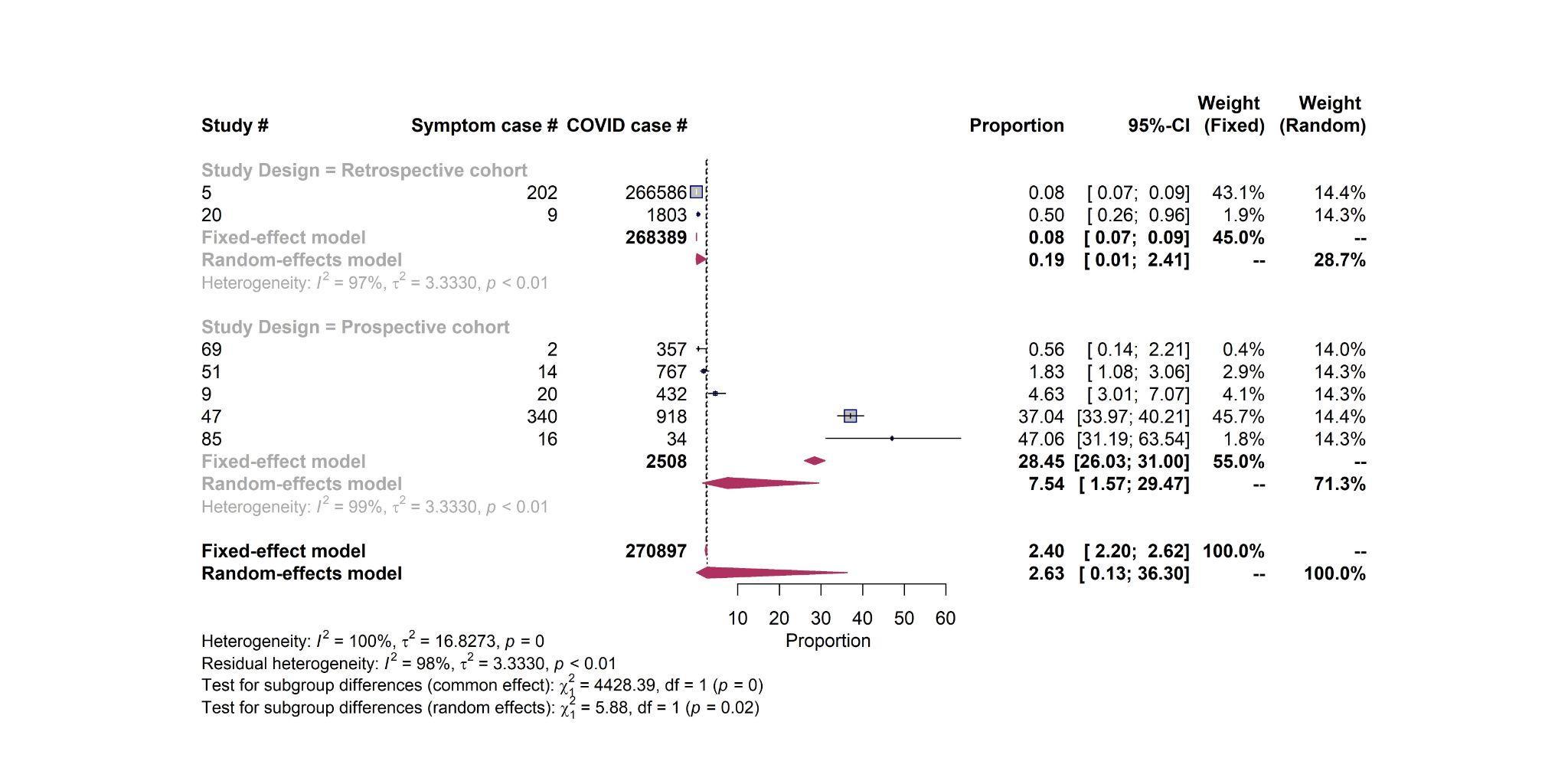

**Thromboembolism – Quality score**

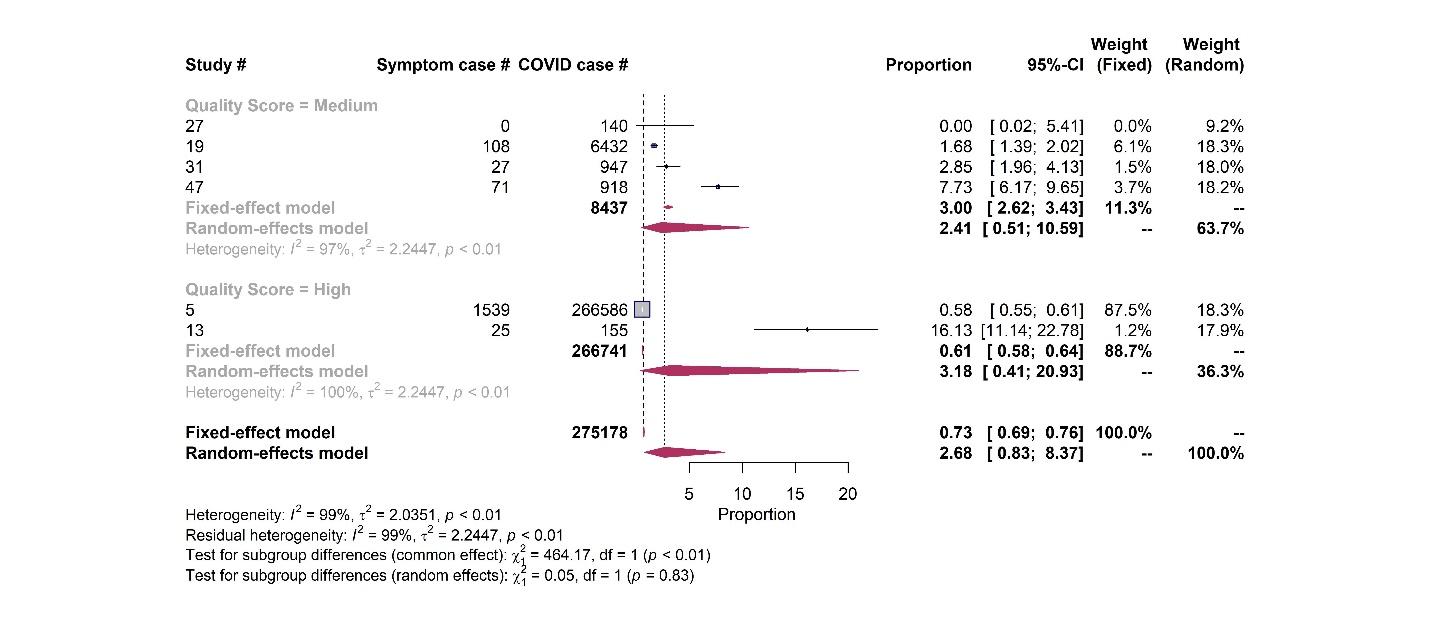

**Thromboembolism – Sample size**

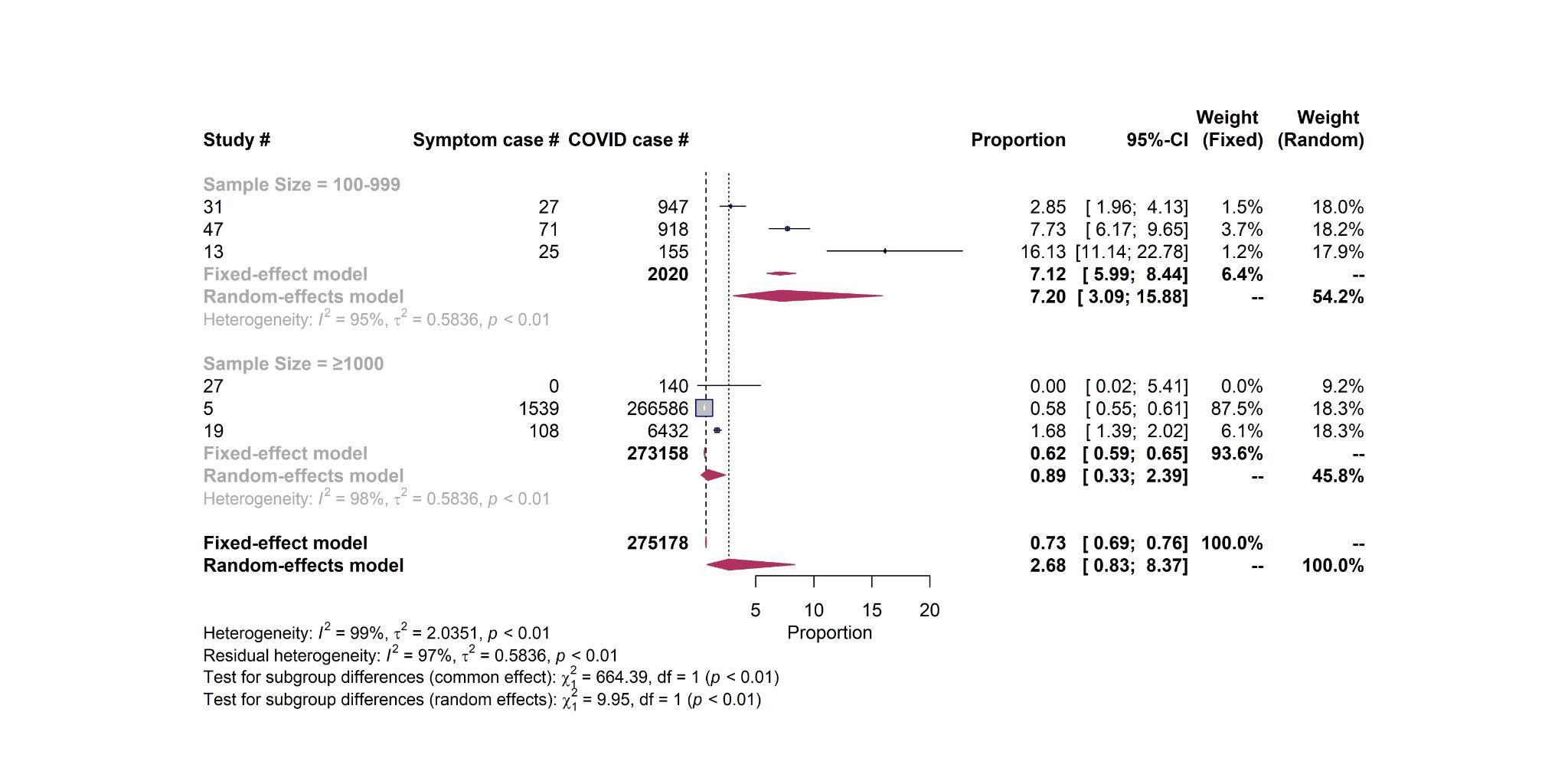

**Thromboembolism – Sampling representativeness**

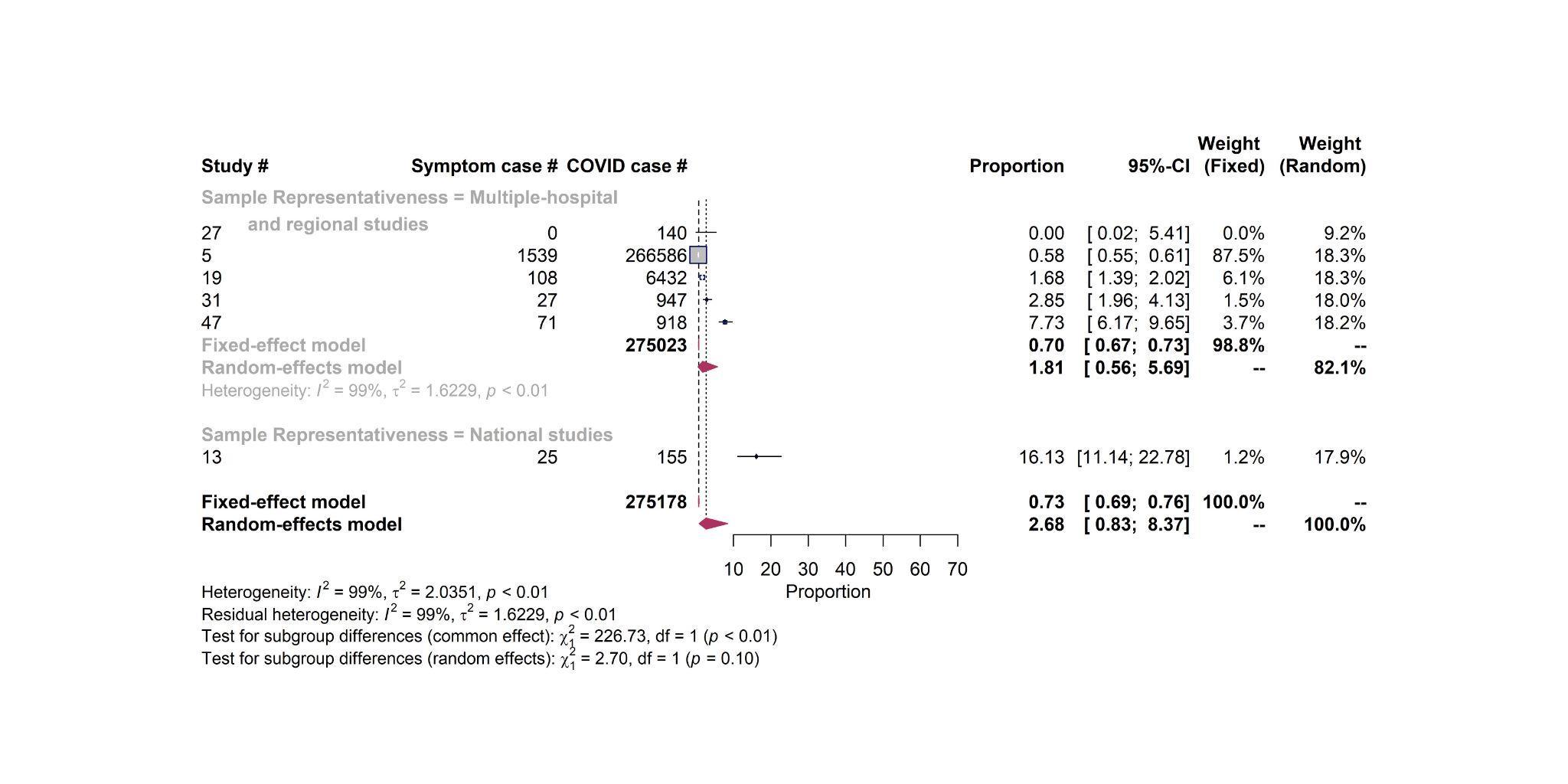

**Thromboembolism – Study design**

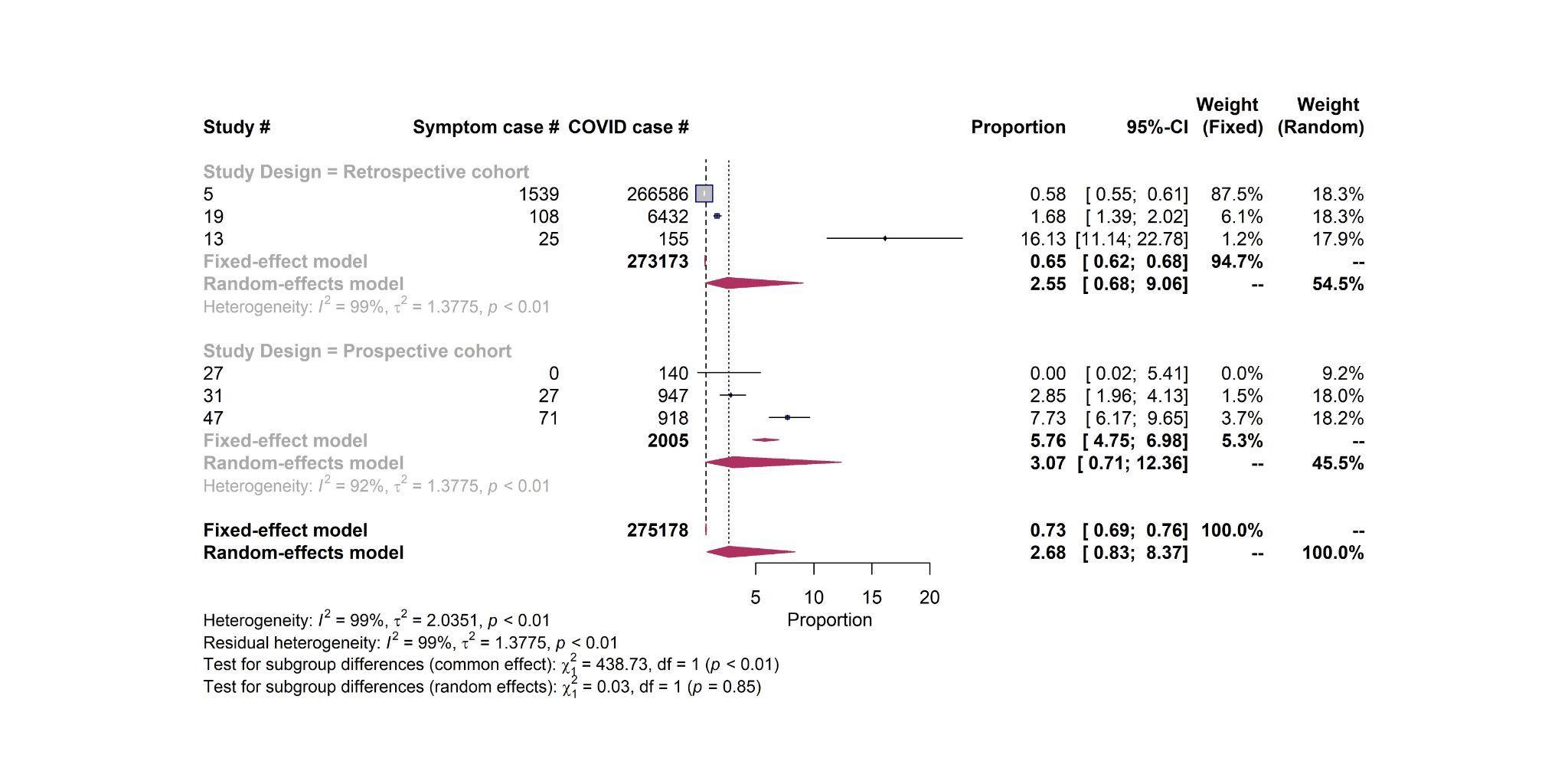

**Stroke – Quality score**

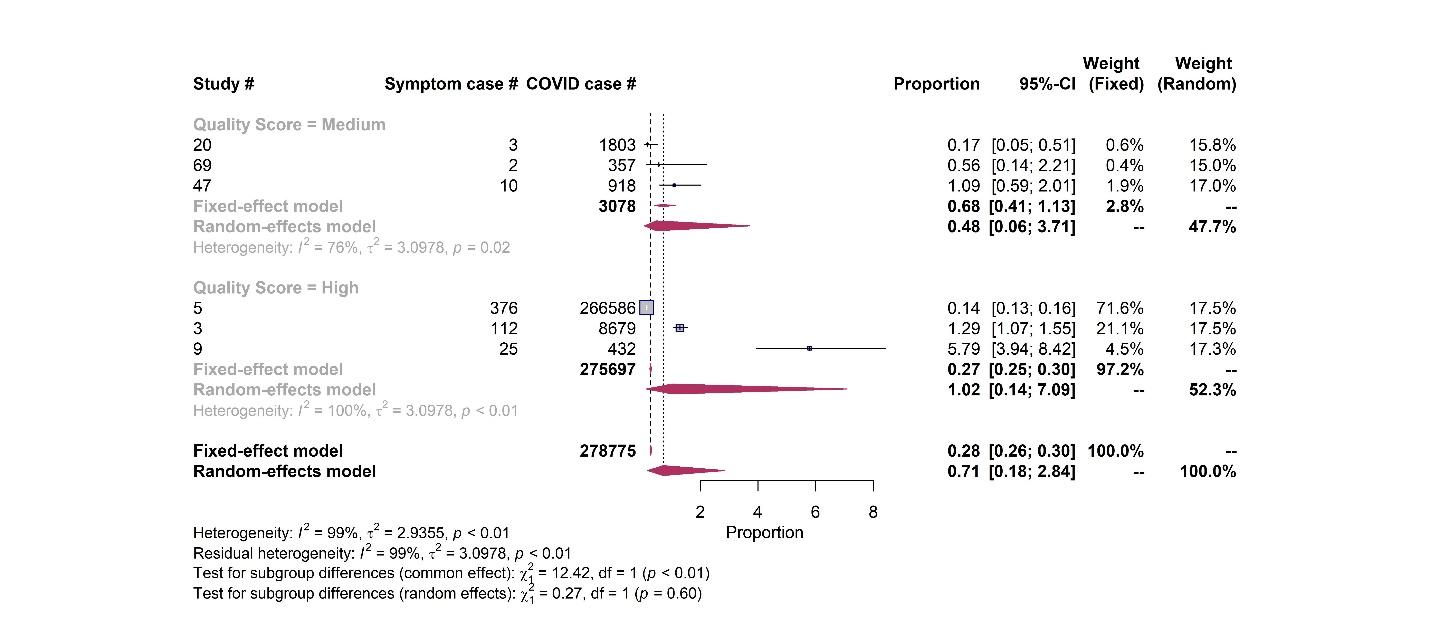

**Stroke – Sample size**

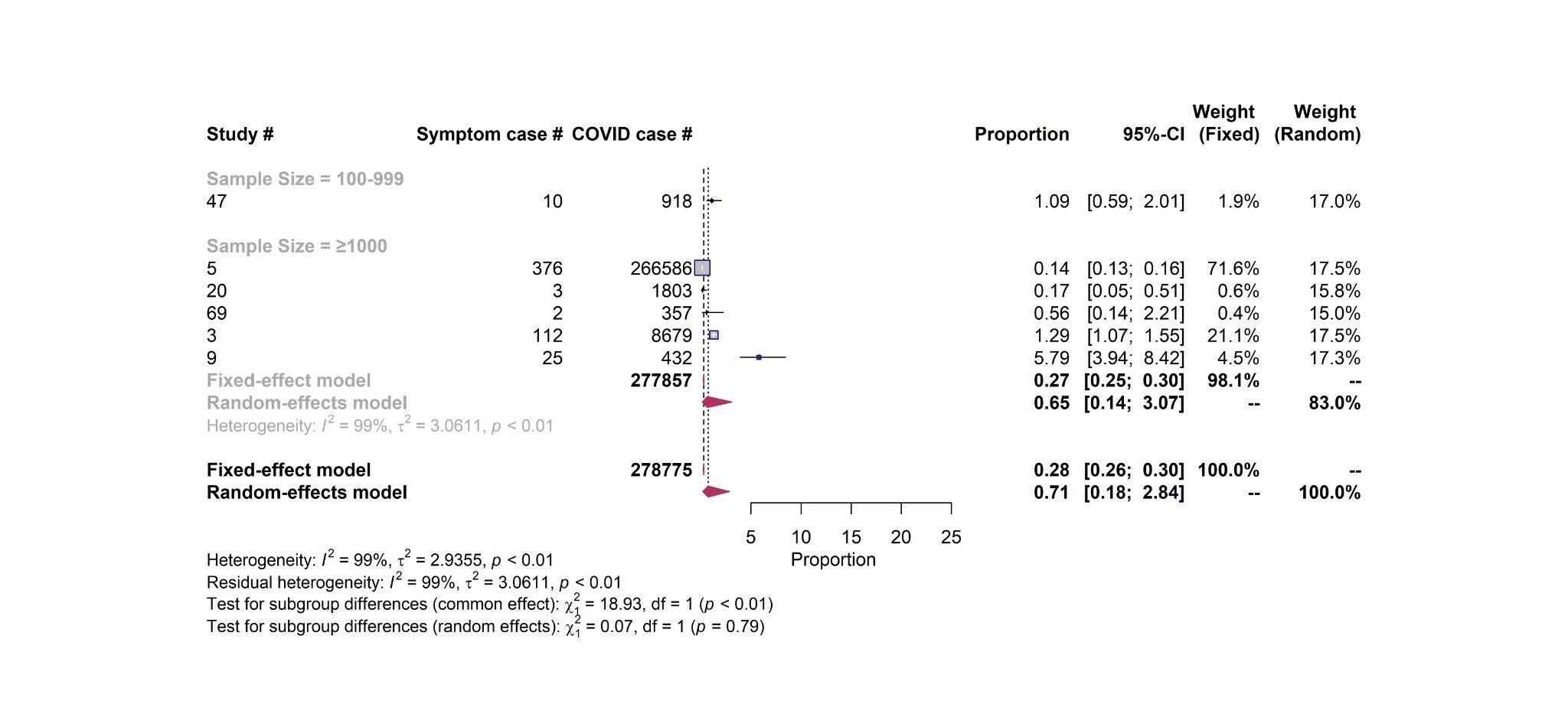

**Stroke – Sampling representativeness**

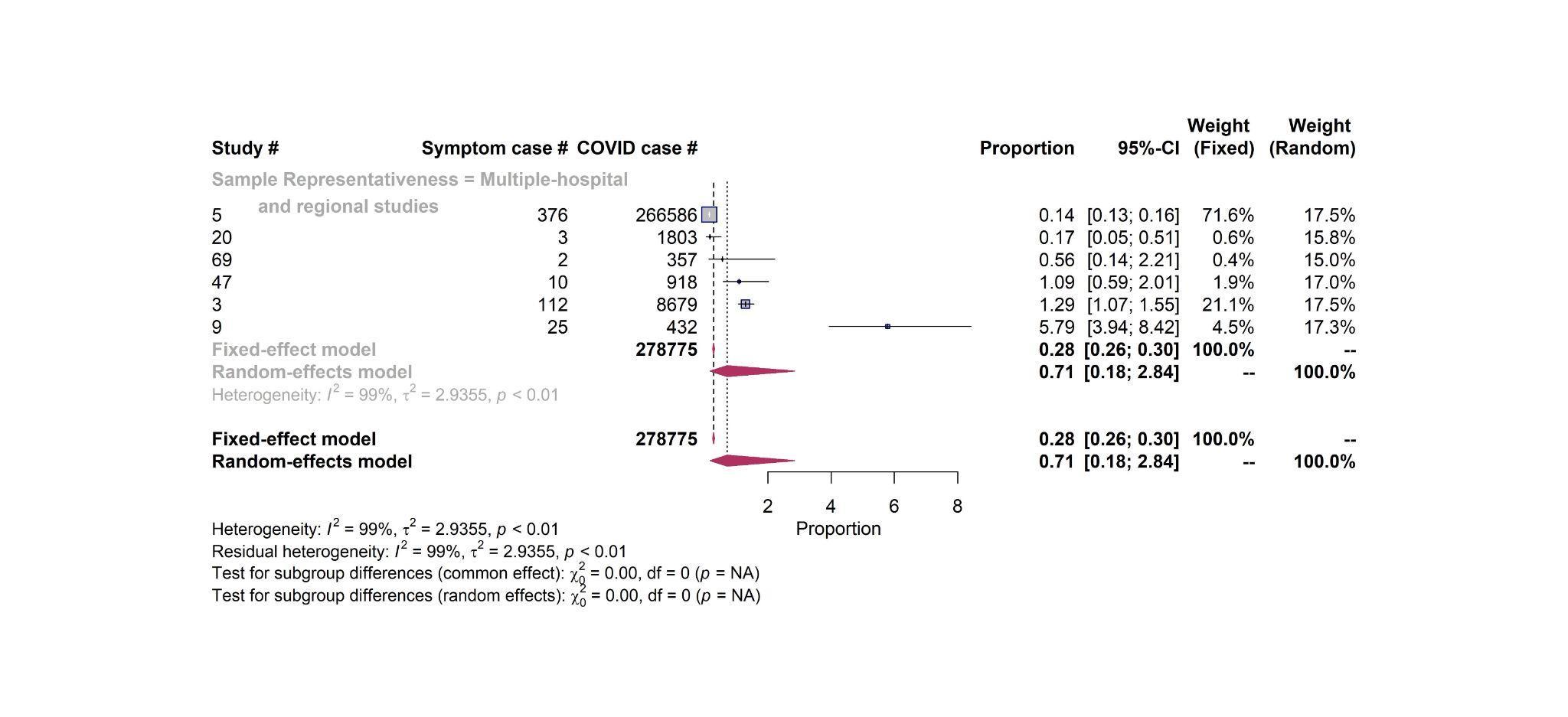

**Stroke – Study design**

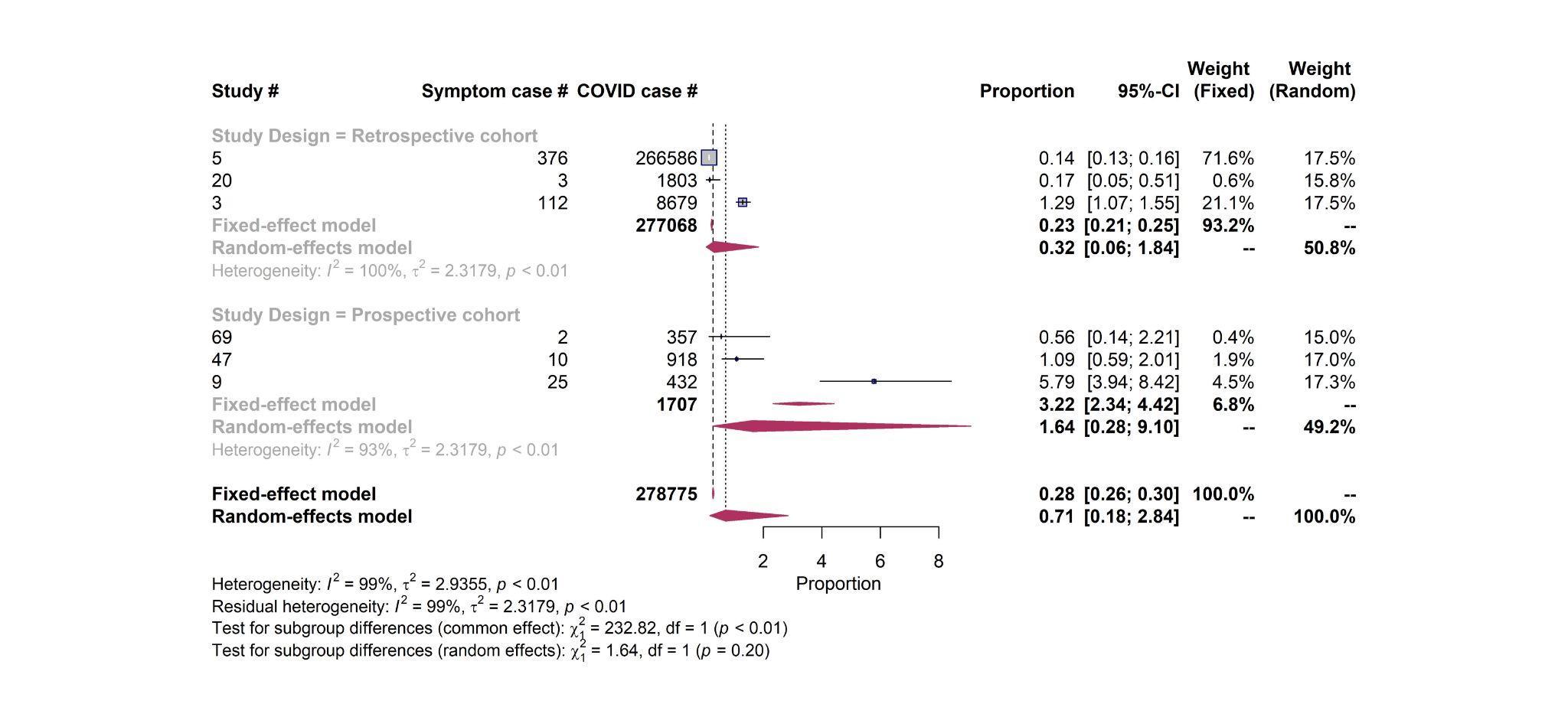

**Heart failure – Quality score**

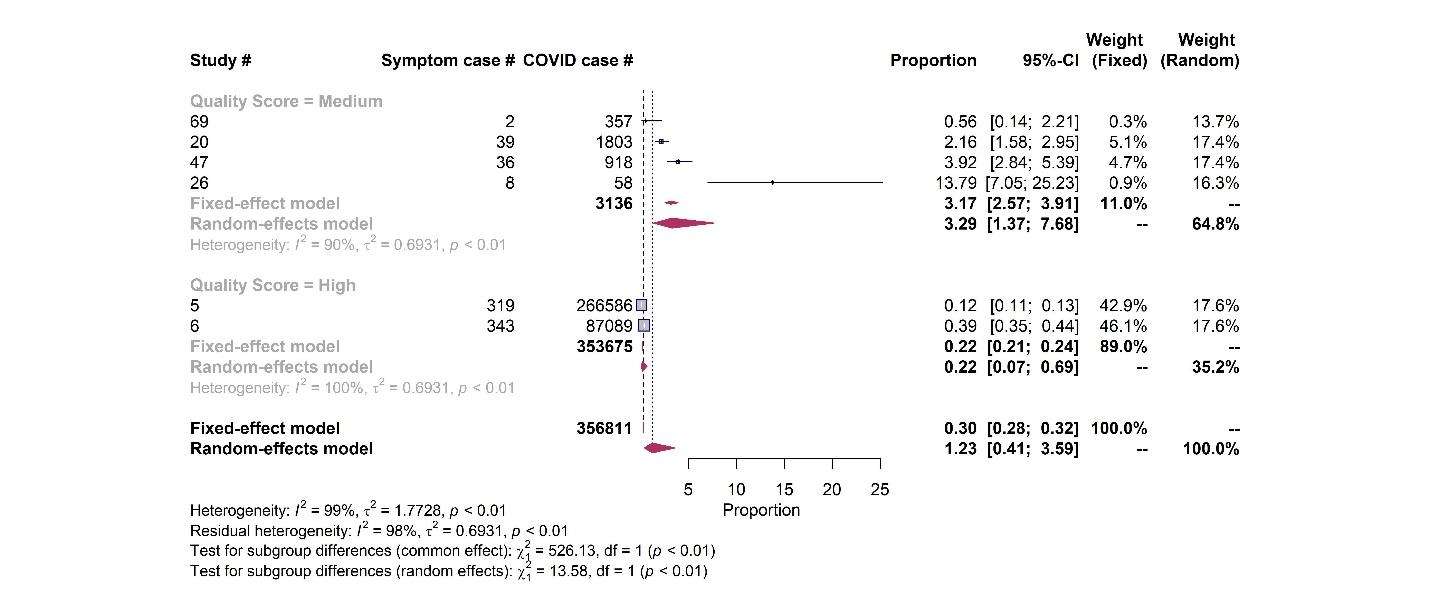

**Heart failure – Sample size**

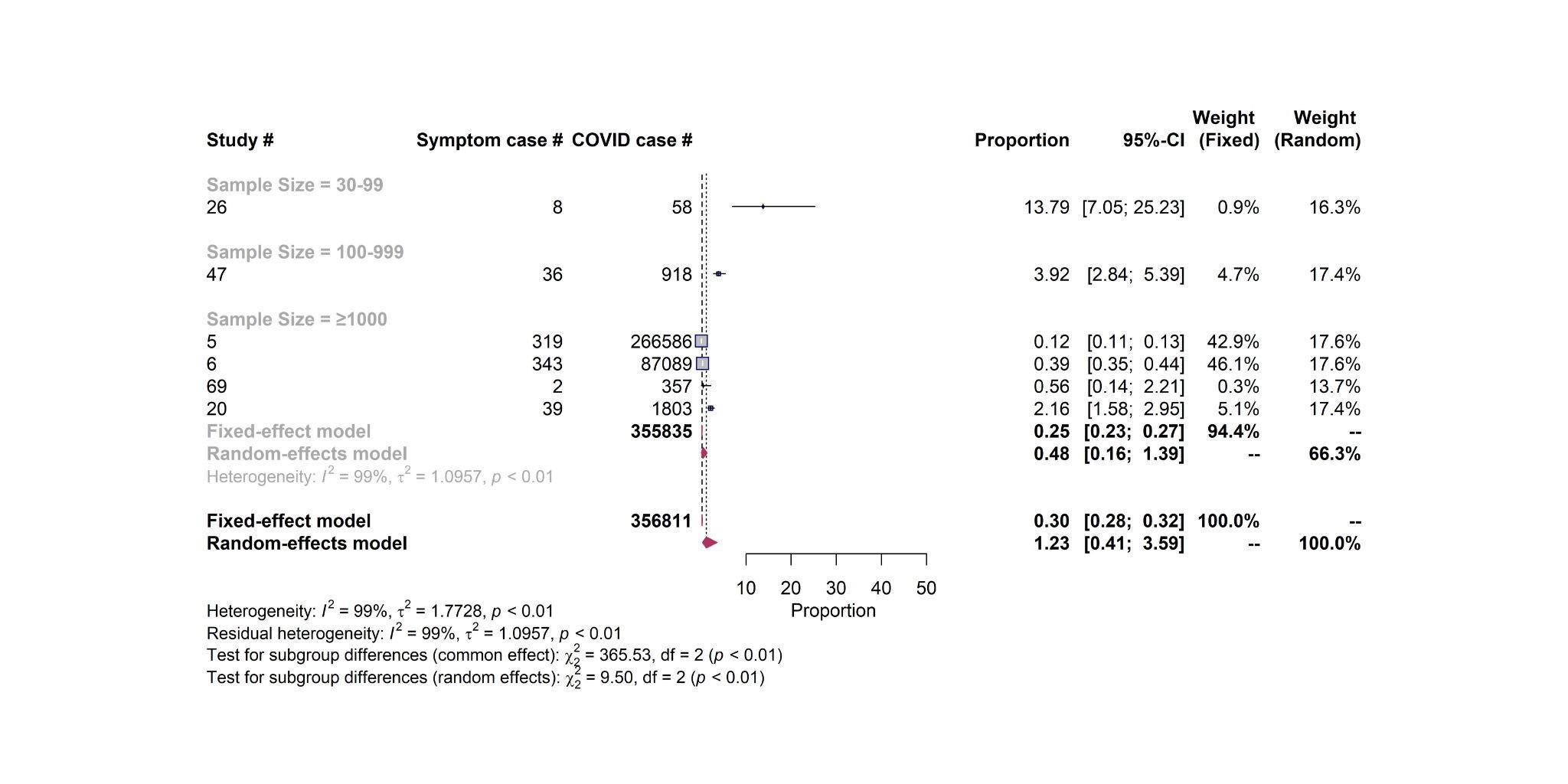

**Heart failure – Sampling representativeness**

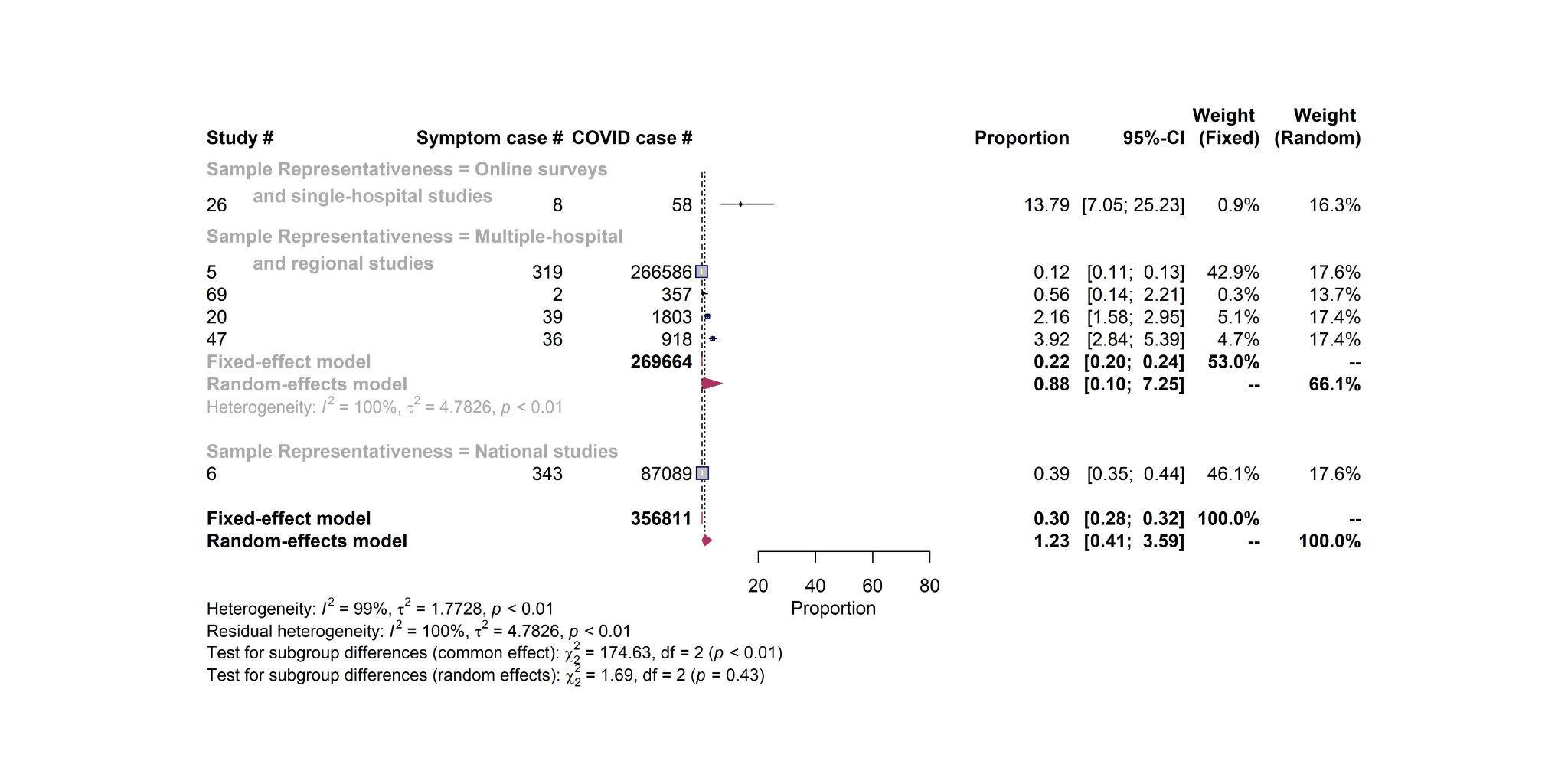

**Heart failure – Study design**

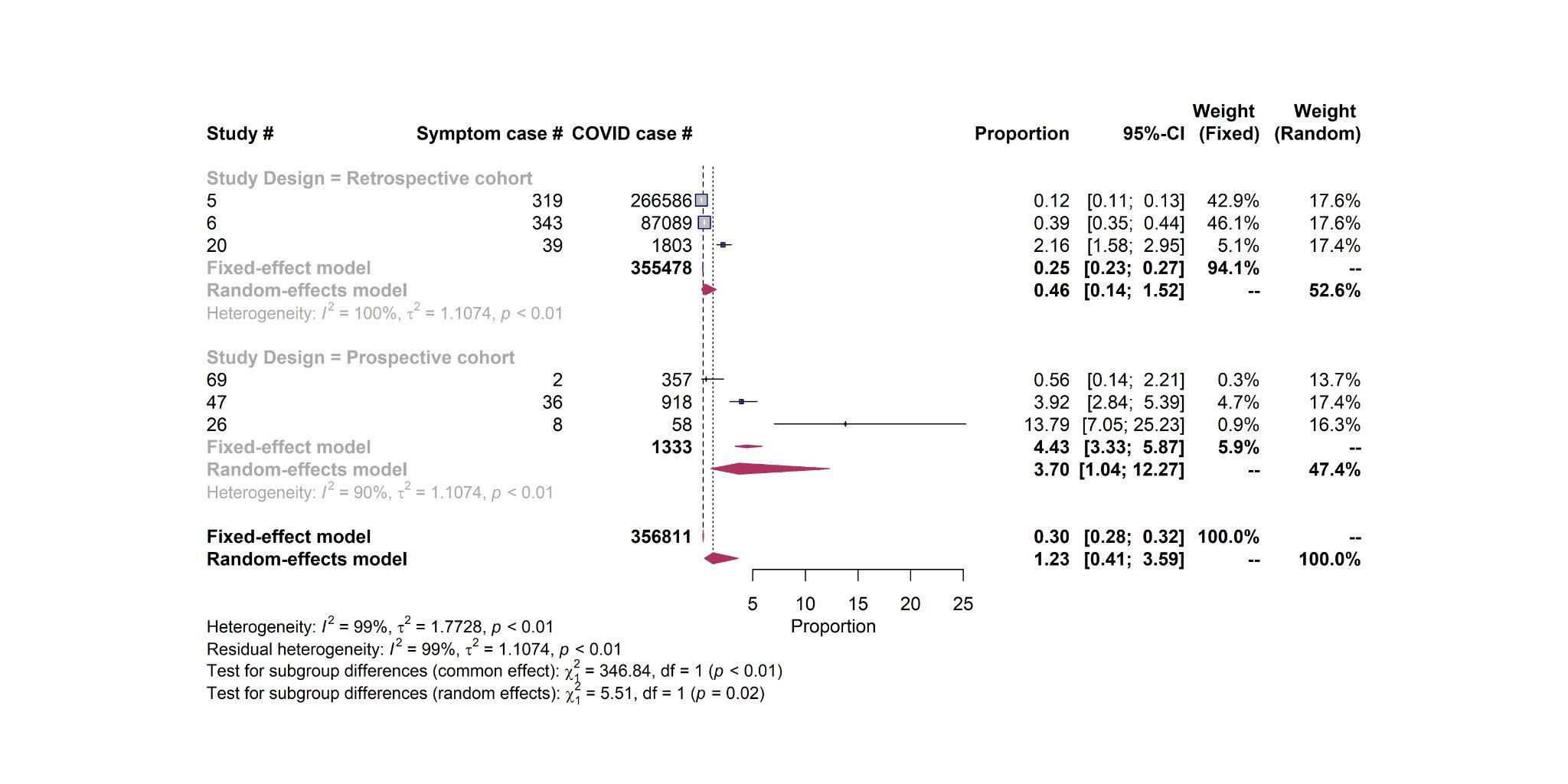

**Coronary disease – Quality score**

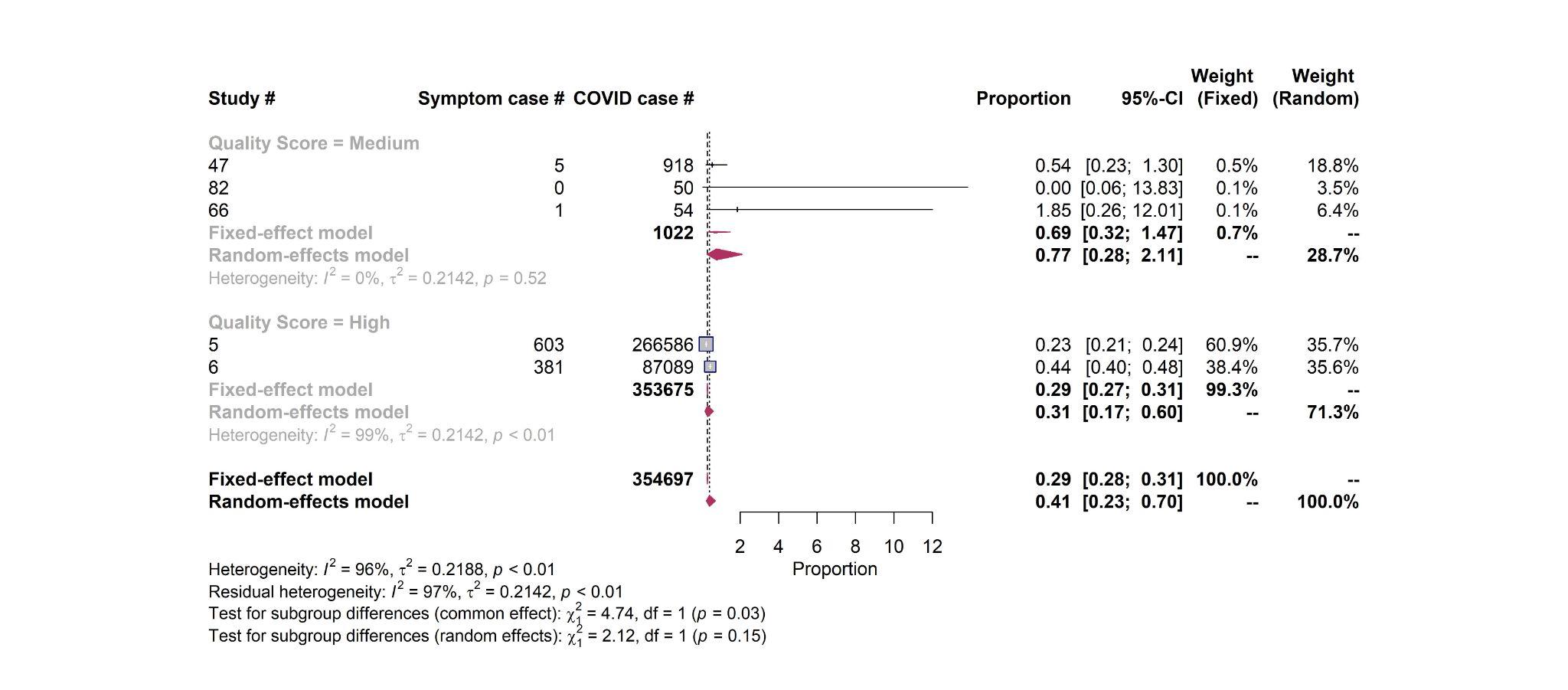

**Coronary disease – Sample size**

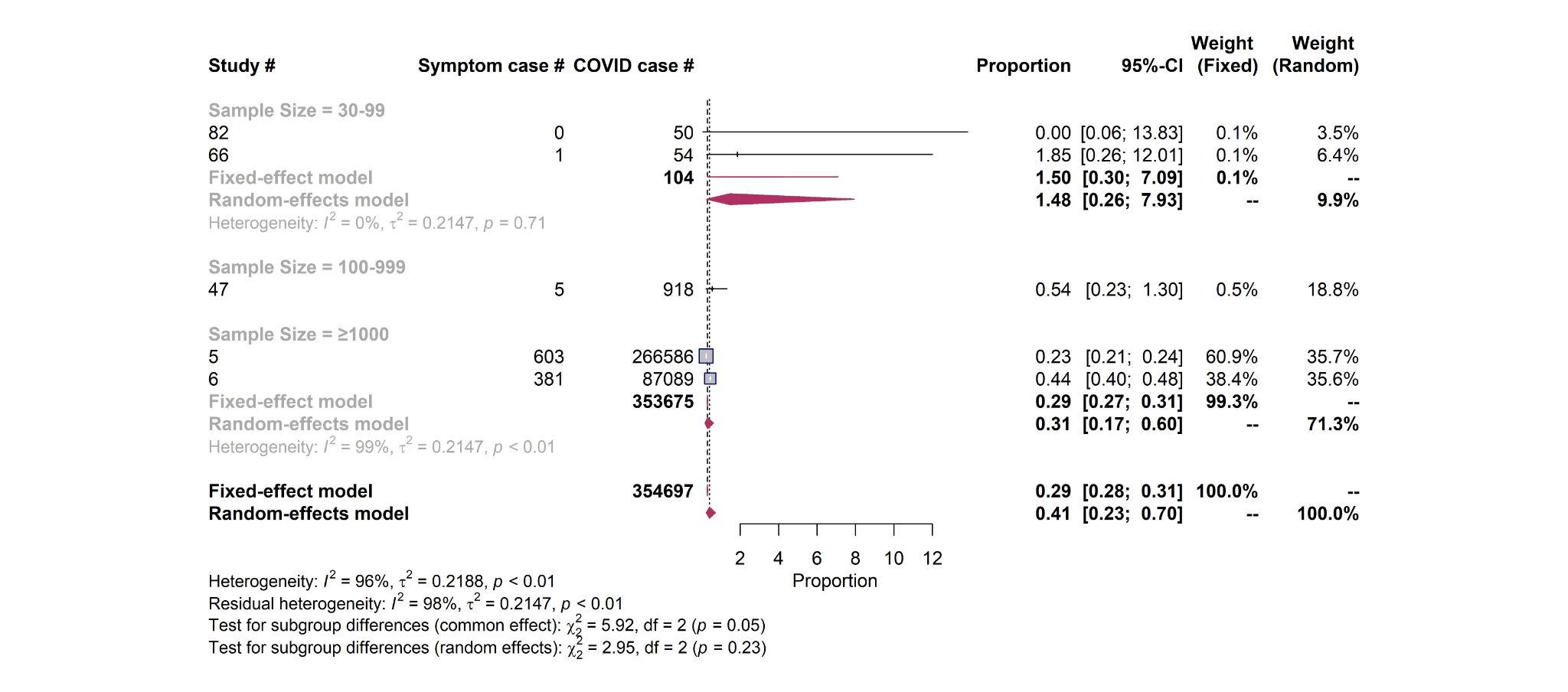

**Coronary disease – Sampling representativeness**

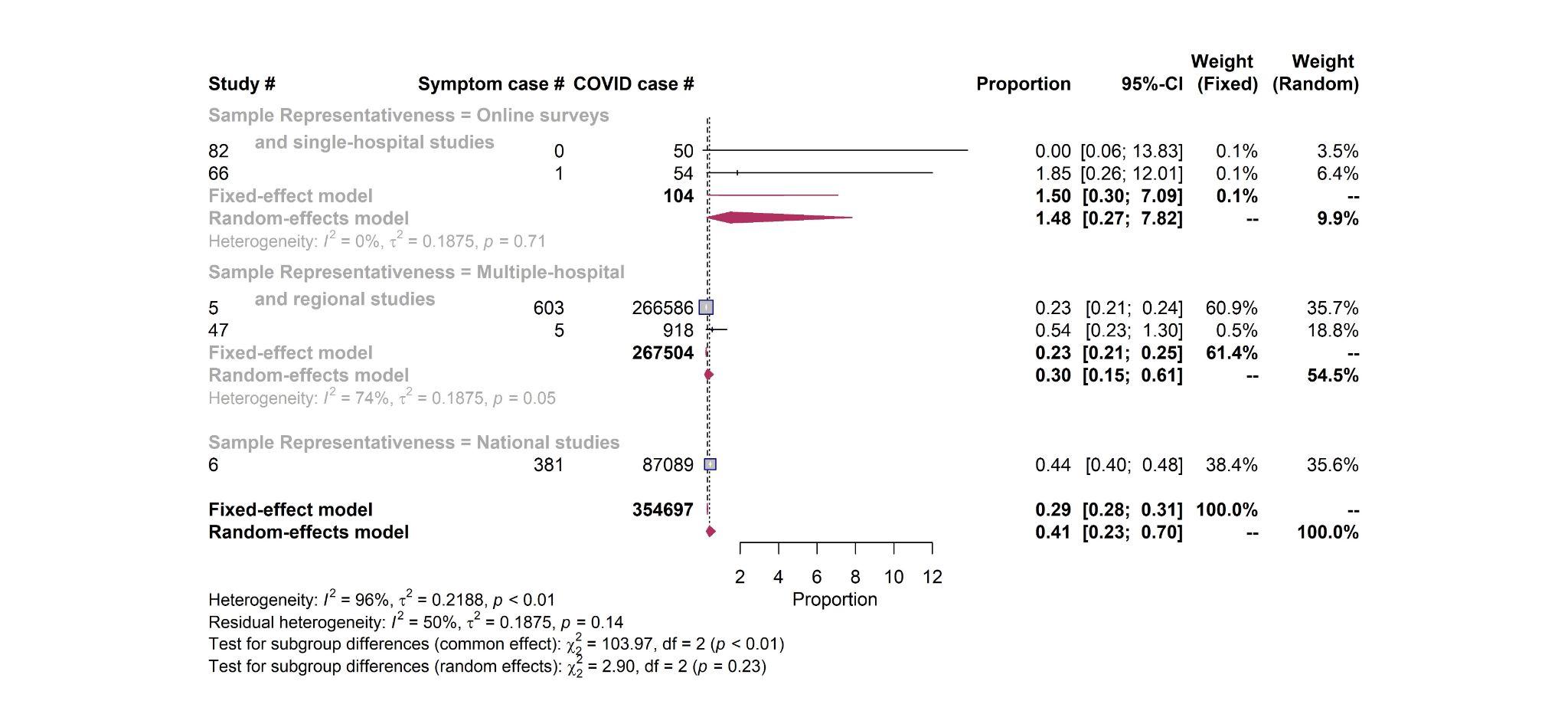

**Coronary disease – Study design**

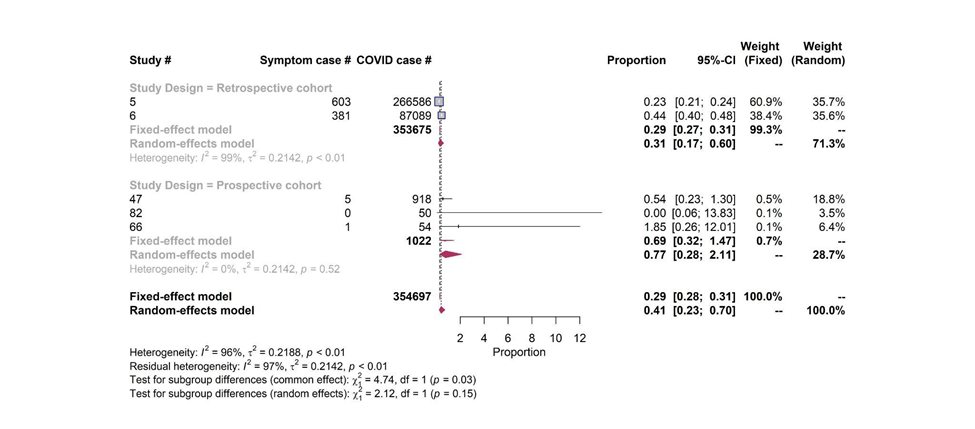

**Myocarditis – Quality score**

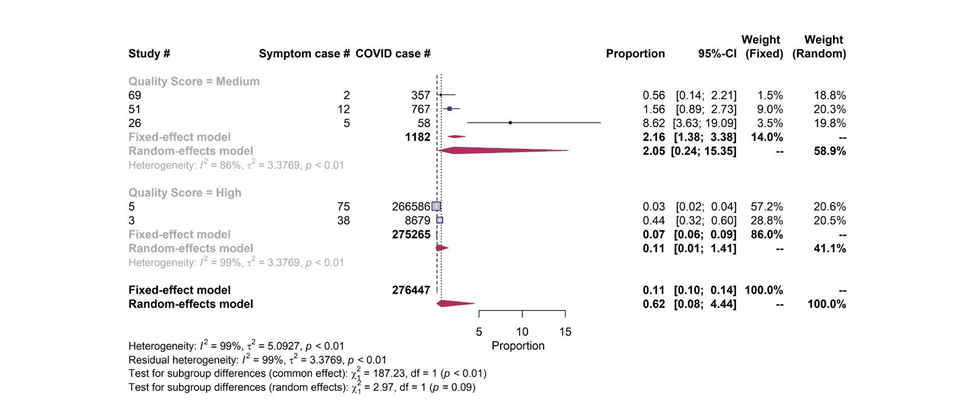

**Myocarditis – Sample size**

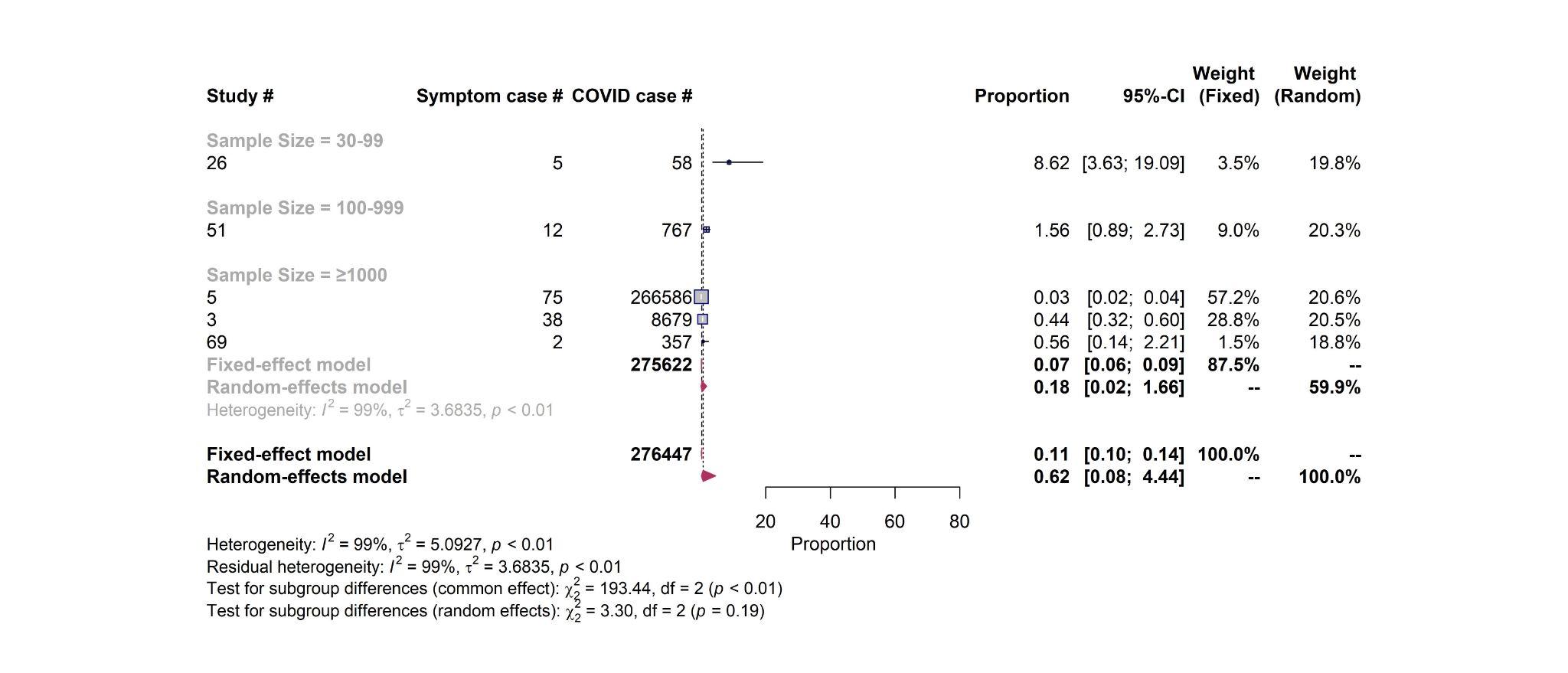

**Myocarditis – Sampling representativeness**

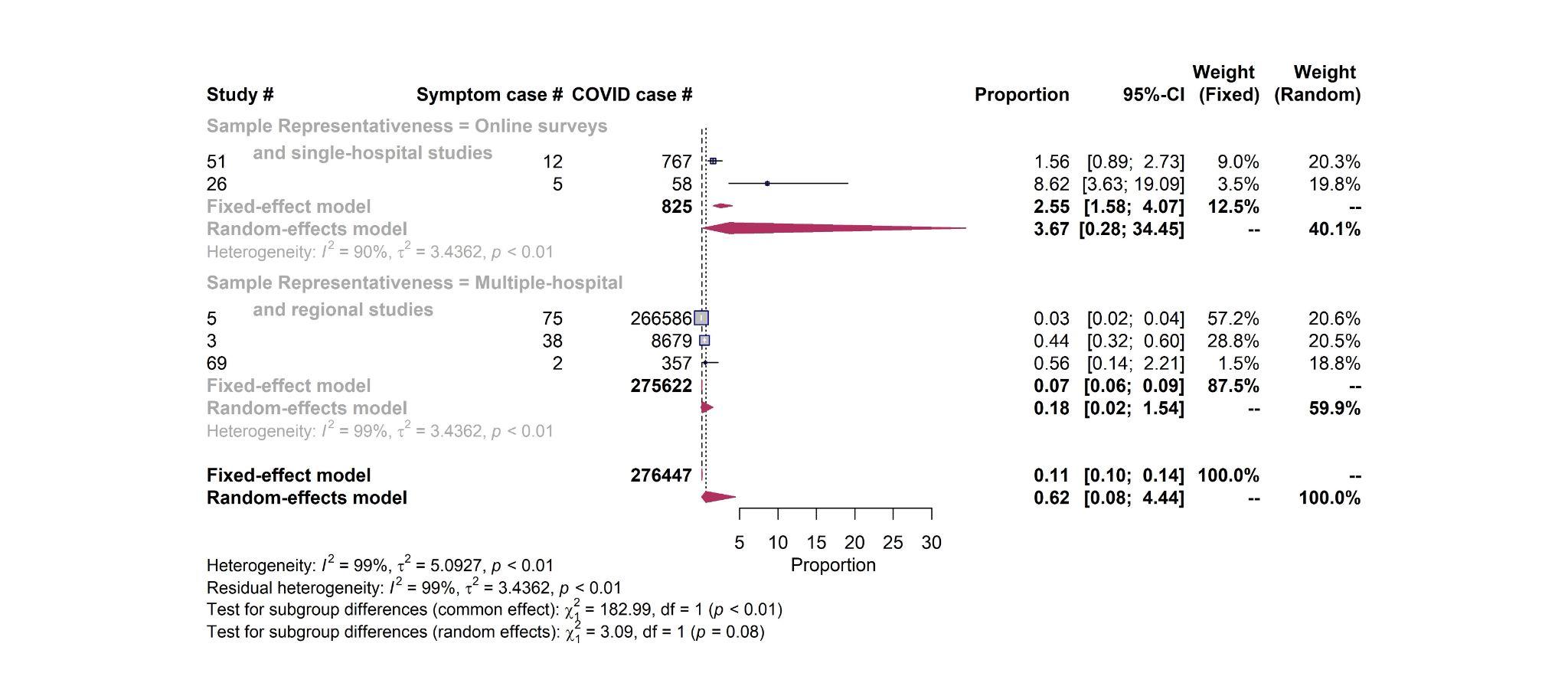

**Myocarditis – Study design**

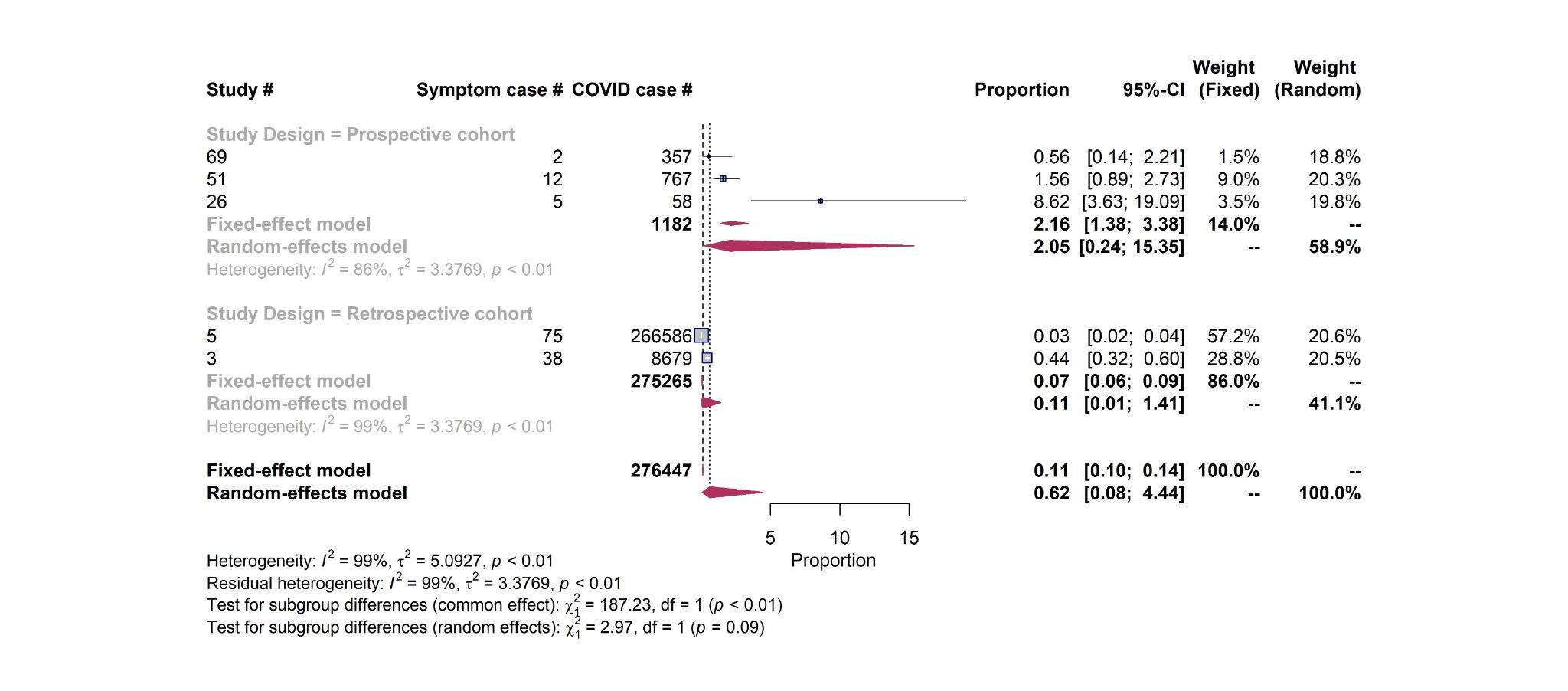

**Figure S4.** Funnel plots of top 10 most reported long-COVID cardiac symptoms.

**Chest pain**
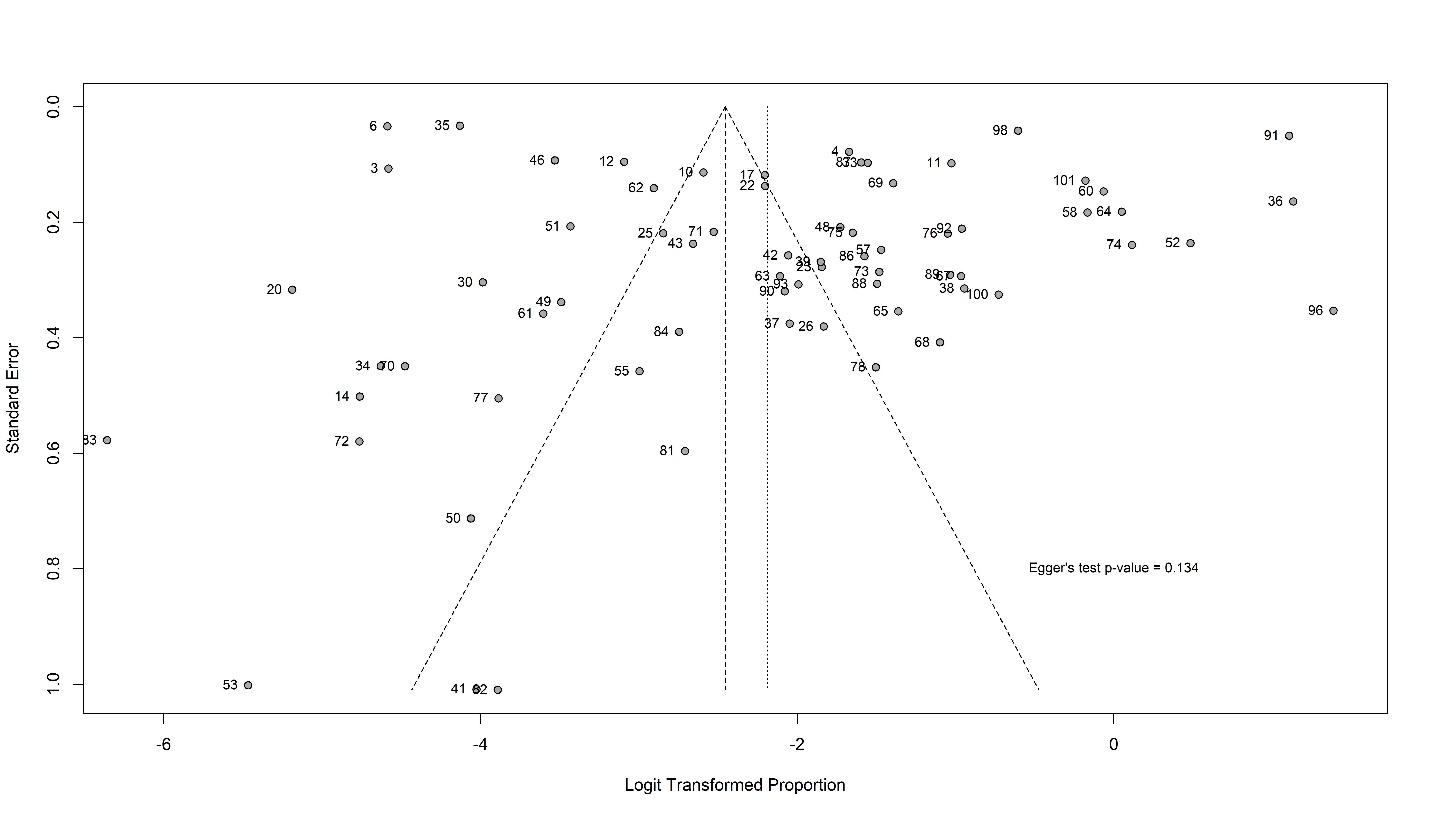

**Arrhythmia**

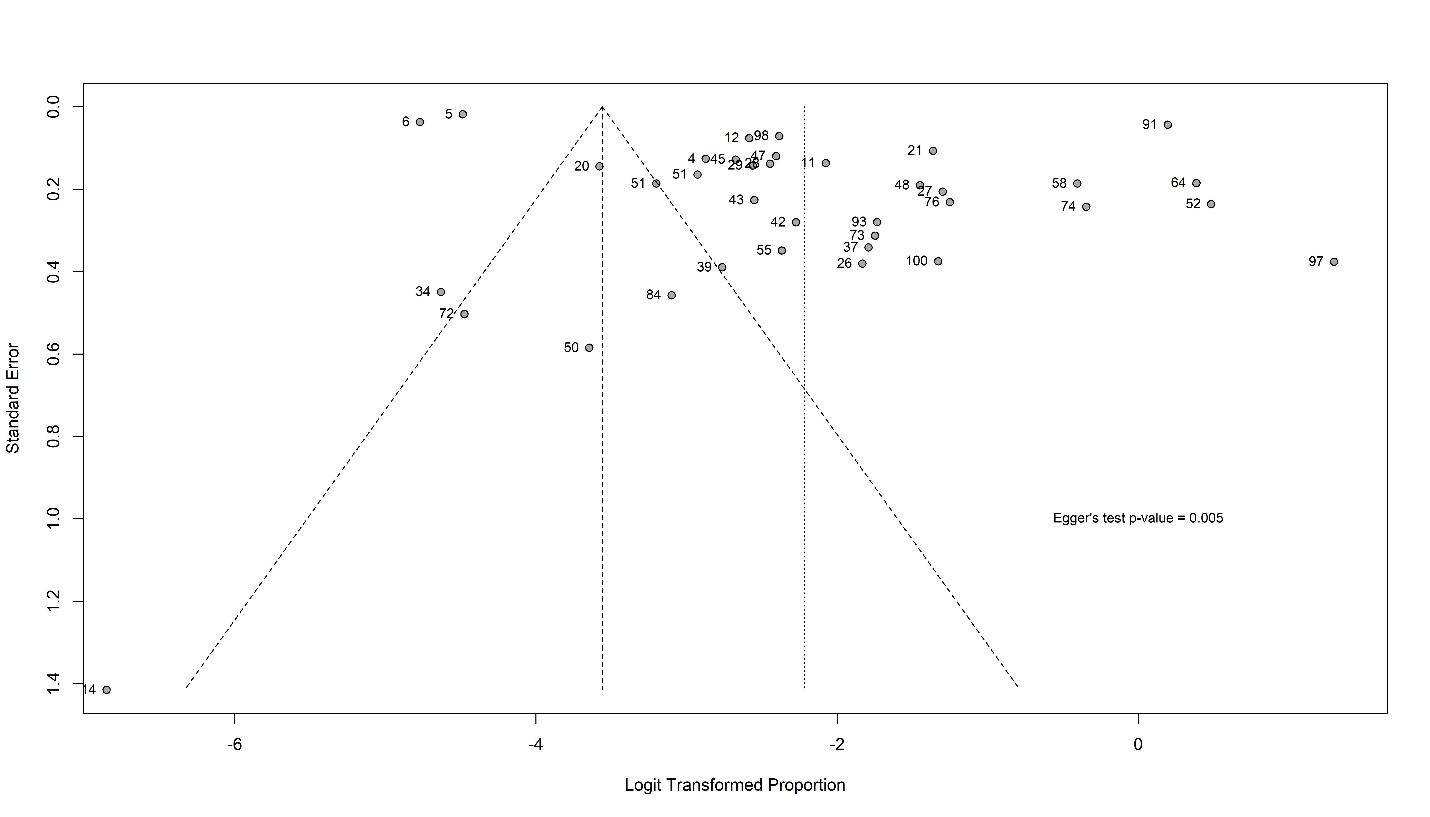

**Hypertension**

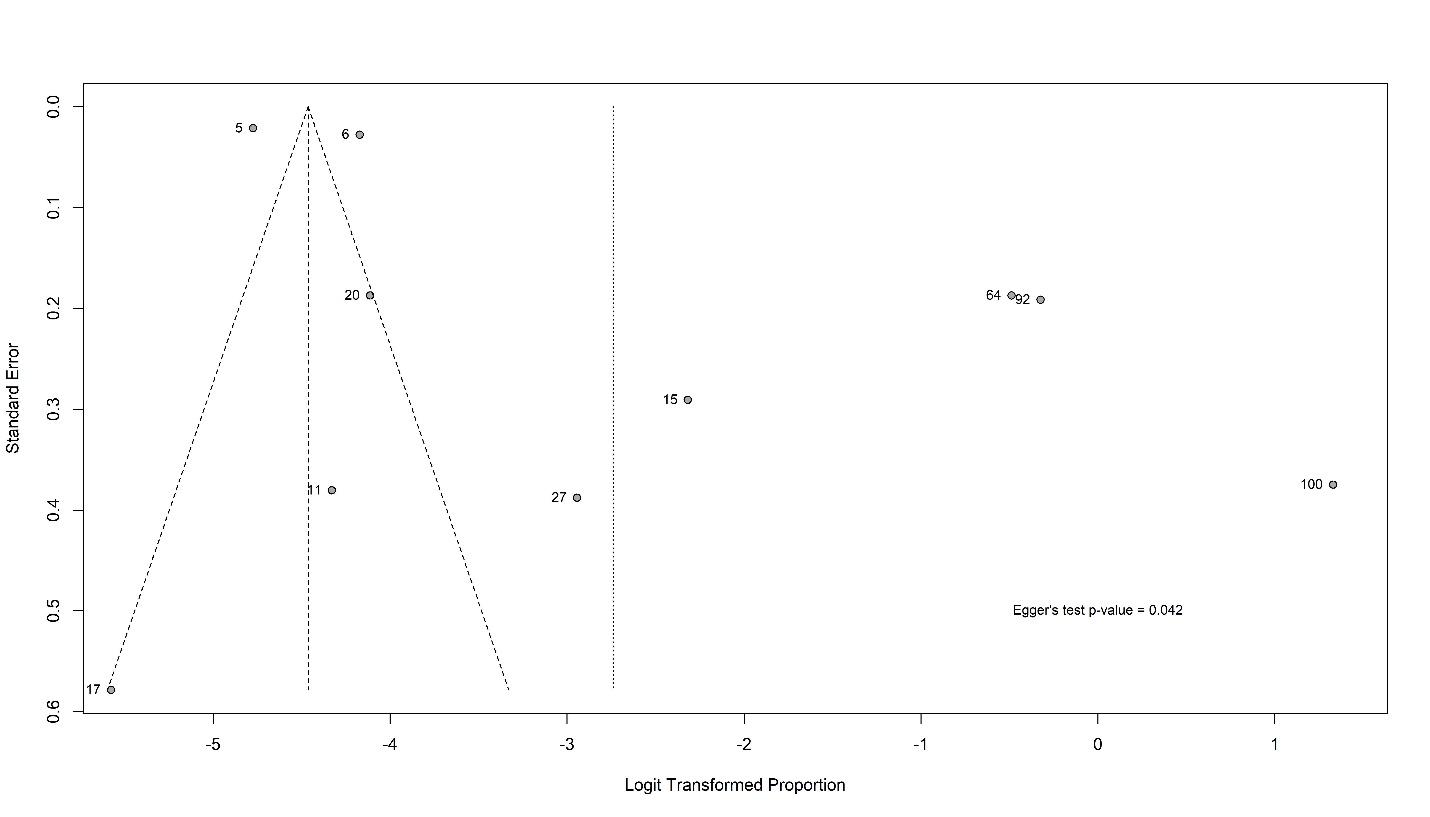

**Cardiac abnormalities**

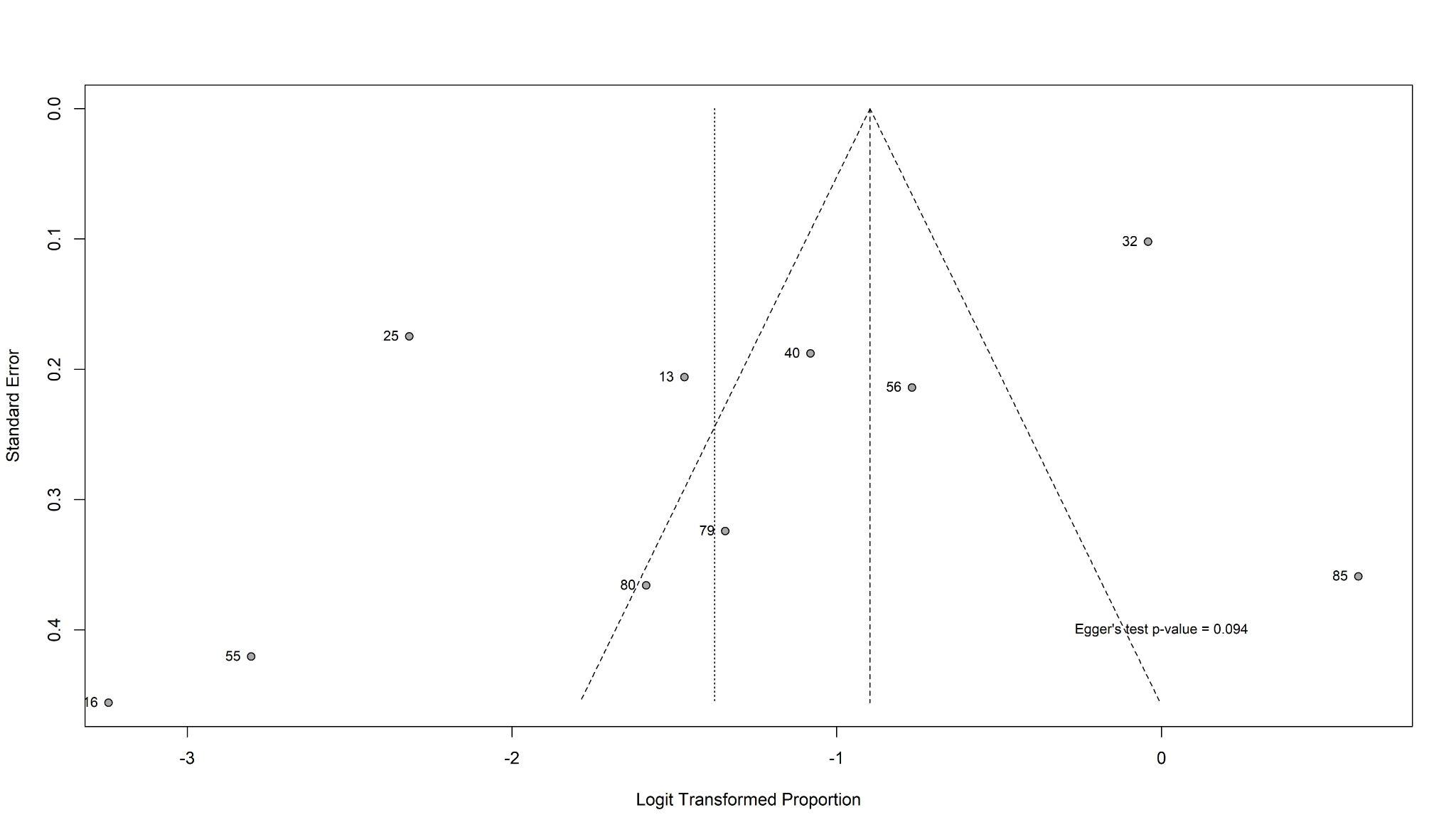

**Myocardial injury**

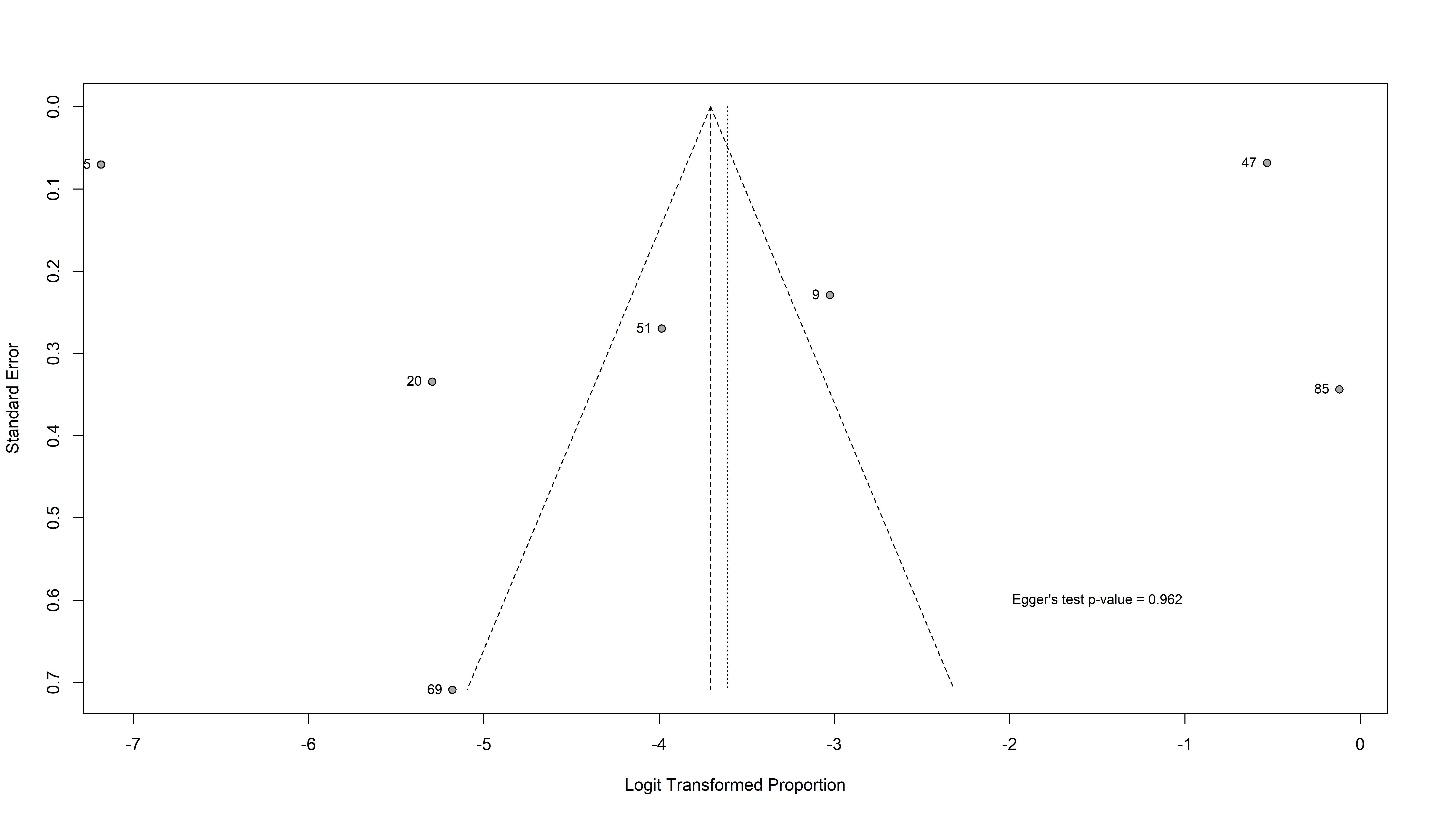

**Thromboembolism**

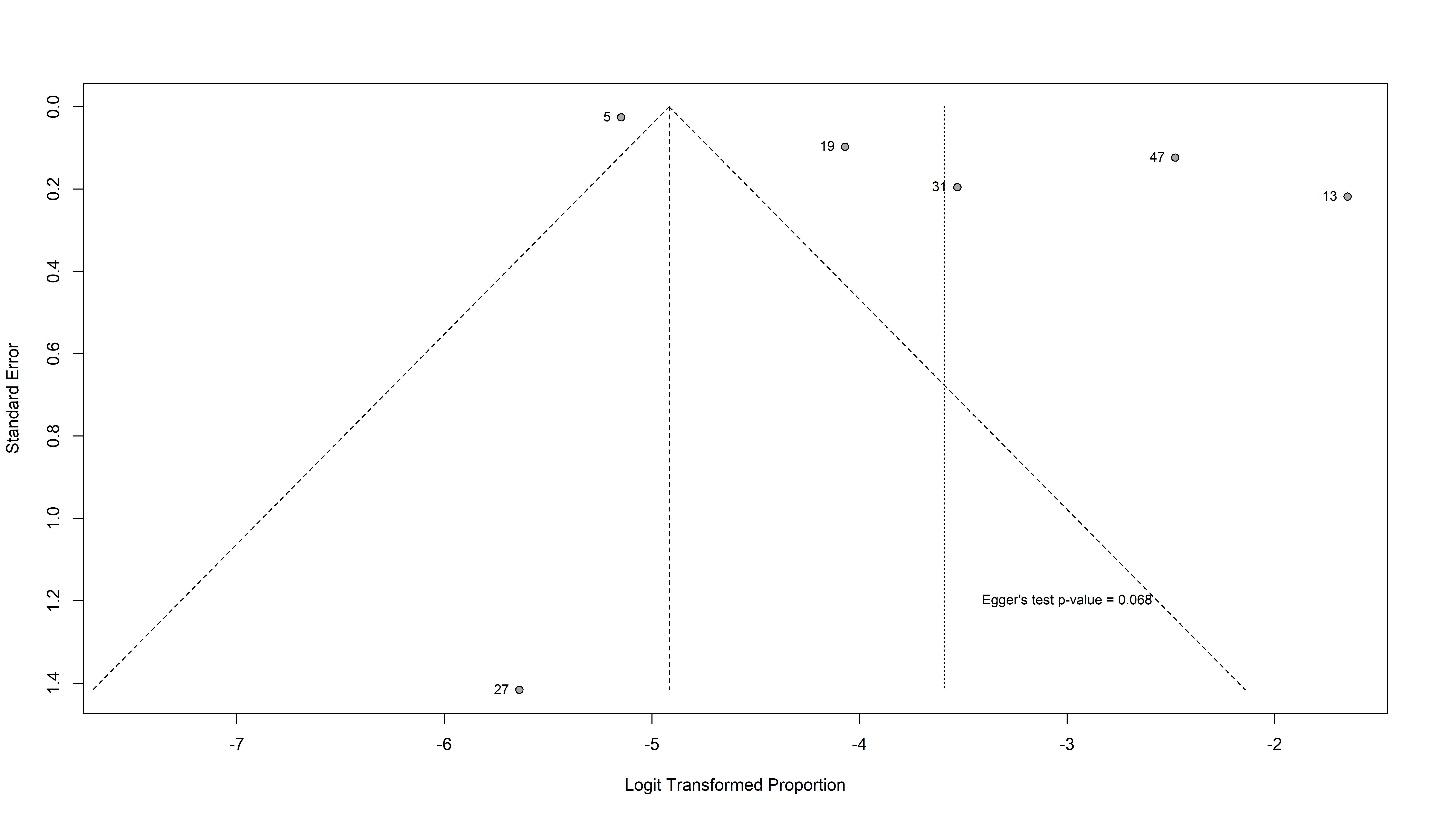

**Stroke**
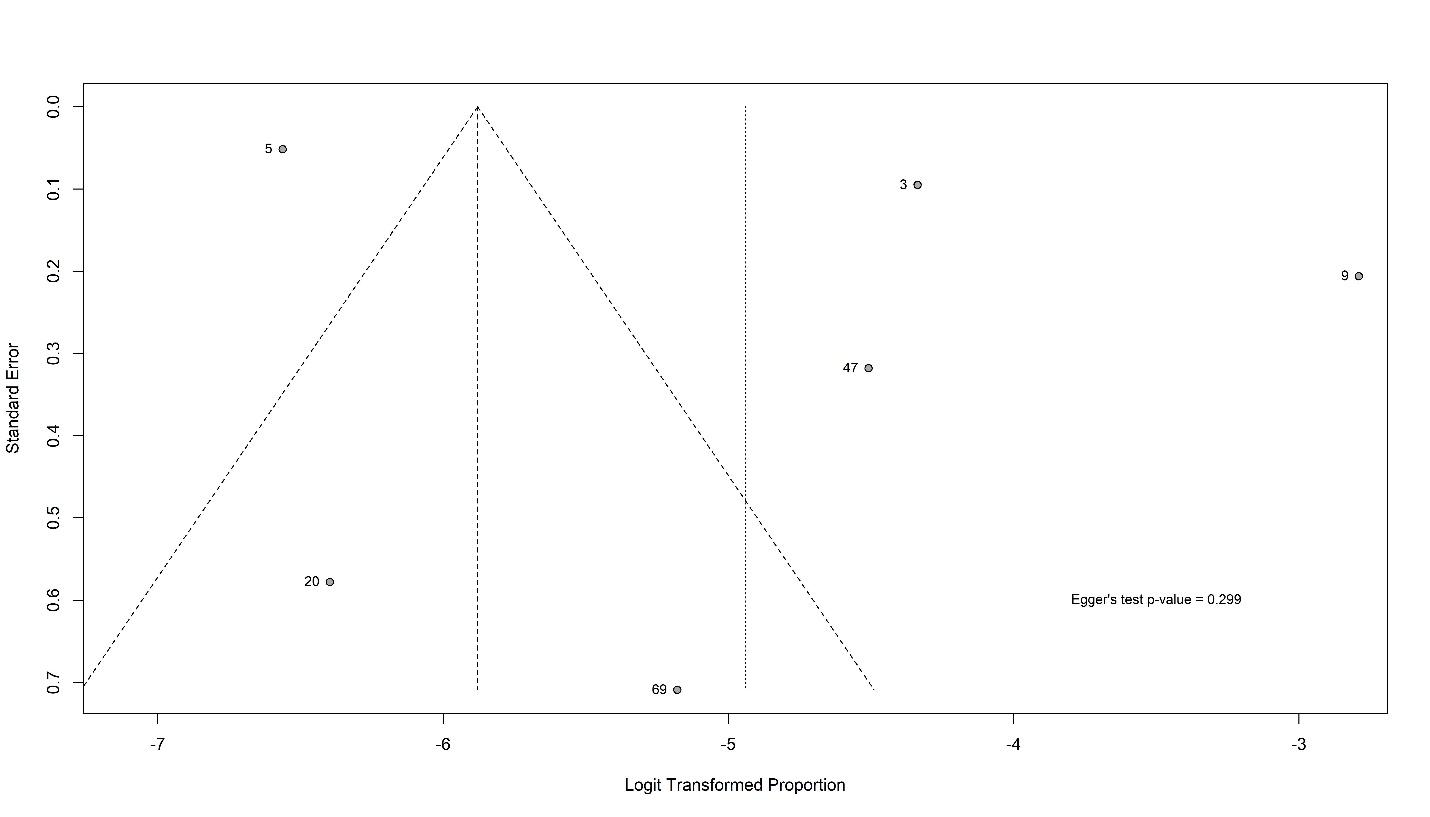

**Heart failure**

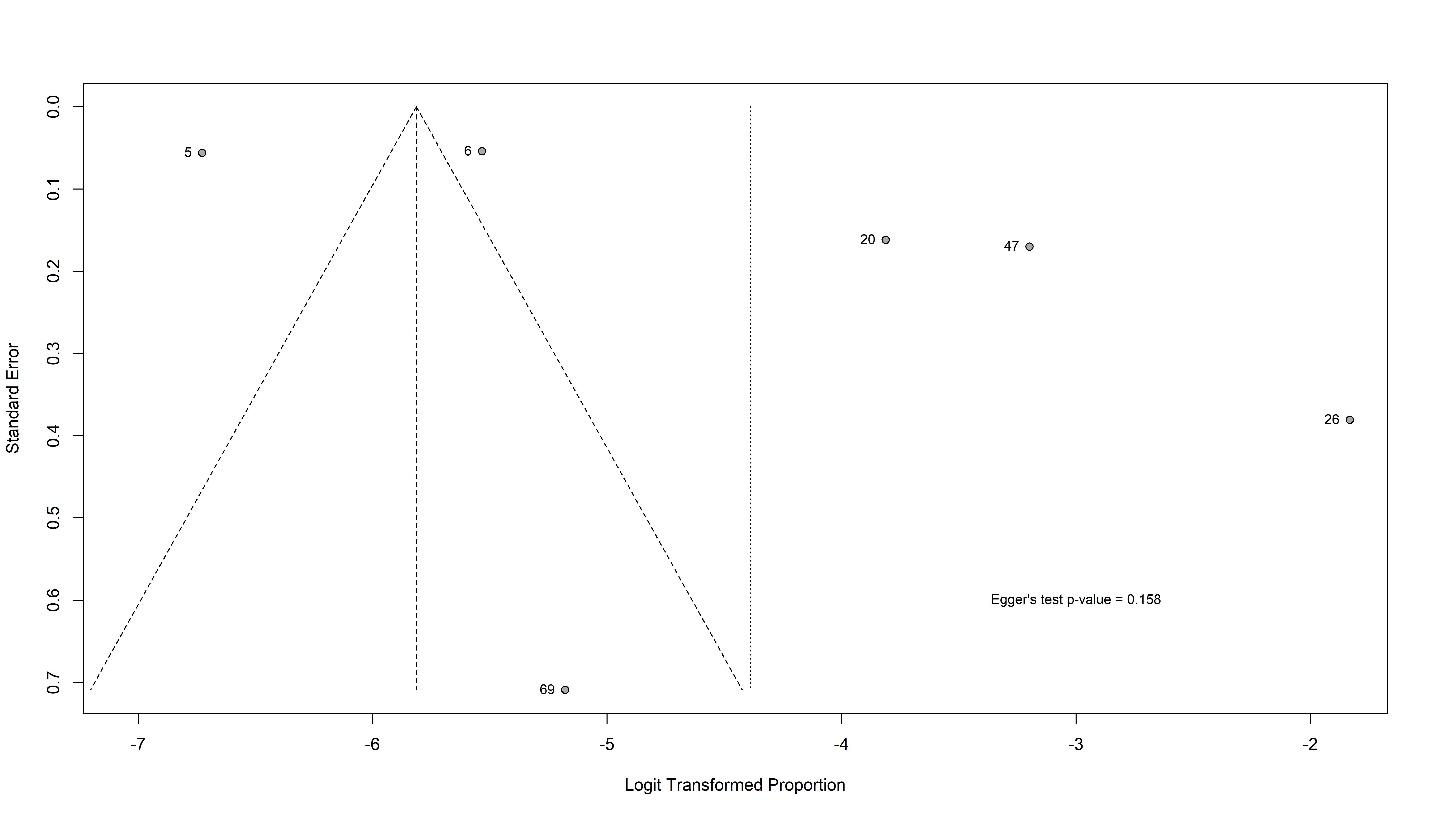

**Coronary disease**

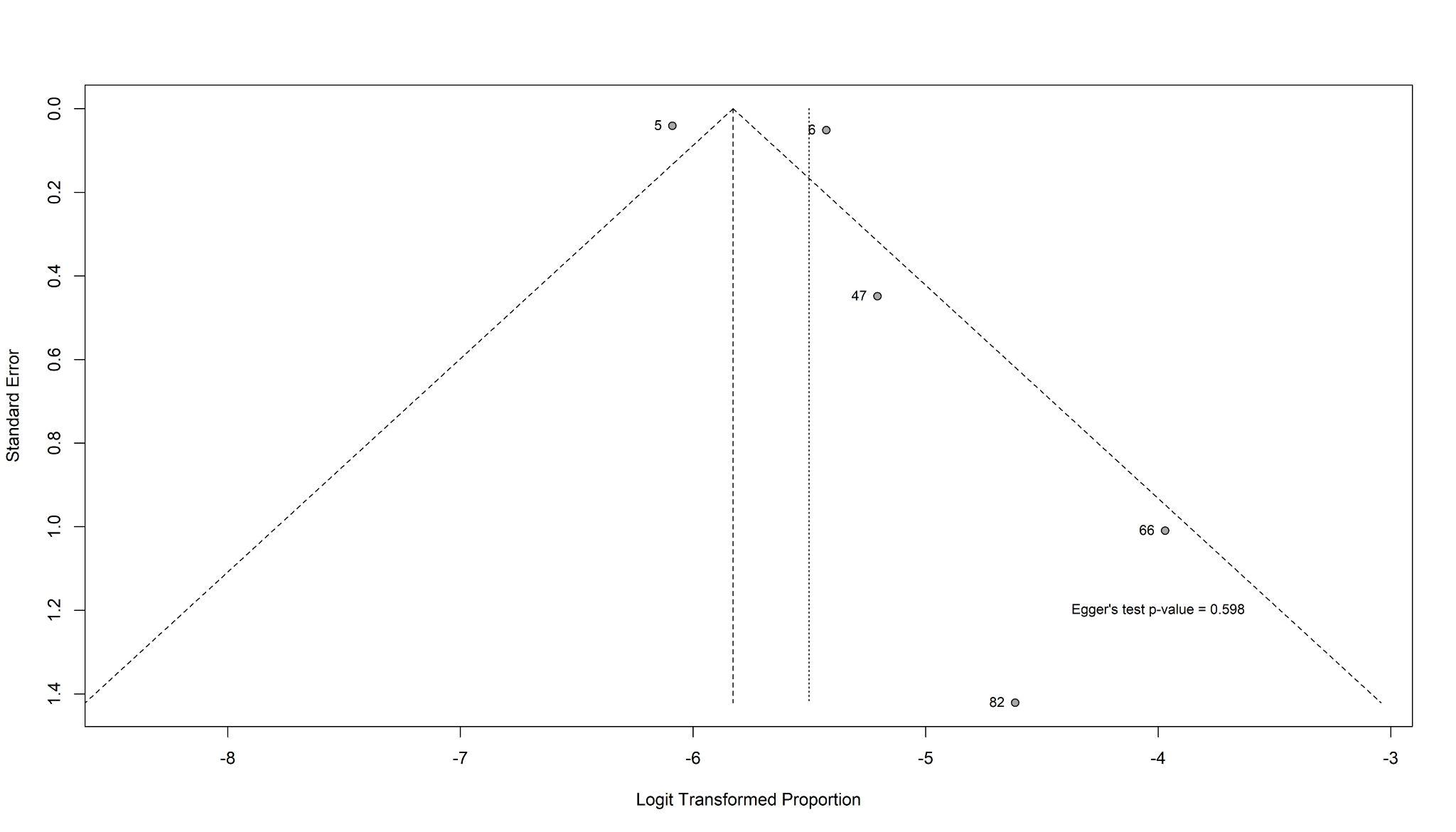

**Myocarditis**

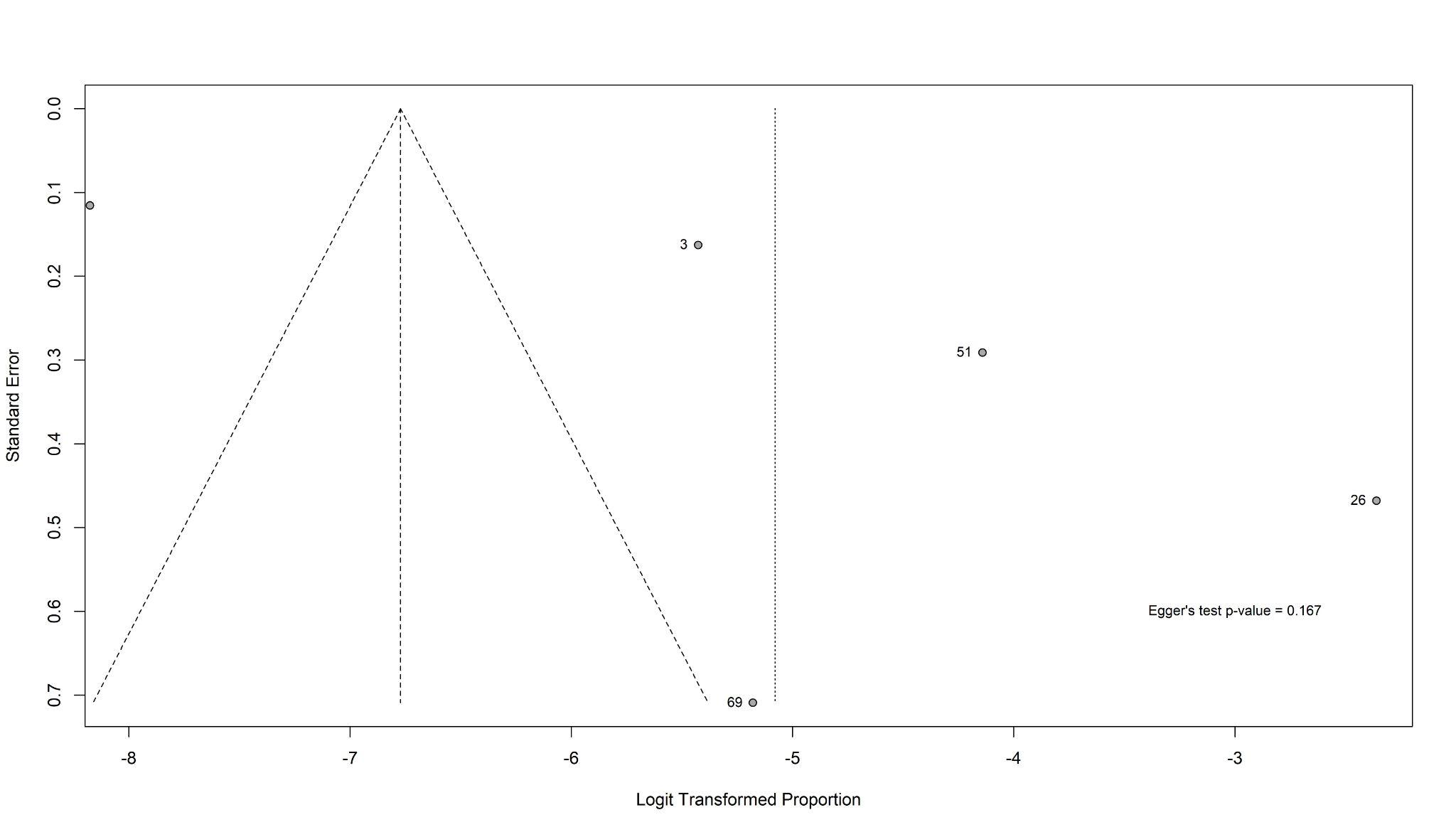
